# Systems Pharmacology Model Predicts Zinc and Copper Can Be Repurposed as Endometriosis Therapies

**DOI:** 10.1101/2025.10.06.25337149

**Authors:** Pablo Garcia-Acero, Francisco Jose Sanz, Patricia Sebastian-Leon, Almudena Devesa-Peiro, Antonio Parraga-Leo, Patricia Diaz-Gimeno

**Author notes:** **Corresponding author** Patricia Diaz-Gimeno IVIRMA Global Research Alliance, IVI Foundation, Instituto de Investigación Sanitaria La Fe (IIS La Fe), Valencia, Spain IVI Foundation, Edificio Biopolo – Instituto de Investigación Sanitaria la Fe Avenida Fernando Abril Martorell, 106 - Torre A, Planta 1ª 46026 Valencia /.

## Abstract

**BACKGROUND:** Endometriosis is a complex and polygenic disease characterized by the growth of endometrial-like tissue outside the uterus. Associated conditions include pelvic pain and infertility among others. Current treatments of endometriosis are not effective in many patients and are prone of side effects, being laparoscopy the main solution for them. However, laparoscopy is an invasive solution that can negatively affect ovarian reserve among others fertility consequences. Therefore, more effective treatments are needed for inhibiting the disease progression. Some approaches as systems pharmacology can offer new avenues to study complex diseases at a systematic level, allowing us the integration of information at different levels for prioritizing effective therapies.

**OBJECTIVE:** To use systems pharmacology and *in vitro* analyses to predict more effective endometriosis treatment among approved drugs on the market for further clinical trials.

**STUDY DESIGN:** Data-driven discovery study integrating protein-protein interactions, transcriptomics, approved drugs and endometriosis-related gene targets to predict therapies that can be repositioned in endometriosis. An *in-silico* molecular drug efficacy screening was performed to compare proposed treatments with the current ones. The candidate drugs with the safest clinical profiles were prioritized for *in-vitro* validation in an endometriosis cell model.

**RESULTS:** Our endometriosis molecular model showed that the endometriosis progression and the infertility associated are molecularly highly related since they share 210 genes through direct protein-protein interactions from 553 infertility-related and 2,931 progression-related endometriosis genes, an overlap that is higher than randomly expected (p-value = 1.9 × 10⁻¹¹). In addition, these genes are also closer physically interacting in the network than random expectation (z-score = −1.75, p-value = 0.03). Both results highlighted the similar molecular basis of endometriosis progression and infertility associated to endometrium. Leveraging this endometriosis disease network, we have identified sixteen potential candidate drugs, which had a superior molecular efficacy treating the disease genes as they target more endometriosis-related genes (p-value = 0.01) than randomly expected compared with current endometriosis treatments (p-value = 0.97). The drug safety analysis deemed adenine, copper, zinc, NADH, glutathione, and resveratrol as the safest candidate drugs for endometriosis, with no or hardly any clinical side effects. Zinc, copper and resveratrol suppressed endometriosis-related phenotypes in 12Z cells, including cell proliferation, migration, and overexpression of endometriosis-related biomarkers (*ERB*, *IL-6* and *VEGF)*.

**CONCLUSION:** This approach highlights the nutraceutics zinc and copper for having superior molecular efficacy than current endometriosis treatments, no considerable side effects, and the ability to reduce endometriosis cell phenotypes. Long-term clinical trials will be needed to confirm women *in vivo* effectiveness and provide new treatment opportunities for refractory endometriosis.

**Condensation page:** *Tweetable Statement:* A drug repurposing approach based on systems pharmacology and *in vitro* validation predict that zinc and copper nutraceuticals target 76 endometriosis-related genes, reverse endometriosis phenotypes, and exceed efficacy of current treatments

**AJOG at a Glance:** *A. Why was this study conducted?:* Current endometriosis treatments do not effectively address root causes of the disease and have side effects. Systems pharmacology-based models help uncover existing drugs that can be repurposed for complex diseases such as endometriosis.

*B. What are the key findings?:* A systems pharmacology-based model prioritized sixteen approved pharmacotherapies with superior molecular efficacy than current endometriosis therapies. Experimental validation of three of the safest prioritized compounds (resveratrol, copper, and zinc) showed a significant reduction of endometriotic cell phenotypes.

*C. What does this study add to what is already known?:* There are several opportunities to improve the standard of care for patients suffering endometriosis with existing drugs on the market. Multi-target pharmacotherapies are likely more effective and safer than current endometriosis treatments and surgical interventions.

## Introduction

Endometriosis is a complex and multifactorial disease that affects up to 10% of reproductive-age women.^1^ Its phenotype is influenced by both environmental and genetic factors, with an associated heritability of 47%.^2^ Endometriosis is characterized by the growth of endometrial-like tissue outside of the uterus (ectopic endometrium), which is classified into four stages (I, II, III and IV) depending on lesion severity, size, depth, location, and quantity.^3,4^ Endometriotic progression is associated with pelvic pain, dysmenorrhea, gastrointestinal discomfort, dyspareunia and infertility.^5^ Although the exact etiology remains elusive,^3,6^ several studies have found that endometrial biopsies from patients with endometriosis present functional dysregulations related to apoptosis, angiogenesis, cell proliferation, adhesion, inflammation and oxidative stress.^7,8^

Endometriosis is often treated first line with combined oral contraceptives followed by gonadotropin-releasing hormone (GnRH) agonists in second line. As these therapies are counterproductive for women who desire pregnancy,^9^ alternative therapies, such as GnRH antagonists, aromatase inhibitors, or selective estrogen and progesterone receptor modulators are being investigated. However, all these treatments have limited efficacy due to their palliative uses and clinical side effects. Invasive laparoscopy remains the most effective solution to halt progression of endometriosis but negatively affect the ovarian reserve.^10^ Therefore, there is an urgent need for new approaches considering the complexity and multifactorial nature of endometriosis.

In this context, multidisciplinary frameworks leveraging systems pharmacology, network medicine, and bioinformatics can study targets in the underlying molecular signaling networks, and prioritize drugs targeting the most influential genes.^16^ Alternatively, drug repurposing aims to find new indications for approved drugs on the market, saving time and costs compared to classical drug development.^17^

This study aimed to propose safe and potentially more effective pharmacotherapies for endometriosis using an *in silico* systems pharmacology platform integrating drug and molecular information of endometriosis pathophysiology. We prioritized candidates based on their predicted molecular efficacy in endometriosis, clinical safety profiles, and validated their ability to suppress endometriosis cell phenotypes *in vitro*.

## Materials and methods

### Study design

The study design is represented in **Figure 1**. Endometriosis-related genes were prioritized using differential expression analysis of transcriptomic data from endometrial samples from healthy women, and eutopic and ectopic samples from patients with endometriosis. Genes differentially expressed in eutopic vs. healthy endometrial samples were defined as endometriosis infertility-related genes, while genes differentially expressed in ectopic vs. eutopic endometrial samples were defined as endometriosis disease progression genes. All endometriosis-related genes were mapped in the protein-protein human interactome to build the endometriosis disease network (EDN). Then, promiscuous drugs were prioritized based on the importance of their targets in the EDN.^18–20^ Next, the drugs’ clinical safety profiles were ranked using the quantity and severity of reported side effects. Finally, candidate drugs deemed to have the safest profiles were experimentally validated in an *in vitro* endometriosis model.

## Systems pharmacology model for drug repurposing in endometriosis

### Ethics statement

This *in silico* study used raw endometrial gene expression data and patient meta-data available in the Gene Expression Omnibus (GEO) data repository.^21^ Following GEO policies, patient data were anonymized and encrypted, thus no additional institutional review board approval was required to access or process the data.^22^ (Figure 1A).

### Endometriosis disease network (EDN)

The human interactome used in this study was built by aggregating protein-protein interactions (PPIs) datasets from the Human Reference Protein Interactome Mapping Project (HuRi).^23^

Genes related to endometriosis were identified using two systematic reviews in GEO, as described in Devesa-Peiro *et al.* (2020). These searches focused on studies comparing intrapatient ectopic and eutopic tissues to identify endometriosis-related progression genes; eutopic tissue from endometriosis patients vs. endometrial tissue from healthy women to identify infertility-associated genes. Next, the datasets related to progression and infertility were integrated separately, as described in Tajti *et al.* (2020) (see Supplemental Methods). Differential expression analyses were performed independently on endometriosis progression and infertility datasets using the *limma* R-package^25^ to prioritize genes associated to them.

A one-sided hypergeometric test was performed to evaluate overlapping between target genes related to both endometriosis progression and infertility. A proximity analysis^26^ was done to study the biological relationship between progression and infertily-related endometriosis genes and search for common treatments (see Supplemental Material).

Finally, EDN was created by filtering the human interactome for direct physical interactions between the prioritized endometriosis disease genes. The degree (number of connections for a given gene in the disease network) distribution of EDN genes was analyzed using Pearson correlations to confirm that they followed a power-law distribution, hence confirming its biological nature.^27^) (see Supplemental Material).

### Endometriosis drug prioritization

To prioritize potential candidate drugs to repurpose in endometriosis, all approved drugs (n=3,883), and their human target genes (n=2,479) were extracted from the DrugBank database.^28^ After the target genes were mapped into the EDN, the degree and betweenness centrality (number of shortest paths between any two genes that pass through a given gene) for all genes on the EDN was computed using Cytoscape.^29^ The degree identifies genes with a high connectivity with other endometriosis genes and the betweenness identifies genes with a high influence in the EDN network. Drugs were prioritized based on these network parameters together with drug promiscuity (see Supplemental Material) (Figure 1B).

### *In silico* evaluation of drugs’ molecular effectiveness and safety profiles

We systematically searched for current endometriosis therapies in guidelines of the European Society of Human Reproduction and Embryology (ESHRE),^10^ American Society for Reproductive Medicine (ASRM), Human Reproduction (HR),^30^ Spanish Fertility Society (SEF), and Spanish National Health System (SNS).

A proximity analysis was performed to compare the molecular efficacy between candidate and current endometriosis drugs *in silico*. This analysis allows able to distinguish between palliative and effective drugs.^31^ (See supplementary Material).

Finally, the quantity and severity of side effects were manually extracted from available drug data sheets published online by the Spanish Agency of Medicines and Medical Devices (AEMPS).

## *In vitro* validation of candidate drugs in an endometriosis cell model

### Ethical approval

This prospective study was approved by the IVI Research Ethics Committee, Valencia, Spain (1706-FIVI-048-PD). Written informed consent was obtained from all participants.

### Endometrial biopsy collection and processing

For gene disease prioritization we used GEO gene expression samples files. For drug validation in endometrium, endometrial biopsies (n=6) were collected from volunteers undergoing hormone replacement therapy, using a Pipelle catheter (CCD Laboratories, Paris, France) on day P+5 to isolate the human endometrial epithelial cells (HEECs) as previously described in.^32^

### Cell cultures and viability assays

HEECs and immortalized human epithelial-like endometriotic 12Z cells (Applied Biological Materials, Canada, T0764) were cultured in Dulbecco’s Modified Eagle Medium/Nutrient Mixture F-12 (DMEM/F12) / MCDB 131 Medium (Sigma-Aldrich, Madrid, Spain) (3:1) supplemented with 10% fetal bovine serum (FBS) (Sigma-Aldrich), 0.4 μg/mL amphotericin B (Thermo Fisher Scientific, Waltham, USA), and 80 μg/mL gentamicin (Thermo Fisher Scientific) at 37°C and 5% CO_2_.

HEECs and 12Z cells were seeded in a 96-well plate at a density of 1.5×10^4^ cells/well and cultured with the medium described above supplemented with 10– 500 µM NADH (Santa Cruz Biotechnology, Texas, USA), 50–800 µM adenine (Santa Cruz Biotechnology), 10–500 µM copper(II) sulfate pentahydrate (Sigma-Aldrich), 10–500 µM glutathione (Santa Cruz Biotechnology), 10–500 µM zinc acetate dihydrate (Sigma-Aldrich) or 10–500 µM resveratrol (Santa Cruz Biotechnology) for 24h. 0.1% DMSO was used as a vehicle.

Cell viability was measured using the CellTiter 96^®^ AQueous One Solution Cell Proliferation Assay (MTS) (PROMEGA, Wisconsin, USA) following manufacturer’s instructions. Absorbance intensity was measured at 490 nm using a SpectraMax® 190 (bioNova cientifica S.L., Madrid, Spain). All assays were performed in triplicate (Figure 1C).

### qRT-PCR

Total RNA was extracted from treated 12Z cells using the RNeasy Mini Kit (QIAGEN, Hilden, Germany) following manufacturer’s instructions. RNA was reversed transcribed using PrimeScript^TM^ RT Reagent Kit (TaKaRa, Japan). Quantitative real-time polymerase chain reaction (qRT-PCR) was performed using PowerUp^TM^ SYBR^TM^ Green Master Mix (Applied Biosystems, Massachusets, USA) on an ABI StepOnePlus^TM^ Real-Time PCR System (Applied Biosystems). Gene expression was normalized to *β-actin*. Specific primers used for qRT-PCR are listed in **Supplemental Table 1**.

### *In vitro* scratching assay

To evaluate if the proposed treatments hindered cell migration, one of the main phenotypic characteristics of ectopic endometriosis cells, we performed an *in vitro* scratching assay. Endometriotic 12Z cells were seeded in a 24-well plate and cultured until a monolayer was formed. A micropipette tip was used to scratch a straight line in the cell layer. After washing cells with phosphate buffer solution (PBS), cells were treated with 125 µM zinc or 40 µM copper for 24h. Images were captured by microscope (Zeiss AX10) at 0 and 24h after the scratch. These assays were performed in triplicate (Figure 1C).

### Statistical analysis

Statistical analyses were performed in R and visualized using the *ggplot2* R-package.^33^ Significant differences between means were assessed using t-tests or ANOVA with post-hoc Tukey test. In all cases, p-values < 0.05 were considered statistically significant.

## Results

### Endometriosis disease network simultaneously target endometriosis progression and associated infertility allowing the searching for common treatments

A total of 4,176 differentially expressed genes (DEGs) related to endometriosis progression and 831 DEGs related to infertility-associated endometriosis were compiled (see supplemental file results section) (**Supplemental Figure 1A-B; Supplemental Table 2**). Notably, we found a significant overlap of 210 shared genes (p-value = 1.9×10⁻¹¹) and proximity between progression-related and infertility-related endometriosis genes (z-score = −1.75, p-value = 0.03), confirming that they share molecular pathways, which allows the repurposing of existing treatments on the market for both conditions (**Supplemental Figure 1C**). Finally, The EDN was built integrating all genes to prioritize approved drugs that can specifically target endometriosis progression and/or associated infertility (see supplemental file results section for specific information of the EDN).

### Sixteen approved drugs may be repurposed for endometriosis treatment

We prioritized drugs targeting influential proteins of the EDN that: (i) had the most physical interactions with other endometriosis proteins (130 drugs with a network degree > 13), (ii) were central communicators between protein groups (188 drugs with a betweenness > 0.0025), and (iii) targeted multiple endometriosis proteins (41 drugs with a promiscuity > 3) (**Supplemental Table 4**). Sixteen prioritized drugs met all three criteria and were selected as candidate drugs to repurpose in endometriosis (**Figure 2**).

### Drug repurposing may improve molecular efficacy of endometriosis treatment

Reproductive medicine guidelines reported 44 pharmacotherapies are currently being used to treat endometriosis (**Supplemental Table 5**). These endometriosis drugs collectively had 85 gene targets annotated in DrugBank. Nineteen of these targets (22.3%) were present in our endometriosis disease network (15 progression-related and 4 infertility-related endometriosis genes). Proximity analyses showed that these current pharmacotherapies have significantly longer distances in the drug–target interaction network than our 16 candidate drugs (z=3.95 vs. 3.78, p-value=0.97 vs. 0.01, respectively), reinforcing the therapeutic potential of improving endometriosis treatment with our prioritized compounds according to the molecular effect in relation with current therapies.

### Adenine, resveratrol, zinc, copper, NADH, and glutathione are proposed as the safest candidate drugs to repurpose for endometriosis treatment

To narrow the candidate drugs for *in vitro* validation, we focused on their clinical safety profiles. We ranked compounds according to the quantity and severity of side effects reported in AEMPS drug data sheets (**Figure 2B**). The six candidate drugs (i.e., copper, resveratrol, adenine, glutathione, NADH and zinc) with no considerable side effects reported were selected for *in-vitro* validation, only zinc presented as a description the side effect of nausea.

### Zinc, copper and resveratrol reduce endometriosis cell proliferation *in vitro*

To develop a treatment that is cytotoxic to proliferating endometriotic cells but not in healthy HEECs, we first established the maximal tolerable dose of each drug in HEECs using dose-dependent cell viability assays (**Supplemental Figure 2**). Viable HEECs in proliferation were most sensitive to copper from 50 µM (p-value = 5.48 x 10⁻⁴). Meanwhile, zinc and resveratrol were not significantly cytotoxic until 200 µM (p-value = 1.6 x 10⁻⁴ and p-value = 0.0018, respectively). Adenine, NADH and glutathione did not significantly reduce HEEC viability at any of the doses tested.

Remaining under the maximal tolerable drug doses established for HEECs, we then determined the concentrations required to effectively reduce viability of proliferating endometriotic cells. We found that 150 µM zinc treatment reduced 12Z cell viability by 85.07%, without significantly affecting HEECs. Although concentrations of zinc ≥175 µM were highly effective at reducing viability of 12Z cells, they impaired also HEEC viability by 24.92% at 175 µM (p-value < 0.05) and 50.42% at 200 µM (p-value < 0.001) **(Figure 3A)**. Alternatively, copper was detrimental to cell viability with 20 µM doses in both HEECs and 12Z cells.

However, 12Z cells and HEECs were differentially sensitive to 40 µM (36.15% vs. 61.2%, respectively) and 50 µM doses (38.23% vs. 81.6%, respectively) (**Figure 3B**).

On the contrary, 800 µM adenine, 500 µM glutathione, and 500 µM NADH did not reduce viability of healthy endometrial cells (**Supplemental Figure 2**) or 12Z cells (**Figure 3C**), suggesting that these treatments would not effectively prevent the progression of endometriosis. Indeed, concentrations of glutathione and NADH over 500 µM favored proliferation of 12Z cells (data not shown), suggesting that these compounds could promote the progression of endometriosis.

Resveratrol treatments for endometriosis were widely studied in several preclinical models, such as 12Z cells^40,41^ and other endometriosis models.^42–51^ We observed that 100 µM resveratrol also significantly decreased 12Z cell viability by 67.1% compared to the vehicle control (**Figure 3D**).

### Zinc and copper reduced endometriosis-related phenotypes in 12Z cells

Next, we evaluated whether zinc and copper treatment targeted overexpressed genes in endometriosis cells using qRT-PCR. Both zinc and copper significantly reduced the relative mRNA expression of estrogen receptor 2 (*ERB)* (FC = −2.15, p-value = 2.25 x 10⁻⁴ and FC = −1.30, p-value = 0.003, respectively), interleukin 6 (*IL-6)* (FC = −2.63, p-value = 0.012 and FC = −4.64, p-value = 0.002, respectively) and vascular endothelial growth factor (*VEGF)* (FC = −1.83, p-value = 0.001 and FC = −1.24, p-value = 0.005, respectively) in 12Z cells. However, no significant effect was found in the relative mRNA expression of cytochrome P450 family 19 subfamily A member 1 (*CYP19A1)*, cyclooxygenase-2 (*COX2)* or tumor necrosis factor alpha (*TNF-α)* (**Figure 4A**).

Lastly, the scratching assay revealed that the migration capacity of 12Z cells was significantly reduced after zinc (86.8% vs. 69.5%; p-value = 0.006) or copper treatment (85.7% vs 58.1%; p-value = 0.007), compared to the vehicle control (**Figure 4B**).

## Comment

### Principal findings

Sixteen pharmacotherapies with multiple target genes related to endometrial physiopathology and/or infertility on the market that are more effective than existing endometriosis treatments were highlighted. *In vitro* validation of the six safest compounds showed zinc, copper and resveratrol differentially reduced proliferation of endometriosis cells over healthy endometrial cells, supporting their potential to be repurposed for endometriosis therapies. We observed that zinc and copper reduced the overexpression of genes associated with oxidative stress, the estrogen response and immune system regulation in endometriosis (i.e., *ERB*, *IL-6* and *VEGF)*. Moreover, both compounds impaired the cell migration capacity of endometriotic cells, a characteristic manifestation of ectopic endometriosis cells.

### Results in the context of what is known

Studying molecular interaction networks has been proposed as an outstanding strategy to improve understanding of complex diseases such as endometriosis. Compiling the transcriptomic data from the endometrium of healthy women with that of eutopic and ectopic endometrium from women with endometriosis, we constructed the EDN based on the molecular proximity and overlap of progression-related and infertility-related genes. In the future, our EDN could be used to discover additional biomarkers for endometriosis and/or propose new treatments.^16^ Leveraging this strategy, we have proposed a set of drugs as novel candidate compounds for endometriosis.

Reduced blood levels of zinc have been associated with the development of endometriosis in an infertile population.^52^ Interestingly, women with endometriosis were found to have a lower zinc intake in their diet than healthy women,^53^ while women that got pregnant presented higher zinc levels in follicular fluid than infertile women.^54^ These studies support our predictions that zinc might not only be effective for treating endometriosis onset and/or development, but also for the associated infertility.

The use of copper in endometriosis remains controversial. Aligning with our *in silico* predictions, previous studies related copper deficiency to endometriosis and infertility. Infertile women with endometriosis presented lower copper levels in peritoneal fluid than infertile women with unexplained infertility.^58^ Low copper blood levels were found in women experiencing infertility^59^ or miscarriage.^60^ However, some groups have reported a detrimental effect of copper. Turgut *et al.* (2013) found a positive correlation between copper levels and the etiopathogenesis of endometriosis in women. Meanwhile, Conforti *et al.* (2023) demonstrated that a copper chelator reduced the volume and weight of endometriotic lesions in TNFR1-deficient mice. In both studies, the harmful effect of copper in endometriosis was associated with elevated oxidative stress.

Finally, our *in silico* study identified resveratrol as a promising candidate compound. Interestingly, resveratrol is widely studied and was already proposed for treating endometriosis. Our *in vitro* results support those of two recent studies reporting that resveratrol reduces cell viability, cell migration, and induces apoptosis in 12Z endometriotic cells.^40,41^ Additionally, *in vivo* experiments in rats and mice demonstrated that resveratrol reduces endometriotic lesions.^63–65^

### Clinical implications

Current treatments for endometriosis are palliative and show important side effects as menopausal symptoms, menstrual irregularities, or even reduce ovarian reserve in case of laparoscopy. They focus on the consequences of the condition instead of addressing the underlying causes. In addition, laparoscopy, the most effective therapy nowadays, is invasive and negatively affects the ovarian reserve^68^ and other fertility aspects. Therefore, new strategies addressing this disease from a global perspective are required.

In this study, we propose zinc and copper as potential endometriosis therapies, based on their clinical safety as nutraceuticals and predicted effectiveness for targeting multiple genes involved in endometriosis progression and associated infertility. Our results suggest that the candidate compounds might be more effective than standard endometriosis therapies due to the more direct interactions with genes associated to the disease. As these compounds are already approved for other indications, repurposing these drugs reduces the time and cost of clinical trials, accelerating translation into clinical practice.^70^ Moreover, zinc and copper are nutraceuticals that were already validated *in vitro*, supporting safe clinical implementation in women with endometriosis.^71,72^

### Research implications

Emerging approaches like network pharmacology are shifting the paradigm to map out drugs or combinations of drugs that address the root cause of diseases from a holistic medical perspective,^74^ while optimizing efficacy and safety.

Notably, network pharmacology was recently employed to propose new therapies for cancer^75,76^ and cardiovascular diseases,^77^ among others. These promising findings encourage the use of technology to repurpose drugs for complex diseases like endometriosis.

Our EDN highlighted 16 candidate drugs with a potential to improve standard of care for endometriosis. Among these, copper and zinc had the safest clinical profiles and highest efficacy in an endometriosis cell model.

### Strengths and Limitations

Among the strengths of this work, our *in silico* analysis compiled independent findings from seven accurate molecular datasets to provide a robust overview of the pathophysiology of endometriosis.^24^ Although the human interactome consulted to build the EDN included only 11,517 out of the estimated 20,000 human protein-coding genes at the time of this study, the high level of evidence curation ensures that our predictions are reliable. Plus, the scale-free nature of the current human interactome, ensures that it represents a real biological network.

While our *in vitro* validations provide evidence that the candidate drugs are effective, a clinical trial performed in endometriosis patients are required to validate the effectiveness of zinc and copper as repurposed endometriosis therapies. In addition, it would be necessary to determine the adequate posology for both drugs in such clinical trial. Nevertheless, identifying resveratrol as a candidate reinforces our results, as this widely studied drug was proposed as a new treatment following *in vitro* and *in vivo* validations.^40,41,78–80^ Moreover, treatment regimens combining resveratrol and contraceptives effectively reduced pain in women with endometriosis.^66^

### Conclusions

A network pharmacology approach identified several existing drugs that can be repurposed for endometriosis. The EDN provided a holistic perspective of the molecular complexity of endometriosis, integrating omics information related to existing drugs, pathophysiology and related infertility to identify drugs which can treat the root causes of the disease rather than the symptoms like current therapies. Among the proposed and *in-vitro* validated candidates to repurpose in endometriosis, we highlight zinc and copper, which are both essential minerals commercialized as nutraceuticals. As these compounds are already approved for other indications, and have high clinical safety profiles, given the evidence of this paper we propose to carry out a clinical trial aimed to repurpose them, a strategy that reduces clinical trial duration and cost, accelerating the translation into clinical practice.

## Data Availability

All data produced in the present study are available upon reasonable request to the authors

## Acknowledgements

The authors thank IVIRMA Global and the Instituto de Salud Carlos III (ISCIII) for their research support, all the study participants who donated endometrial biopsies, and the clinical staff who recruited and managed the participants. Also, authors thank Rosa Ferrando, Maria Jose Herrero Cervera and Salvador Francisco Aliño Pellicer for their help and support in this work.

## Author contribution

Pablo Garcia-Acero: Methodology, Formal analysis, Investigation, Writing - Original Draft, Writing – Review & Editing, Visualization. Francisco Jose Sanz: Methodology, Formal analysis, Investigation, Writing - Original Draft, Writing - Review & Editing, Visualization. Patricia Sebastian-Leon: Methodology, Software, Formal analysis, Data Curation, Writing - Original Draft, Writing - Review & Editing. Almudena Devesa-Peiro: Formal analysis, Conceptualization, Writing - Original Draft. Antonio Parraga-Leo: Software, Formal analysis, Data Curation. Patricia Diaz-Gimeno: Conceptualization, Supervision, Methodology, Writing - Review & Editing, Project administration, Funding acquisition.

Part of the results included in the paper have been presented in the e 72^nd^ Annual Scientific Meeting of the Society for Reproductive Investigation (SRI, 2025) in a poster entitled “Drug repurposing in endometriosis: zinc and cooper as potential endometriosis therapiesafter being prioritized by systems pharmacology”.

---

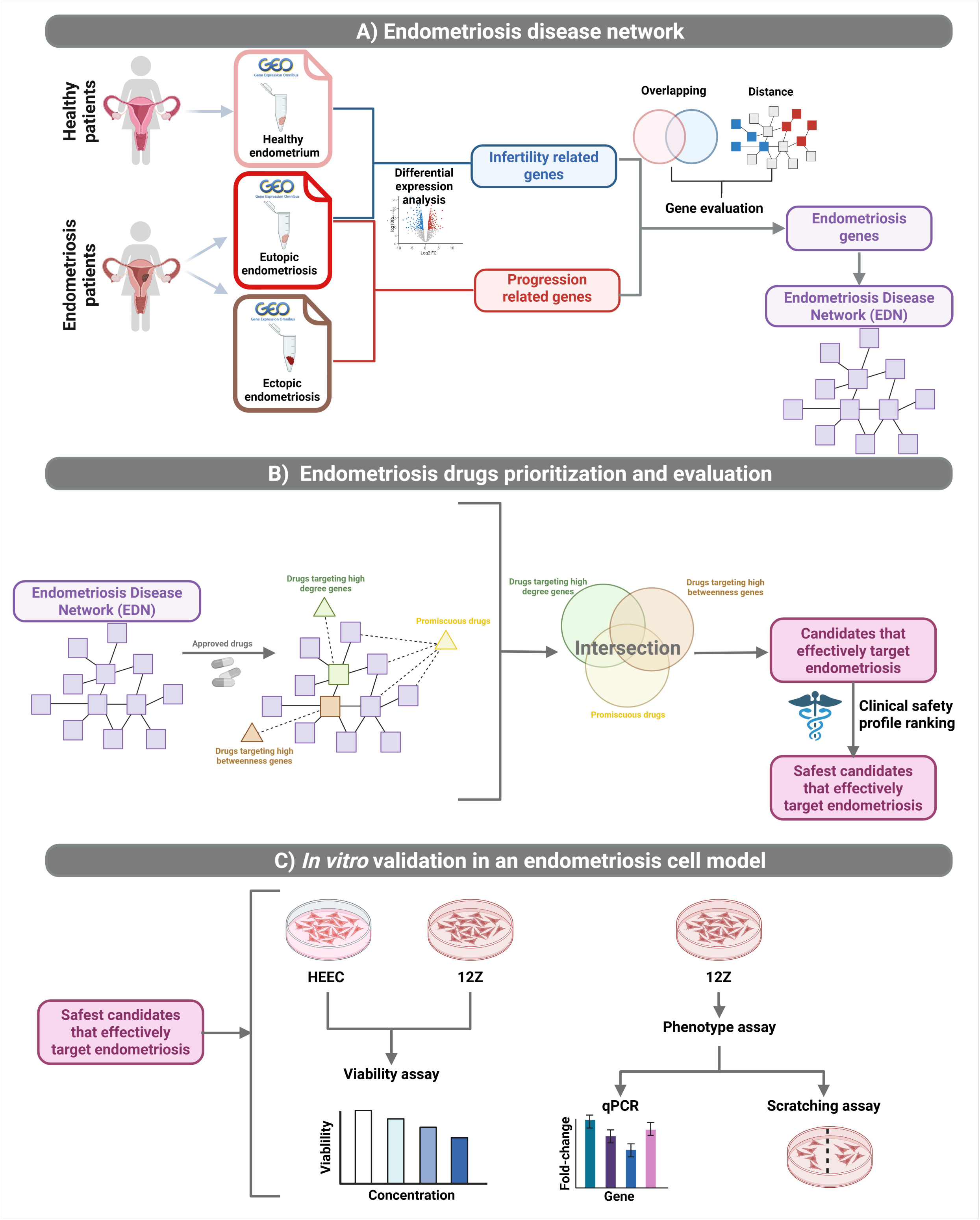

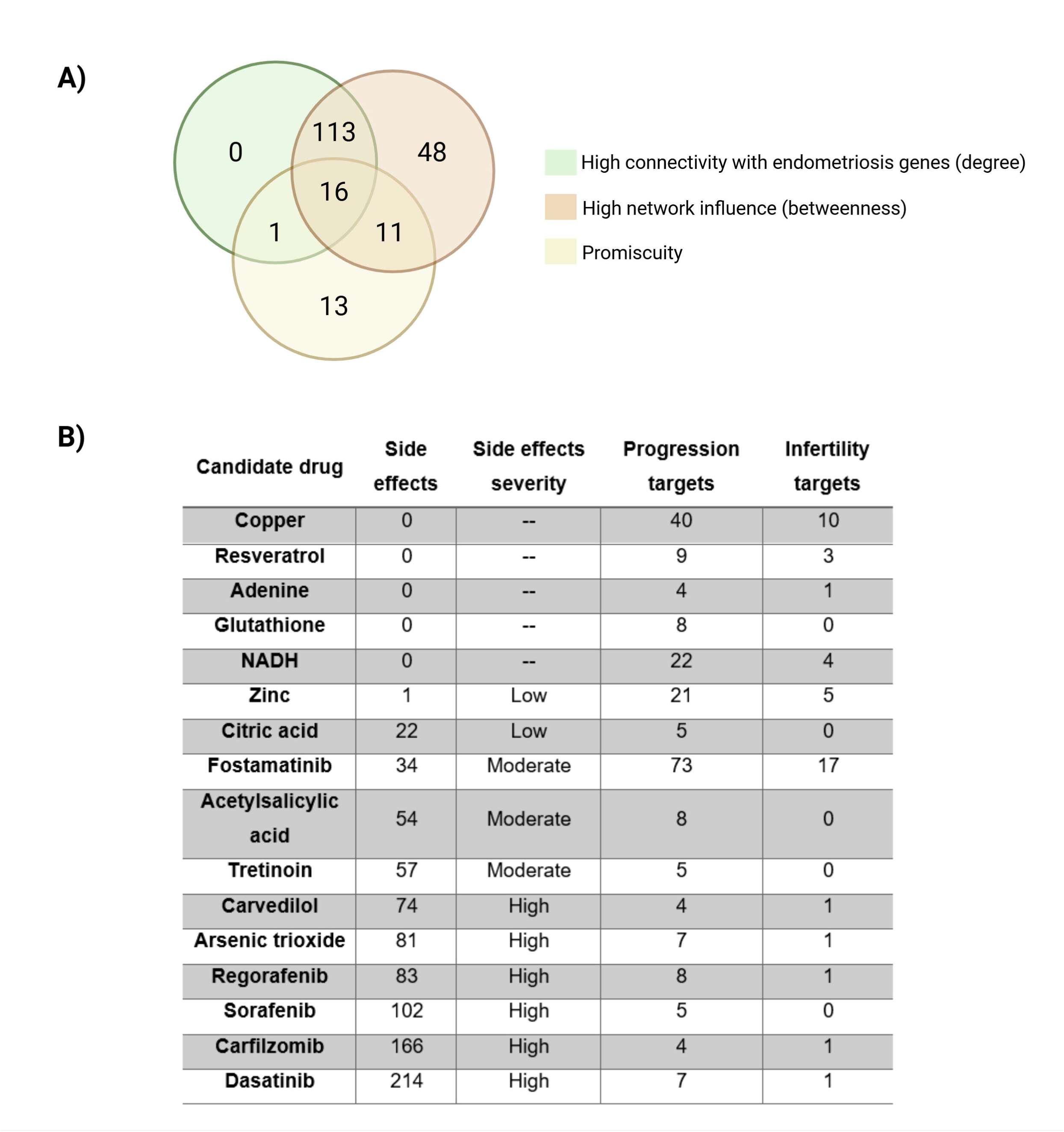

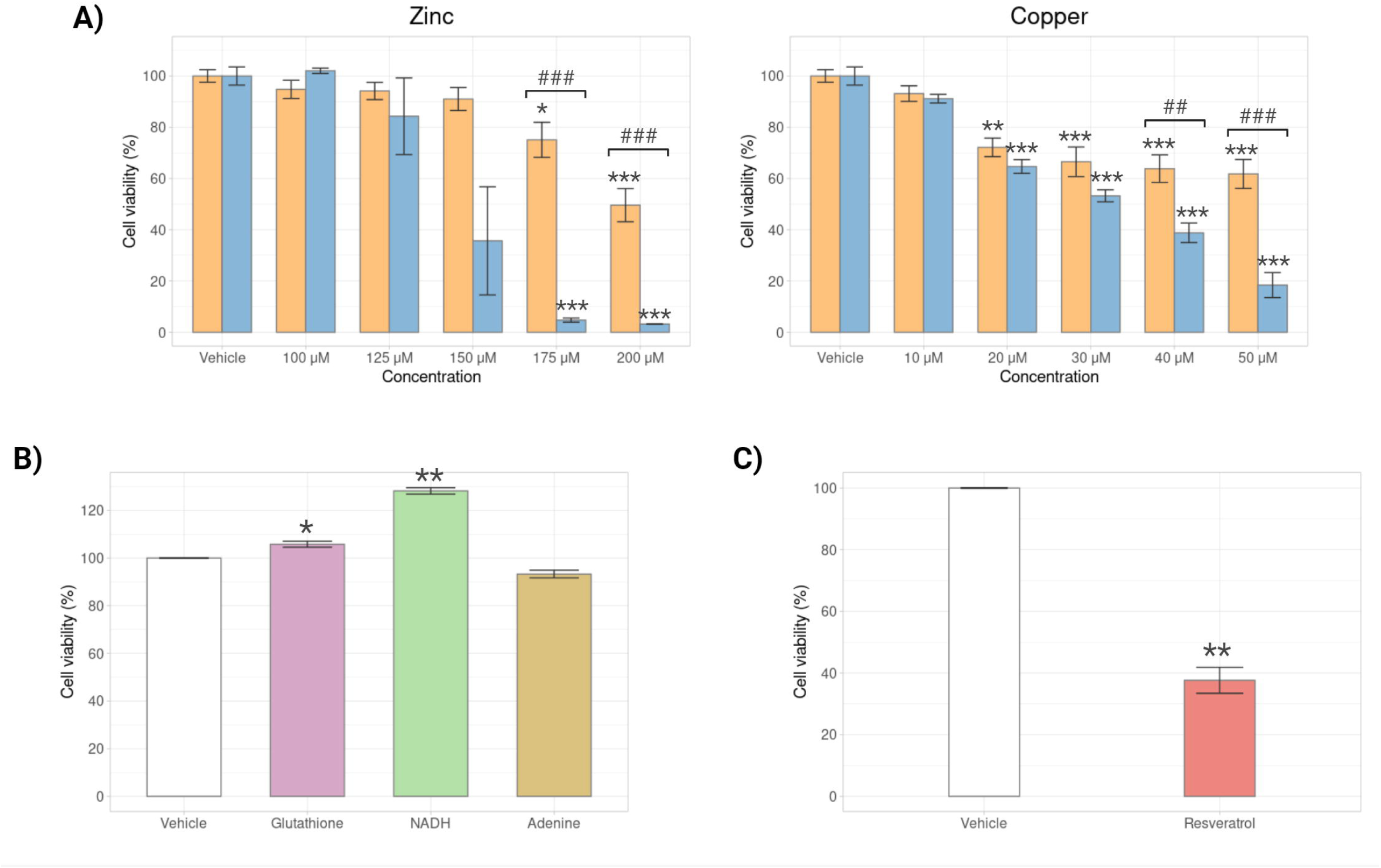

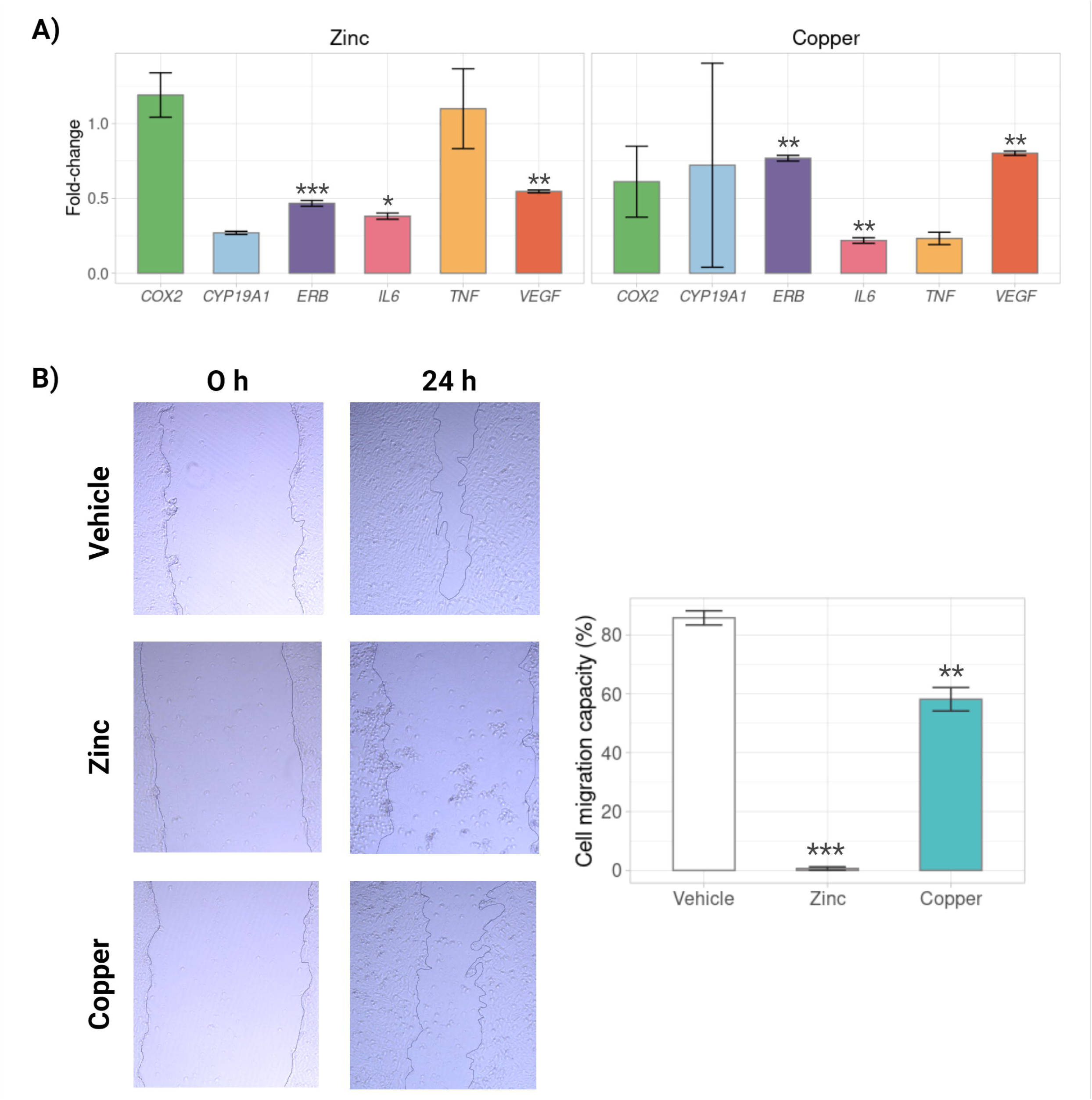

