## Supplemental Material for "Systems Pharmacology Model Predicts Zinc and Copper Can Be Repurposed as Endometriosis Therapies"

**Supplemental information**

**Supplemental material and methods**

**Disease gene selection**

Disease genes were prioritized from genomic sample repositories as follows. The raw data from microarray experiments of intrapatient ectopic and eutopic tissue from endometriosis women and endometrial tissue from healthy women, obtained from the two systematic reviews in GEO, were pre-processed independently using the Robust Multichip Average method included in the *limma* R-package (v3.46.0) (Ritchie *et al.*, 2015). Meanwhile, raw RNA-Seq data was processed by removing low-count genes (Counts Per Million (CPM) < 1.5) using the *edgeR* R-package (v3.32.1) (Robinson *et al.*, 2010) and *Voom* transformation included in *limma* R-package. Then, between-sample quantile normalization was achieved in all studies using *limma* R-package. An exploratory analysis was done using Principal Component Analysis (PCA) to detect possible outliers and batch effects. Batch effects related to the endometriosis stage (I-IV) and menstrual cycle phase at biopsy collection were corrected using linear models in *limma* R-package.

Once datasets were individually pre-processed, the guidelines provided in Tajti *et al.*, (2020) were followed to integrate them according to the study type: (i) infertility-related datasets included endometrial samples from healthy women and eutopic endometrial samples from patients with endometriosis; and (ii) progression-related datasets included eutopic and ectopic endometrial samples from patients with endometriosis. All datasets were integrated into a single dataset, retaining only the common genes among all datasets. Then, dataset variability was represented using PCA and linear models in *limma* (Ritchie *et al.*, 2015) were used to remove batch effects associated with each experiment. Next, a differential expression analyses (DEA) comparing each individual dataset was done independently in each condition using *limma* R-package to identify those genes still affected by the batch differences between individual dataset. Those genes with a false discovery rate (FDR) < 0.05 were excluded from subsequent analyses.

**Scale-free analysis**

For inferring network molecular properties, the scale free criterium is required. Scale-free analysis was carried out with the Network Analyzer plugin of Cytoscape v.3.8.2. This software analyzes whether the degree distribution of the nodes from a network follows a power law and plots the distribution in a log-log graph. Scale-free networks are characterized by having a few nodes with very high degree (hubs) and many nodes with low degree, implying that the network lacks a characteristic scale for the number of connections, as commonly observed in biological networks.

**Proximity analysis**

One of the properties for testing the drug efficacy is the molecular proximity to the target. For each set of gene pairs, we assess the network-based distance *d* between them by computing the average shortest path length, and compare it to the expected distance *drandom*, obtained by randomly selecting sets of genes (proteins) of the same set size within the interactome. The average shortest path length is a fundamental concept in network topology, defined as the average number of steps along the shortest paths between all pairs of nodes in the network. It serves as a measure of efficiency with which information or matter can be transmitted across the network. Then, the statistical significance of each distance (*d*) between each set of gene pairs was assessed by calculating a one-tailed z-test ($z=\frac{d-\mu}{\sigma}$,*d* = distance; *μ* = average of the group; *σ* = standard deviation), which reflects a statistical measure that indicates how far or close a value is from the mean of a group of values (a z-score < -1.645 corresponds to a p-value < 0.05 in a one-sided test). This proximity analysis was conducted to evaluate the distance between genes related to endometriosis progression and infertility, in order to assess their potential biological relationship. Additionally, the analysis was used to measure the proximity between drug targets and disease-associated genes within the human interactome, aiming to distinguish between palliative and effective drugs (see the Materials and Methods section for details). This analysis was conducted using the iGraph package v.0.11.3 (Ju *et al.*, 2016) within the Python programming language environment v.3.2.2 (Rossum and Drake, 2010) (Supplemental Figure 3).
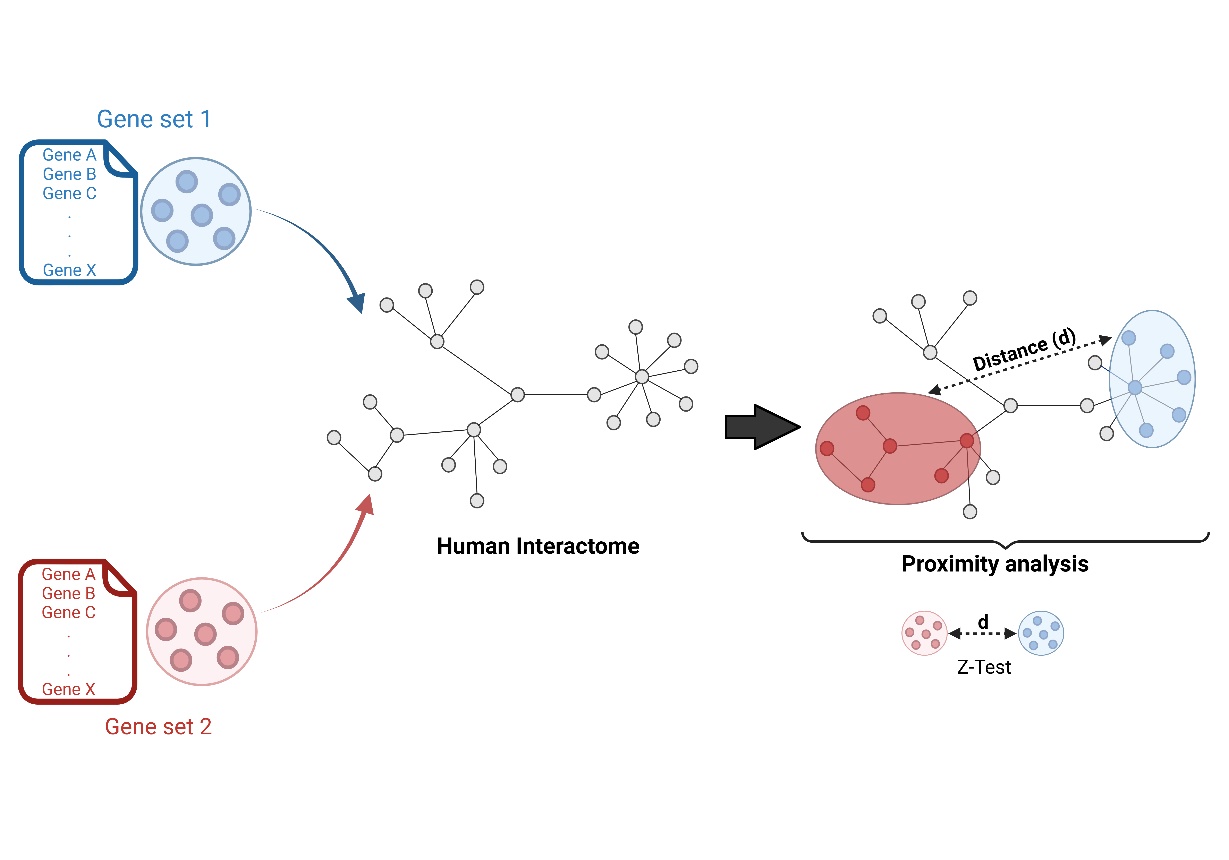
**Supplemental Figure 3:** Overview of proximity analysis. Different gene sets compared were mapped in the human interactoma. Next, the shortest distance between each set of genes were measured by calculating a one-tailed z-test.

**Drug prioritization based on network parameters**

The genes with degree and betweenness values higher than the relative maximum for their respective distributions were highlighted as the most influential in endometriosis pathophysiology, and drugs targeting them were prioritized. Next, drugs targeting a high number of disease genes (higher than the relative maximum) in the EDN network were labelled as promiscuous drugs. Drugs targeting high influence nodes (high degree and betweenness) were intersected with the most promiscuous drugs (higher than the relative maximum) to prioritize candidates to repurpose for endometriosis (Supplemental Figure 4).

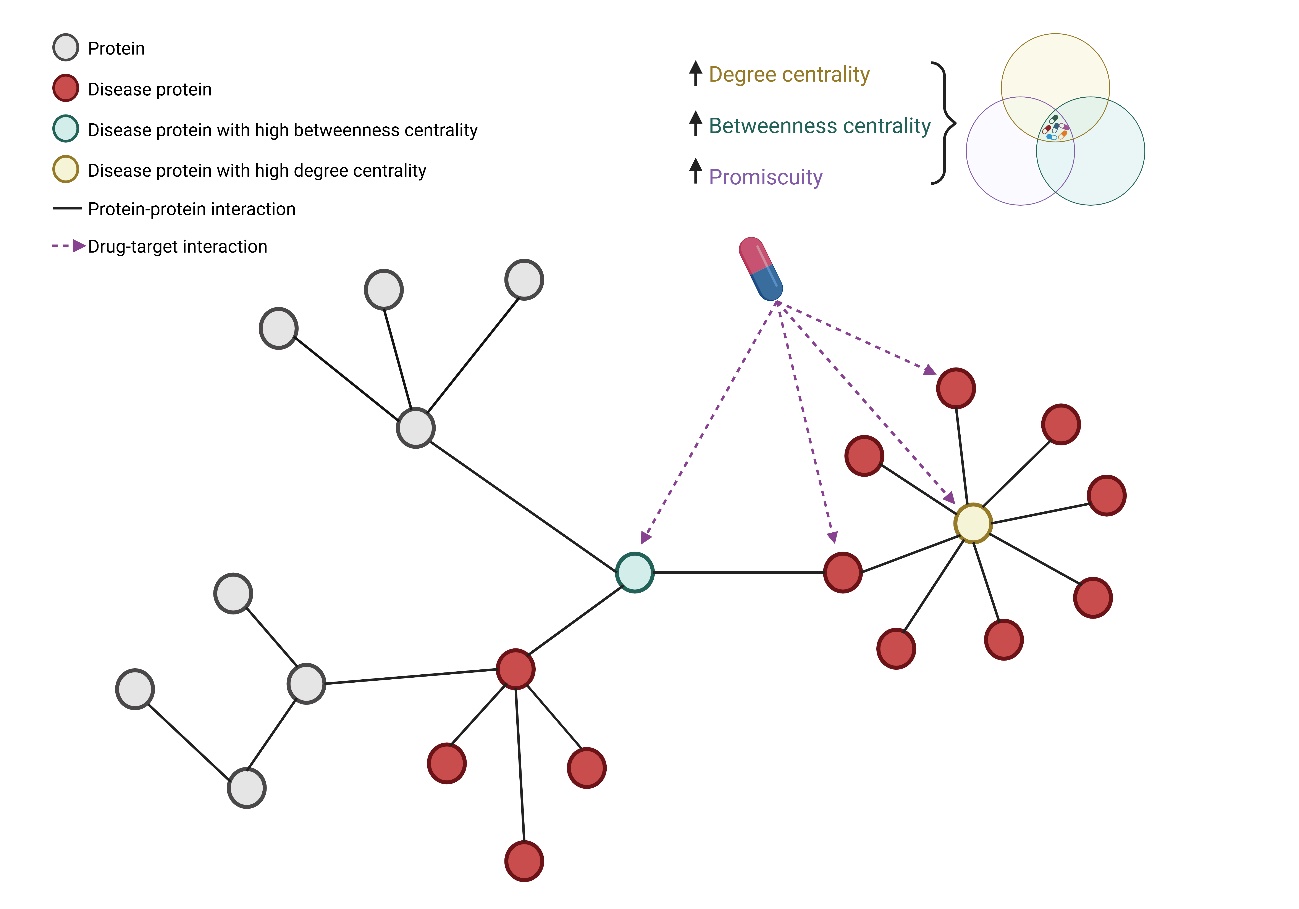

**Supplemental Figure 4:** Overview of drug prioritization based on network parameters. Circles represent proteins from the interactoma. Red circles represent proteins from the endometriosis disease network. Yellow circles are proteins with high degree centrality. Green circles represent proteins with high betweenness centrality. Purple dotted arrows represent proteins targeted by drugs,. Drugs prioritized were those targeting disease proteins with degree centrality, betweenness centrality and promiscuity (the number of targeted disease proteins) values higher than the relative maximum for their respective distributions.

**Supplemental results**

**Endometriosis Infertility-related dataset**

Three infertility-related datasets were retrieved from the systematic search in the Gene Expression Omnibus (GEO) database (**Supplemental Table 4**).

| **GEO identifier** | **Healthy biopsies (n)** | **Eutopic biopsies (n)** | **Stage** | **Endometrial phase** | **Reference** |
| --- | --- | --- | --- | --- | --- |
| GSE120103 | 18 | 16 | IV | N/A | (Bhat *et al.*, 2019) |
| GSE25628 | 6 | 8 | II-IV | PE | (Crispi *et al.*, 2013) |
| GSE6364 | 16 | 21 | Severe | PE, ESE & MSE | (Burney *et al.*, 2007) |

**Supplemental Table 4:** Infertility-related datasets included. GEO, Gene Expression Omnibus; N/A, Not available; PE, proliferative endometrium; ESE, early secretory endometrium; MSE, Mid-secretory endometrium.

Two outlier samples (GSM3393526 and GSM3393524) were detected and removed from dataset GSE120103. An endometrial phase batch effect was detected and removed in dataset GSE6364. The integration results for these datasets are represented in **Supplemental Figure 5**. A total of 83 samples (40 from healthy endometrium and 43 from eutopic endometrium) and 11,234 common genes were considered in the integrated dataset. After removing 2,993 genes with residual technical batch effects related to the experiment, the final infertility-related dataset included 83 samples and 8,241 genes.

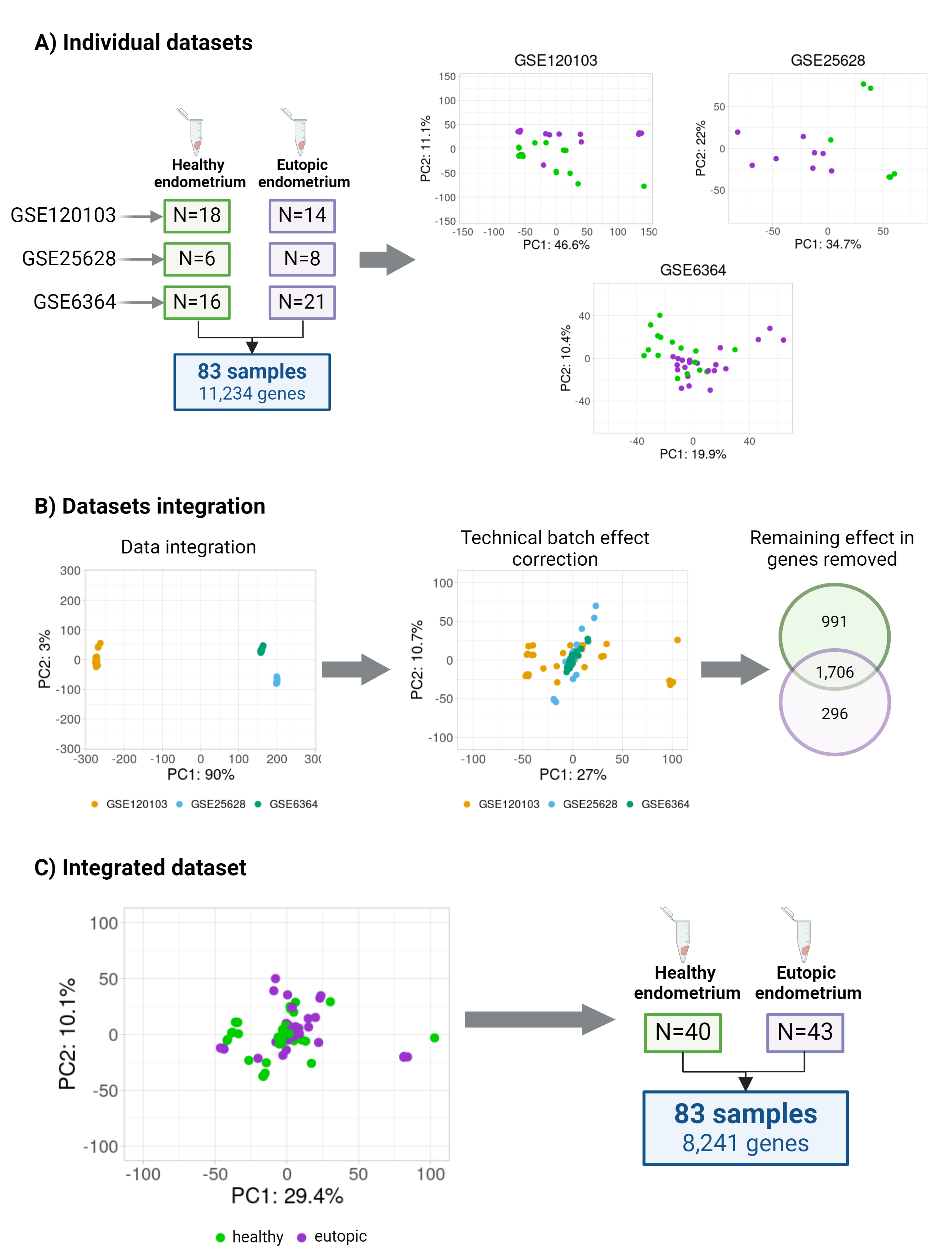

**Supplemental Figure 5: Endometriosis Infertility-related datasets integration. (A) Individual datasets.** Number of healthy (green) and eutopic (purple) endometrial tissue samples included in each dataset along with Principal Component Analysis (PCA) plots showing the transcriptomic behaviour of each dataset. **(B) Datasets integration.**PCA plots showing the transcriptomic behaviour of the integrated dataset before and after technical batch effect correction. In addition, Venn diagram showing the number of healthy (green) and eutopic (purple) genes with residual technical batch effects that were excluded from subsequent analyses. **(C) Integrated dataset.** PCA plot showing the transcriptomic behaviour in the final integrated dataset and final number of samples and genes considered for the subsequent analyses. PC1, first principal component; PC2, second principal component.

**Endometriosis Progression-related dataset**

Four progression-related datasets were retrieved from the systematic search in Gene Expression Omnibus (GEO) database (**Supplemental Table 5**).

| **GEO identifier** | **Eutopic biopsies (n)** | **Ectopic biopsies (n)** | **Stage** | **Endometrial phase** | **Reference** |
| --- | --- | --- | --- | --- | --- |
| GSE105764 | 8 | 8 | III-IV | SE | (Zhao *et al.*, 2018) |
| GSE11691 | 8 | 9 | II-IV | PE | (Hull *et al.*, 2008) |
| GSE25628 | 8 | 7 | II-IV | PE | (Crispi *et al.*, 2013) |
| GSE7305 | 10 | 10 | N/A | PE & SE | (Hever *et al.*, 2007) |

**Supplemental Table 5:** **Endometriosis Progression-related datasets included.** GEO = Gene Expression Omnibus; No. = Number of.; Ref. = Reference; N/A = Not available; PE = Proliferative endometrium; SE = Secretory endometrium.

Outlier samples were removed from dataset GSE11691 (GSM296876 and GSM296877) and dataset GSE25628 (GSM629733 and GSM629719). The integration results for these datasets are presented in **Supplemental Figure 6**. A total of 64 samples (32 from eutopic endometrium and 32 from ectopic endometrium) and 10,467 common genes were considered in the initial integrated dataset. After removing 2,612 genes with residual technical batch effects, the progression-related dataset included 64 samples and 7,855 genes.

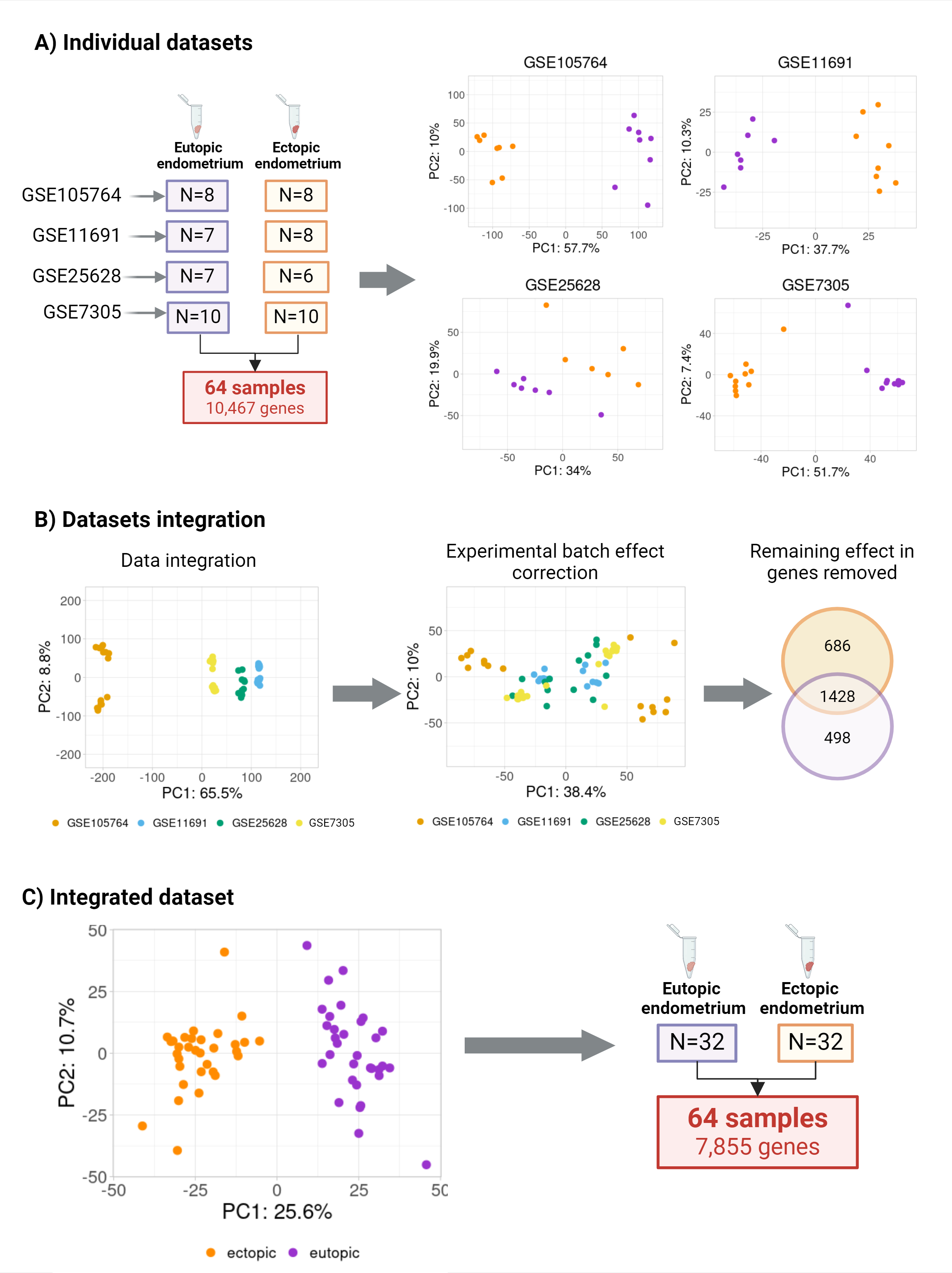

**Supplemental Figure 6: Progression-related datasets integration. (A) Individual datasets.** Number of ectopic (orange) and eutopic (purple) endometrial tissue samples included in each dataset along with Principal Component Analysis (PCA) plots showing the transcriptomic behaviour of each dataset. **(B) Datasets integration.** PCA plots showing the transcriptomic behaviour of the integrated dataset before and after technical batch effect correction. In addition, Venn diagram showing the number of ectopic (orange) and eutopic (purple) genes with residual technical batch effects that were excluded from subsequent analyses. **(C) Integrated dataset.** PCA plot showing the transcriptomic behaviour in the final integrated dataset and final number of samples and genes considered for the subsequent analyses. PC1, first principal component; PC2, second principal component.

**Endometriosis disease network**The EDN was constructed using all genes related to endometriosis infertility and endometriosis progression, encompassing 6,930 direct physical interactions between 2,567 endometriosis-related genes (2,151 exclusively related to progression, 250 exclusively related to infertility, and 166 related to both) **(Supplemental Table 3)**. This network followed a scale-free structure (R² = 0.911), indicating it is a valid model to mathematically infer endometriosis disease molecular mechanisms. The EDN was subsequently used to prioritize approved drugs targeting endometriosis-related genes.
