## Supplemental Table 1 for "Systems Pharmacology Model Predicts Zinc and Copper Can Be Repurposed as Endometriosis Therapies"

**Supplemental table 1.** **Primers used for RT-qPCR.**

| **Gene name** | **Forward** | **Reverse** |
| --- | --- | --- |
| ACTB | 5’-CGTACCACTGGCATCGTGAT-3’ | 5’-GTGTTGGCGTACAGGTCTTTG-3’ |
| CYP19A1 | 5’-GACGCAGGATTTCCACAGAAGAG-3’ | 5’-ATGGTGTCAGGAGCTGCGATCA-3’ |
| ERB | 5’-GATCGCTAGAACACACCTTAC-3’ | 5’-CGACCAGACTCCATAGTGATA-3’ |
| IL-6 | 5’-GAGAAAGGAGACATGTAACAAGAGT-3’ | 5’-GCGCAGAATGAGATGAGTTGT-3’ |
| VEGF | 5’-TCTTCAAGCCATCCTGTGTG-3’ | 5’- GGTGAGGTTTGATCCGCATA-3’ |
| TNF | 5’-ACTTTGGAGTGATCGGCCC-3’ | 5’-ATTGGCCAGGAGGGCATTG-3’ |
| COX2 | 5’-TTCAAATGAGATTGTGGGAAAATTGCT-3’ | 5’-AGATCATCTCTGCCTGAGTATCTT-3’ |
