## Supplemental Table 2 for "Systems Pharmacology Model Predicts Zinc and Copper Can Be Repurposed as Endometriosis Therapies"

**Supplemental Table 2. Prioritized genes**

|  | **Progression** | | **Infertility** | | **Network parameters** | |
| --- | --- | --- | --- | --- | --- | --- |
| **GeneID** | **Fold-Change** | **FDR** | **Fold-Change** | **FDR** | **Betweenness centrality** | **Degree** |
| A2M | 2.3293 | **1.96E-13** | -1.1961 | 0.4862 | 0.0001 | 2 |
| AAGAB | -1.3167 | **2.09E-05** | NA | NA | 0.0000 | 2 |
| AAK1 | 1.1916 | **0.0040** | -1.0100 | 0.9183 | 0.0000 | 1 |
| AAMDC | 1.5652 | **1.06E-11** | NA | NA | 0.0000 | 1 |
| AATF | 1.1069 | **0.0393** | -1.0892 | 0.4191 | 0.0001 | 4 |
| ABCA1 | 1.3210 | **0.0262** | NA | NA | **0.6667** | 2 |
| ABCB1 | 1.5599 | **1.46E-05** | -1.0431 | 0.7404 | 0.0000 | 1 |
| ABCD1 | 1.1745 | **0.0414** | -1.1821 | 0.0887 | 0.0000 | 2 |
| ABCD3 | -1.4181 | **3.06E-05** | NA | NA | 0.0000 | 2 |
| ABCE1 | -1.1794 | **0.0464** | 1.0499 | 0.7059 | 0.0000 | 1 |
| ABCF3 | -1.1744 | **0.0013** | -1.0594 | 0.6560 | 0.0000 | 3 |
| ABHD5 | -1.4541 | **0.0016** | NA | NA | 0.0001 | 3 |
| ABHD6 | 1.4043 | **8.60E-07** | 1.0931 | 0.3172 | 0.0000 | 1 |
| ABI2 | -1.2326 | **5.86E-06** | 1.0500 | 0.5600 | **0.0217** | **54** |
| ABL2 | 1.2354 | **0.0096** | -1.0691 | 0.3773 | 0.0006 | 6 |
| ABLIM3 | 1.3767 | **0.0047** | -1.2408 | 0.1471 | 0.0008 | 4 |
| ACACB | 2.7076 | **7.15E-18** | -1.0310 | 0.7107 | 0.0000 | 1 |
| ACADVL | -1.2166 | **0.0002** | -1.1242 | 0.1378 | 0.0000 | 2 |
| ACAP1 | 1.1313 | 0.2248 | -1.2347 | **0.0268** | 0.0008 | 2 |
| ACAT1 | 1.2280 | **0.0032** | -1.0270 | 0.7803 | 0.0000 | 1 |
| ACAT2 | -1.1951 | **0.0175** | -1.0760 | 0.5231 | 0.0000 | 1 |
| ACBD4 | 1.2098 | **0.0005** | 1.0942 | 0.6058 | 0.0000 | 1 |
| ACD | 1.2042 | **0.0065** | -1.0853 | 0.4541 | 0.0000 | 1 |
| ACE | -1.8318 | **1.07E-07** | 1.0415 | 0.8340 | 0.0000 | 1 |
| ACIN1 | -1.2382 | **0.0002** | NA | NA | 0.0000 | 2 |
| ACKR3 | 1.4582 | **0.0005** | NA | NA | 0.0000 | 1 |
| ACO2 | -1.4000 | **7.40E-06** | NA | NA | 0.0000 | 1 |
| ACOT8 | -1.2143 | **0.0018** | -1.0007 | 0.9960 | 0.0000 | 1 |
| ACOX2 | 2.4112 | **3.89E-13** | NA | NA | 0.0000 | 1 |
| ACP1 | -1.3658 | **7.55E-07** | 1.0142 | 0.8979 | 0.0002 | 2 |
| ACSF2 | 1.3023 | **0.0005** | 1.0094 | 0.9395 | 0.0001 | 3 |
| ACTL6A | -1.4352 | **2.85E-05** | 1.2458 | 0.2172 | 0.0000 | 1 |
| ACTN1 | 1.5416 | **1.24E-05** | NA | NA | **0.0026** | 11 |
| ACTR2 | -1.2277 | **0.0186** | 1.5285 | **0.0185** | 0.0000 | 1 |
| ACVR1 | 1.2281 | **0.0202** | -1.0422 | 0.7594 | 0.0000 | 1 |
| ADAM19 | 1.4589 | **0.0004** | NA | NA | 0.0000 | 1 |
| ADAMTS12 | -1.3956 | **0.0003** | 1.0788 | 0.2456 | 0.0001 | 2 |
| ADAMTS13 | -1.2934 | **0.0003** | -1.2224 | 0.3084 | 0.0000 | 1 |
| ADAMTS3 | 2.3156 | **4.38E-08** | -1.0385 | 0.7524 | 0.0000 | 1 |
| ADAMTSL4 | -1.2324 | **0.0309** | -1.2080 | 0.0767 | **0.0156** | **49** |
| ADAP1 | -1.0708 | 0.4513 | -1.2101 | **0.0161** | 0.0003 | 3 |
| ADARB1 | 1.4408 | **1.93E-05** | -1.2392 | **0.0161** | 0.0005 | 5 |
| ADCYAP1R1 | -2.1629 | **0.0007** | 1.1745 | 0.1934 | 0.0000 | 1 |
| ADD1 | 1.3163 | **1.74E-06** | -1.0752 | 0.4929 | 0.0008 | 2 |
| ADGRA2 | 1.3329 | **0.0046** | NA | NA | 0.0000 | 1 |
| ADGRA3 | -1.8734 | **5.60E-10** | NA | NA | 0.0000 | 1 |
| ADGRB1 | 1.3559 | **0.0238** | NA | NA | 0.0000 | 1 |
| ADGRF5 | 3.0368 | **1.40E-22** | NA | NA | 0.0000 | 1 |
| ADGRG3 | -1.3150 | **0.0338** | NA | NA | 0.0000 | 2 |
| ADIPOR1 | -1.0535 | 0.3863 | 1.1693 | **0.0365** | 0.0000 | 2 |
| ADNP2 | 1.0218 | 0.7095 | 1.2412 | **0.0176** | 0.0000 | 1 |
| AEN | -1.5466 | **4.75E-05** | NA | NA | 0.0005 | 9 |
| AFF1 | 1.1689 | **0.0272** | 1.1912 | **0.0278** | 0.0010 | 4 |
| AFTPH | -1.2148 | **0.0012** | NA | NA | 0.0000 | 1 |
| AGFG1 | -1.1846 | **0.0318** | 1.0803 | 0.3450 | 0.0002 | 2 |
| AGO1 | 1.2931 | **2.01E-05** | NA | NA | 0.0016 | 3 |
| AGO2 | -1.4332 | **2.98E-06** | NA | NA | 0.0000 | 2 |
| AGPAT3 | -1.1687 | **0.0035** | 1.0153 | 0.8545 | 0.0007 | 5 |
| AGPAT4 | 1.5613 | **7.08E-08** | -1.1772 | 0.0580 | **0.0029** | 7 |
| AGPAT5 | -2.2882 | **2.01E-09** | NA | NA | 0.0000 | 2 |
| AGPS | -1.1472 | **0.0311** | 1.0500 | 0.5712 | 0.0000 | 1 |
| AGRN | -1.5824 | **1.28E-07** | 1.0707 | 0.5026 | 0.0000 | 2 |
| AGXT | NA | NA | -1.2196 | **0.0481** | 0.0001 | 6 |
| AHCTF1 | 1.2390 | **0.0040** | 1.3121 | **0.0161** | **0.0032** | 4 |
| AHCYL1 | -1.2973 | **2.37E-05** | 1.1622 | 0.1140 | 0.0001 | 4 |
| AHR | 2.1987 | **1.98E-11** | NA | NA | 0.0016 | 2 |
| AHSA1 | -1.3053 | **6.15E-06** | NA | NA | 0.0000 | 2 |
| AIFM1 | -2.2963 | **1.70E-13** | NA | NA | 0.0000 | 2 |
| AIMP2 | -1.5919 | **4.81E-12** | -1.0064 | 0.9664 | 0.0017 | 10 |
| AIP | 1.3883 | **8.29E-06** | NA | NA | 0.0008 | 2 |
| AIRE | NA | NA | -1.2761 | **0.0088** | 0.0004 | 5 |
| AKAP1 | -1.1953 | **0.0012** | 1.0932 | 0.2888 | 0.0008 | 3 |
| AKAP7 | -1.3978 | **1.12E-06** | -1.0199 | 0.8795 | 0.0008 | 4 |
| AKT3 | 2.3774 | **4.13E-12** | 1.0412 | 0.7303 | 0.0000 | 2 |
| AKTIP | 1.3009 | **0.0001** | 1.1356 | 0.1391 | 0.0000 | 2 |
| ALAS1 | -1.3633 | **0.0002** | NA | NA | 0.0014 | 9 |
| ALCAM | -2.0453 | **1.30E-07** | NA | NA | 0.0000 | 1 |
| ALG13 | -1.1627 | **0.0340** | -1.0552 | 0.6861 | 0.0000 | 2 |
| ALG3 | -1.2756 | **0.0008** | NA | NA | 0.0000 | 1 |
| ALG8 | -1.4203 | **1.14E-09** | 1.0053 | 0.9511 | 0.0000 | 5 |
| ALK | NA | NA | -1.1711 | **0.0097** | 0.0000 | 1 |
| ALOX12 | -1.1708 | **0.0272** | 1.1040 | 0.4754 | 0.0000 | 1 |
| ALOX5 | 1.5647 | **0.0020** | -1.1696 | 0.3449 | 0.0000 | 2 |
| AMBRA1 | 1.0709 | 0.2458 | 1.2001 | **0.0184** | 0.0002 | 3 |
| AMD1 | -1.6422 | **4.17E-05** | NA | NA | 0.0000 | 1 |
| AMFR | -1.2537 | **0.0089** | NA | NA | 0.0022 | 6 |
| AMOTL2 | 1.6516 | **2.16E-05** | -1.1094 | 0.5309 | **0.0062** | **28** |
| AMPD2 | 1.0864 | 0.2722 | -1.1737 | **0.0476** | 0.0000 | 1 |
| ANGPT1 | 4.2673 | **7.23E-16** | NA | NA | 0.0000 | 2 |
| ANGPTL4 | 1.5394 | **0.0079** | -1.2475 | 0.1932 | 0.0000 | 1 |
| ANK2 | 3.3651 | **2.96E-10** | -1.0053 | 0.9785 | 0.0000 | 1 |
| ANK3 | -2.0069 | **5.64E-07** | NA | NA | 0.0000 | 1 |
| ANKMY1 | -1.3058 | **0.0001** | 1.0218 | 0.7923 | 0.0000 | 1 |
| ANKRD46 | 1.5218 | **1.65E-06** | 1.1187 | 0.4889 | 0.0019 | **14** |
| ANOS1 | 3.7746 | **4.15E-10** | NA | NA | 0.0000 | 1 |
| ANP32A | -1.4663 | **1.09E-06** | -1.0548 | 0.5183 | 0.0001 | 2 |
| ANXA2 | -1.6602 | **1.65E-05** | -1.0096 | 0.9272 | 0.0017 | 6 |
| AOC1 | -2.2262 | **0.0015** | NA | NA | 0.0001 | 4 |
| AOC3 | 9.5206 | **3.00E-26** | -1.3731 | 0.0892 | 0.0001 | 3 |
| AP1G1 | -1.6419 | **4.04E-13** | 1.1337 | 0.1473 | 0.0007 | 3 |
| AP2A2 | -1.2096 | **0.0011** | -1.0283 | 0.7524 | 0.0004 | 2 |
| AP2B1 | -1.1394 | **0.0057** | 1.0166 | 0.8754 | **0.0037** | 13 |
| AP3B1 | -1.2398 | **3.38E-05** | 1.0328 | 0.6582 | 0.0008 | 2 |
| AP4M1 | -1.4171 | **9.14E-06** | NA | NA | **0.6667** | 2 |
| APBA2 | -1.2271 | **0.0048** | -1.0062 | 0.9627 | 0.0000 | 1 |
| APBB1 | 1.2700 | **0.0003** | -1.2179 | **0.0452** | 0.0021 | 4 |
| APBB2 | 1.2463 | **0.0019** | NA | NA | 0.0000 | 1 |
| APC | 1.1803 | **0.0211** | NA | NA | 0.0019 | 8 |
| APEH | -1.3715 | **1.46E-05** | NA | NA | 0.0000 | 1 |
| APEX1 | -1.3389 | **3.14E-07** | 1.0579 | 0.5699 | 0.0001 | 2 |
| APEX2 | -1.1917 | **0.0029** | NA | NA | 0.0001 | 3 |
| APOA1 | 4.1276 | **9.80E-08** | NA | NA | 0.0000 | 1 |
| APOL1 | -1.5192 | **0.0045** | -1.2183 | 0.1598 | 0.0000 | 1 |
| APOL3 | 3.1325 | **8.48E-16** | -1.2577 | 0.0913 | 0.0002 | 6 |
| APP | 1.6680 | **9.29E-08** | 1.5824 | **0.0078** | **0.0133** | **25** |
| APPL1 | 1.0892 | 0.3318 | 1.4841 | **0.0160** | 0.0020 | 8 |
| APRT | -1.3062 | **3.04E-05** | NA | NA | 0.0002 | 2 |
| APTX | -1.2507 | **1.10E-05** | 1.0257 | 0.7577 | 0.0000 | 3 |
| AQP1 | 5.6882 | **1.12E-24** | NA | NA | **0.0213** | **28** |
| ARCN1 | -1.2211 | **0.0012** | -1.1326 | 0.1048 | 0.0000 | 1 |
| AREG | 1.9176 | **0.0172** | -1.1101 | 0.5577 | 0.0000 | 1 |
| AREL1 | -1.5662 | **9.53E-10** | NA | NA | 0.0000 | 1 |
| ARF1 | -1.3061 | **5.60E-09** | NA | NA | 0.0021 | 3 |
| ARF4 | -1.2239 | **0.0022** | -1.1000 | 0.1636 | 0.0000 | 1 |
| ARF6 | -1.1667 | **0.0138** | 1.0702 | 0.4834 | 0.0000 | 1 |
| ARFGAP3 | -1.8745 | **2.55E-11** | 1.0448 | 0.7286 | 0.0000 | 1 |
| ARFGEF1 | -1.4113 | **7.63E-06** | NA | NA | 0.0000 | 1 |
| ARFIP1 | -1.3000 | **1.80E-06** | NA | NA | 0.0004 | 6 |
| ARFIP2 | -1.5158 | **2.75E-11** | -1.1630 | 0.0719 | **0.0064** | 13 |
| ARGLU1 | 1.0767 | 0.4265 | 1.6461 | **0.0161** | 0.0000 | 1 |
| ARHGAP15 | 2.1656 | **6.23E-09** | 1.0323 | 0.7524 | 0.0000 | 1 |
| ARHGAP32 | -1.2309 | **0.0033** | 1.2555 | **0.0433** | 0.0011 | 6 |
| ARHGAP44 | 1.1857 | **0.0116** | -1.0480 | 0.6100 | 0.0000 | 1 |
| ARHGAP45 | 1.5528 | **0.0001** | NA | NA | 0.0000 | 3 |
| ARHGDIA | -1.2620 | **0.0102** | 1.2839 | **0.0381** | 0.0001 | 2 |
| ARHGDIB | 2.1205 | **7.94E-09** | -1.2107 | 0.0540 | 0.0000 | 1 |
| ARHGDIG | 1.0439 | 0.6380 | -1.3342 | **0.0077** | 0.0000 | 2 |
| ARHGEF1 | 1.0432 | 0.5330 | -1.2622 | **0.0013** | 0.0000 | 1 |
| ARHGEF15 | 1.1980 | **0.0340** | -1.3444 | **0.0476** | 0.0001 | 2 |
| ARHGEF16 | NA | NA | 1.2300 | **0.0133** | 0.0000 | 4 |
| ARHGEF3 | 1.5729 | **1.34E-06** | 1.1271 | 0.2467 | 0.0001 | 3 |
| ARHGEF9 | 1.4600 | **2.51E-06** | 1.0413 | 0.7107 | 0.0008 | 2 |
| ARID1A | -1.2003 | **0.0004** | 1.0516 | 0.6333 | 0.0000 | 1 |
| ARID3A | -1.5038 | **9.19E-07** | -1.3117 | 0.2600 | 0.0001 | 4 |
| ARID4B | 1.1716 | **0.0193** | NA | NA | 0.0005 | 4 |
| ARIH1 | -1.1122 | 0.0752 | 1.2335 | **0.0280** | 0.0000 | 1 |
| ARIH2 | -1.0864 | **0.0346** | -1.0001 | 0.9988 | 0.0001 | 3 |
| ARL2 | 1.2328 | **0.0007** | NA | NA | **0.6667** | 2 |
| ARL2BP | -1.1145 | **0.0181** | -1.0314 | 0.8536 | **0.6667** | 2 |
| ARL5A | 1.2156 | **0.0164** | NA | NA | 0.0000 | 1 |
| ARL6IP1 | -1.3333 | **0.0003** | NA | NA | **0.0110** | **17** |
| ARL6IP5 | 1.4154 | **5.55E-07** | NA | NA | 0.0000 | 1 |
| ARL8B | 1.0670 | 0.3380 | 1.1940 | **0.0119** | 0.0000 | 1 |
| ARMC7 | 1.1649 | **0.0019** | 1.0830 | 0.3953 | **0.0030** | **19** |
| ARMC9 | 1.2422 | **0.0323** | -1.0293 | 0.8426 | 0.0000 | 1 |
| ARMCX1 | 1.8775 | **7.08E-08** | -1.1392 | 0.2942 | 0.0000 | 3 |
| ARMT1 | -1.8361 | **5.69E-07** | NA | NA | 0.0000 | 1 |
| ARNT2 | 1.2723 | **0.0159** | -1.0119 | 0.9277 | 0.0005 | 6 |
| ARNTL2 | -1.2292 | **0.0326** | 1.1515 | 0.1267 | 0.0000 | 1 |
| ARPC3 | -1.4032 | **0.0003** | NA | NA | 0.0019 | 5 |
| ARPIN | 1.1593 | **0.0225** | NA | NA | 0.0000 | 2 |
| ARRB1 | 1.5742 | **9.67E-09** | -1.1449 | 0.1971 | 0.0003 | 5 |
| ASAP2 | 1.0964 | 0.3488 | 1.3590 | **0.0028** | 0.0000 | 1 |
| ASAP3 | NA | NA | -1.4035 | **0.0010** | 0.0011 | 5 |
| ASB6 | -1.1088 | **0.0393** | NA | NA | 0.0022 | 6 |
| ASB8 | 1.3004 | **3.33E-06** | 1.0813 | 0.3126 | 0.0000 | 1 |
| ASF1B | -2.5784 | **2.39E-08** | 1.0775 | 0.7074 | 0.0000 | 1 |
| ASL | -2.3363 | **1.19E-12** | 1.2501 | 0.1134 | 0.0000 | 2 |
| ASNA1 | -1.2671 | **0.0003** | NA | NA | **0.0028** | 11 |
| ASPH | 1.5537 | **1.22E-06** | 1.2107 | 0.0605 | **0.0045** | 13 |
| ASPM | -1.9290 | **0.0006** | 1.3772 | 0.1553 | 0.0000 | 1 |
| ASPSCR1 | -1.3502 | **4.93E-05** | NA | NA | 0.0010 | 5 |
| ATAD3A | -1.3429 | **3.69E-07** | 1.0548 | 0.6860 | 0.0000 | 1 |
| ATF1 | 1.2381 | **0.0232** | NA | NA | 0.0000 | 1 |
| ATF7IP2 | 1.3514 | **0.0019** | 1.0327 | 0.7436 | 0.0000 | 1 |
| ATG101 | -1.1935 | **0.0031** | NA | NA | 0.0000 | 1 |
| ATG12 | 1.0863 | **0.0398** | -1.1007 | 0.2713 | 0.0008 | 2 |
| ATG4B | -1.3899 | **1.85E-09** | -1.1266 | 0.1178 | 0.0000 | 2 |
| ATN1 | 1.1684 | **0.0145** | -1.1329 | 0.5671 | **0.0030** | 13 |
| ATP13A1 | -1.2437 | **0.0008** | NA | NA | 0.0000 | 2 |
| ATP13A2 | -1.4412 | **3.82E-05** | NA | NA | **0.0107** | **19** |
| ATP1B3 | -1.4282 | **0.0003** | 1.0089 | 0.9331 | 0.0000 | 1 |
| ATP5F1A | -1.3877 | **1.35E-09** | NA | NA | 0.0005 | 3 |
| ATP5F1B | -1.3618 | **1.10E-08** | NA | NA | 0.0000 | 3 |
| ATP5F1C | -1.1679 | **0.0085** | NA | NA | 0.0000 | 1 |
| ATP5MPL | -1.2110 | **0.0004** | NA | NA | 0.0008 | 2 |
| ATP6V0B | -1.2110 | **0.0294** | 1.0478 | 0.7601 | 0.0004 | 3 |
| ATP6V0D1 | -1.2677 | **0.0003** | -1.1836 | 0.0816 | 0.0000 | 1 |
| ATP6V1B2 | 1.2281 | **0.0162** | NA | NA | 0.0000 | 1 |
| ATP6V1D | -1.0254 | 0.7779 | 1.4249 | **0.0008** | 0.0000 | 2 |
| ATP6V1G1 | -1.3782 | **0.0001** | 1.1774 | 0.1516 | 0.0004 | 6 |
| ATXN10 | -1.2895 | **1.55E-06** | 1.0689 | 0.5692 | 0.0000 | 1 |
| ATXN7 | -1.1913 | **0.0101** | NA | NA | 0.0022 | 7 |
| AURKA | -2.2036 | **4.86E-07** | 1.1744 | 0.3562 | 0.0000 | 1 |
| AURKB | -2.5481 | **4.99E-08** | 1.0828 | 0.6506 | 0.0006 | 7 |
| AVEN | 1.4688 | **3.38E-07** | -1.1083 | 0.1948 | 0.0000 | 1 |
| AVPI1 | 1.3439 | **0.0018** | -1.1844 | 0.1788 | 0.0004 | 6 |
| AXIN1 | -1.1304 | **0.0219** | NA | NA | **0.0081** | **20** |
| AXL | 1.2420 | **0.0032** | NA | NA | 0.0000 | 1 |
| B9D2 | -1.1954 | **0.0041** | NA | NA | 0.0000 | 2 |
| BABAM1 | -1.1321 | **0.0077** | NA | NA | 0.0002 | 2 |
| BACH2 | 1.7458 | **0.0001** | NA | NA | 0.0008 | 9 |
| BAG2 | 1.7635 | **0.0004** | -1.3368 | **0.0353** | 0.0000 | 1 |
| BAG3 | 1.2015 | **0.0209** | -1.0975 | 0.3641 | **0.0041** | **21** |
| BAG5 | -1.6197 | **5.78E-06** | 1.1767 | 0.3905 | 0.0012 | 5 |
| BAG6 | -1.1735 | **0.0019** | 1.0130 | 0.9102 | **0.0030** | 10 |
| BAK1 | -1.1890 | **0.0055** | 1.0238 | 0.8160 | 0.0002 | 3 |
| BAMBI | 2.1275 | **2.01E-07** | -1.4338 | **0.0102** | 0.0000 | 1 |
| BANF1 | -1.1562 | **0.0123** | -1.1616 | 0.1353 | 0.0000 | 2 |
| BARD1 | -1.7625 | **1.87E-06** | 1.2020 | 0.2208 | **0.0059** | 13 |
| BAZ1A | -1.5920 | **1.19E-06** | NA | NA | 0.0000 | 1 |
| BBOF1 | -1.8202 | **1.69E-07** | NA | NA | 0.0000 | 1 |
| BCAM | 1.3017 | **0.0001** | 1.1050 | 0.3351 | 0.0000 | 1 |
| BCAR3 | 1.2665 | **0.0169** | -1.0661 | 0.6497 | 0.0000 | 2 |
| BCKDHA | -1.2492 | **0.0004** | 1.0796 | 0.3356 | 0.0000 | 1 |
| BCKDHB | -1.3354 | **0.0002** | 1.1578 | 0.3066 | 0.0000 | 1 |
| BCL10 | -1.2599 | **0.0005** | NA | NA | 0.0001 | 3 |
| BCL2A1 | 1.9585 | **0.0065** | 1.1036 | 0.7158 | 0.0000 | 2 |
| BCL2L1 | -1.2843 | **0.0019** | 1.0182 | 0.8424 | **0.0132** | **18** |
| BCL2L2 | 1.4464 | **7.48E-14** | -1.0766 | 0.4385 | **0.0064** | **15** |
| BCL7A | -1.2472 | **0.0010** | NA | NA | 0.0000 | 1 |
| BCL9 | -1.2952 | **0.0015** | 1.1438 | 0.3321 | 0.0000 | 1 |
| BCORL1 | -1.2225 | **0.0029** | 1.1073 | 0.5148 | 0.0000 | 1 |
| BCR | -1.6527 | **5.15E-10** | NA | NA | 0.0001 | 3 |
| BDKRB2 | -2.4089 | **8.97E-07** | -1.2428 | 0.1449 | 0.0000 | 1 |
| BECN1 | -1.1917 | **0.0010** | -1.0466 | 0.6231 | 0.0023 | 7 |
| BET1 | -1.3592 | **0.0005** | 1.1449 | 0.2225 | 0.0005 | 12 |
| BEX3 | 1.1668 | **0.0419** | NA | NA | 0.0001 | 6 |
| BICD2 | 1.2246 | **0.0055** | -1.0203 | 0.8360 | 0.0013 | 12 |
| BICRAL | 1.3461 | **0.0009** | NA | NA | 0.0006 | 6 |
| BIK | -1.5188 | **0.0003** | 1.4571 | **0.0161** | **0.0049** | **27** |
| BIN1 | -1.1488 | **0.0484** | NA | NA | 0.0001 | 4 |
| BIN3 | -1.2334 | **0.0001** | NA | NA | 0.0011 | 4 |
| BIRC5 | -2.8279 | **2.40E-09** | NA | NA | 0.0008 | 7 |
| BLCAP | 1.2026 | **0.0020** | 1.1013 | 0.3194 | 0.0000 | 1 |
| BLM | -1.8424 | **4.48E-07** | 1.2106 | 0.1154 | 0.0024 | 6 |
| BLNK | -1.6170 | **0.0008** | 1.1479 | 0.3633 | 0.0000 | 2 |
| BMI1 | 1.3746 | **0.0004** | NA | NA | 0.0003 | 4 |
| BMPR2 | 1.2987 | **0.0001** | 1.1712 | 0.0973 | 0.0000 | 1 |
| BNIP1 | -1.2709 | **2.58E-05** | NA | NA | 0.0003 | 6 |
| BNIP2 | 1.3019 | **0.0006** | NA | NA | **0.0034** | 13 |
| BNIP3L | 1.2237 | **0.0211** | NA | NA | **0.0026** | 4 |
| BORCS6 | 1.1917 | **0.0053** | NA | NA | 0.0020 | 12 |
| BRAF | -1.3905 | **2.76E-07** | -1.0996 | 0.5776 | 0.0003 | 4 |
| BRD1 | -1.2250 | **0.0002** | 1.0250 | 0.8394 | 0.0004 | 3 |
| BRD2 | -1.1494 | **0.0058** | 1.1887 | 0.1163 | 0.0000 | 4 |
| BRD4 | 1.1387 | **0.0433** | NA | NA | 0.0017 | 9 |
| BRIX1 | -1.3606 | **0.0124** | 1.3207 | 0.0859 | 0.0000 | 1 |
| BSG | -1.2461 | **0.0029** | NA | NA | 0.0000 | 1 |
| BTBD3 | -2.8733 | **5.39E-12** | NA | NA | 0.0002 | 3 |
| BTG1 | 1.7426 | **6.82E-10** | 1.0228 | 0.8528 | 0.0023 | 5 |
| BTRC | -1.1385 | **0.0058** | -1.1493 | **0.0262** | 0.0001 | 3 |
| BUB3 | -1.2309 | **0.0001** | 1.1063 | 0.1861 | 0.0011 | 3 |
| BYSL | -1.8021 | **3.74E-09** | NA | NA | **0.0169** | **41** |
| C10orf88 | -1.2301 | **0.0079** | 1.1206 | 0.2038 | 0.0000 | 2 |
| C11orf49 | -1.2913 | **1.49E-06** | 1.1130 | 0.4354 | 0.0004 | 6 |
| C11orf68 | 1.3233 | **1.55E-06** | NA | NA | 0.0013 | 5 |
| C12orf49 | -1.3318 | **0.0002** | -1.0966 | 0.4559 | 0.0000 | 1 |
| C16orf58 | -1.1159 | **0.0416** | NA | NA | **0.0060** | 13 |
| C17orf75 | -1.2010 | **0.0450** | -1.0099 | 0.9286 | 0.0000 | 1 |
| C18orf25 | -1.1953 | **0.0002** | NA | NA | 0.0001 | 3 |
| C19orf66 | 1.2190 | **3.43E-05** | NA | NA | 0.0002 | 8 |
| C19orf73 | -1.1514 | **0.0464** | NA | NA | 0.0000 | 1 |
| C1orf105 | NA | NA | 1.1843 | **0.0447** | 0.0000 | 2 |
| C1orf109 | -1.1528 | 0.2061 | 1.4736 | **0.0069** | **0.0110** | **40** |
| C1QBP | -1.5125 | **4.80E-07** | 1.0611 | 0.6186 | 0.0000 | 1 |
| C1QTNF1 | 2.4548 | **2.34E-12** | NA | NA | 0.0000 | 2 |
| C1S | NA | NA | -1.5470 | **0.0010** | 0.0000 | 1 |
| C2 | 2.0213 | **4.14E-11** | -1.1262 | 0.2350 | 0.0001 | 4 |
| C20orf27 | -1.2819 | **0.0208** | -1.1142 | 0.5072 | 0.0000 | 1 |
| C3orf52 | -1.5311 | **0.0001** | NA | NA | **0.0050** | **17** |
| C6orf47 | 1.1355 | **0.0180** | NA | NA | 0.0000 | 1 |
| CA3 | 2.2511 | **4.32E-08** | -1.0359 | 0.6896 | 0.0000 | 1 |
| CA9 | NA | NA | 1.2166 | **0.0290** | 0.0006 | 2 |
| CAB39 | 1.3121 | **0.0001** | 1.1886 | 0.1555 | 0.0019 | 5 |
| CAB39L | 1.4057 | **0.0114** | -1.0493 | 0.6754 | 0.0000 | 1 |
| CACNA1S | NA | NA | -1.3048 | **0.0266** | 0.0000 | 2 |
| CACTIN | -1.1241 | **0.0417** | NA | NA | 0.0000 | 1 |
| CACYBP | -1.2396 | **0.0036** | NA | NA | 0.0012 | 3 |
| CADM1 | -1.4712 | **0.0207** | 1.2758 | 0.1121 | 0.0000 | 1 |
| CALCOCO1 | 1.5821 | **1.11E-08** | NA | NA | 0.0009 | 4 |
| CALCOCO2 | 1.2335 | **0.0007** | -1.1028 | 0.1836 | **0.0154** | **46** |
| CALM1 | 1.2240 | **0.0007** | 1.1804 | **0.0169** | **0.0035** | 9 |
| CALR | -1.3177 | **3.72E-05** | -1.0279 | 0.7942 | 0.0006 | 4 |
| CAMK2G | 1.5499 | **4.61E-09** | -1.0739 | 0.5074 | 0.0022 | 6 |
| CAMLG | 1.4969 | **8.40E-08** | -1.0177 | 0.8564 | 0.0010 | 8 |
| CAMSAP2 | 1.2313 | **0.0155** | 1.0217 | 0.8562 | 0.0000 | 1 |
| CAND2 | 1.8783 | **2.11E-06** | -1.1387 | 0.3363 | 0.0003 | 2 |
| CAP1 | -1.0009 | 0.9897 | 1.3764 | **0.0407** | 0.0000 | 1 |
| CAPN15 | -1.2396 | **0.0010** | NA | NA | 0.0002 | 2 |
| CAPN3 | 1.2324 | **0.0306** | -1.1891 | 0.1568 | **0.0042** | 13 |
| CAPNS1 | -1.4371 | **7.00E-08** | 1.1111 | 0.5498 | 0.0000 | 1 |
| CARD14 | 1.0609 | 0.5116 | -1.1906 | **0.0423** | 0.0000 | 2 |
| CARD9 | -1.2431 | **0.0416** | 1.0444 | 0.7216 | **0.0072** | **39** |
| CARM1 | -1.1616 | **0.0159** | NA | NA | 0.0004 | 4 |
| CASK | NA | NA | 1.2334 | **0.0172** | 0.0014 | 5 |
| CASP3 | -1.3050 | **0.0002** | NA | NA | 0.0014 | 6 |
| CASP6 | -1.2854 | **0.0004** | NA | NA | 0.0002 | 2 |
| CASP8 | -1.2748 | **0.0022** | 1.0232 | 0.9471 | 0.0008 | 3 |
| CASP9 | -1.1387 | **0.0103** | 1.0014 | 0.9861 | 0.0002 | 3 |
| CASS4 | NA | NA | -1.3028 | **0.0034** | 0.0000 | 1 |
| CAT | -1.1852 | **0.0249** | 1.1336 | 0.1949 | 0.0001 | 2 |
| CAVIN1 | 2.8121 | **2.06E-19** | NA | NA | 0.0000 | 3 |
| CAVIN2 | 4.6047 | **2.59E-18** | NA | NA | 0.0002 | 4 |
| CAVIN3 | 2.3228 | **9.94E-13** | NA | NA | 0.0006 | 6 |
| CBFB | -1.3498 | **2.54E-06** | 1.1096 | 0.2273 | **1.0000** | 2 |
| CBL | 1.0967 | 0.2138 | 1.2111 | **0.0258** | **0.0047** | **18** |
| CBLL1 | -1.2527 | **0.0014** | 1.0175 | 0.9136 | 0.0000 | 1 |
| CBR3 | 1.4246 | **0.0159** | 1.0702 | 0.7524 | 0.0000 | 1 |
| CBX3 | -1.1905 | **0.0205** | NA | NA | **0.0038** | **14** |
| CBX5 | 1.0521 | 0.4953 | 1.2096 | **0.0406** | **0.0044** | **16** |
| CBX6 | 1.2632 | **0.0001** | -1.0182 | 0.8938 | 0.0000 | 1 |
| CBX7 | 2.6588 | **4.97E-16** | -1.0412 | 0.8149 | 0.0002 | 5 |
| CBY1 | -1.1629 | **0.0065** | NA | NA | 0.0001 | 3 |
| CCDC106 | 1.2566 | **0.0006** | -1.1405 | 0.1654 | 0.0000 | 3 |
| CCDC130 | -1.2034 | **0.0012** | 1.0357 | 0.7528 | 0.0001 | 6 |
| CCDC170 | -1.7022 | **3.08E-06** | NA | NA | 0.0000 | 1 |
| CCDC28A | 1.2825 | **0.0009** | 1.0581 | 0.6149 | 0.0000 | 2 |
| CCDC85B | 1.5547 | **2.51E-08** | NA | NA | **0.0111** | **45** |
| CCDC88C | -1.3821 | **0.0001** | NA | NA | 0.0000 | 1 |
| CCDC91 | 1.4078 | **0.0005** | NA | NA | 0.0008 | 2 |
| CCDC92 | 1.1444 | **0.0249** | 1.0403 | 0.7934 | 0.0001 | 2 |
| CCL2 | 5.5532 | **2.01E-14** | -1.3128 | 0.1524 | 0.0005 | 3 |
| CCL21 | 2.8405 | **8.50E-06** | -1.6927 | **0.0212** | 0.0000 | 1 |
| CCL4 | 1.0557 | 0.8021 | -1.5362 | **0.0259** | 0.0000 | 1 |
| CCN1 | 2.8099 | **3.12E-06** | NA | NA | 0.0002 | 2 |
| CCNA1 | -2.5972 | **0.0002** | 1.2110 | 0.3189 | 0.0003 | 3 |
| CCNC | -1.2849 | **0.0001** | 1.2078 | 0.0650 | **0.0043** | **14** |
| CCND1 | -1.4486 | **0.0017** | NA | NA | 0.0000 | 4 |
| CCND2 | -1.1435 | 0.2706 | 1.3959 | **0.0266** | 0.0001 | 4 |
| CCND3 | 1.5816 | **1.05E-10** | -1.1258 | 0.2096 | 0.0016 | 11 |
| CCNE1 | -1.9600 | **1.17E-08** | 1.2033 | 0.1145 | 0.0000 | 2 |
| CCNG1 | 1.3426 | **0.0012** | NA | NA | 0.0000 | 3 |
| CCNH | -1.2015 | **0.0199** | NA | NA | **0.0038** | **15** |
| CCNI | 1.2708 | **0.0001** | -1.1014 | 0.5142 | 0.0000 | 2 |
| CCNK | -1.2305 | **0.0047** | NA | NA | **0.0032** | **15** |
| CCNL2 | -1.1910 | **0.0053** | -1.0557 | 0.7316 | 0.0000 | 2 |
| CCNT1 | 1.1810 | **0.0025** | NA | NA | **0.0040** | 5 |
| CCT2 | -1.6542 | **1.22E-12** | -1.0005 | 0.9960 | 0.0000 | 2 |
| CCT3 | -1.3614 | **6.90E-08** | 1.1693 | 0.3585 | 0.0010 | 5 |
| CCT4 | -1.1318 | **0.0382** | -1.0047 | 0.9592 | 0.0004 | 3 |
| CCT5 | -1.6363 | **7.00E-13** | -1.0809 | 0.2260 | 0.0000 | 1 |
| CCT6A | -1.5078 | **4.95E-08** | NA | NA | 0.0000 | 1 |
| CCT6B | -1.3513 | **0.0025** | 1.0029 | 0.9828 | 0.0000 | 1 |
| CCT7 | -1.1939 | **0.0002** | -1.1065 | 0.3775 | 0.0002 | 2 |
| CCT8 | -1.1955 | **0.0230** | 1.0396 | 0.7214 | 0.0000 | 1 |
| CD2 | 1.7087 | **0.0016** | 1.0649 | 0.7319 | 0.0000 | 2 |
| CD27 | 1.4007 | **0.0008** | -1.0894 | 0.5845 | 0.0000 | 1 |
| CD2AP | -1.5019 | **2.76E-06** | NA | NA | 0.0004 | 3 |
| CD302 | 3.7014 | **4.32E-17** | NA | NA | 0.0000 | 2 |
| CD37 | 2.0061 | **1.28E-06** | -1.2217 | 0.1260 | 0.0000 | 1 |
| CD46 | -1.3302 | **1.65E-07** | 1.2626 | 0.0506 | 0.0000 | 1 |
| CD69 | 2.1746 | **0.0001** | -1.1920 | 0.4765 | 0.0016 | 4 |
| CD72 | 1.2418 | **0.0011** | -1.1214 | 0.1514 | 0.0006 | 3 |
| CD81 | -1.2599 | **0.0005** | 1.0484 | 0.7734 | 0.0013 | 8 |
| CD82 | 1.6033 | **8.53E-07** | 1.1183 | 0.3606 | 0.0000 | 1 |
| CD93 | 2.0096 | **3.26E-13** | -1.0832 | 0.4123 | 0.0000 | 1 |
| CD99 | 1.4280 | **4.19E-10** | -1.0453 | 0.6838 | 0.0000 | 1 |
| CDC14B | 1.1178 | 0.1994 | 1.3982 | **0.0091** | 0.0000 | 1 |
| CDC23 | -1.2456 | **0.0001** | 1.0122 | 0.9136 | **0.0070** | **18** |
| CDC42 | -1.2125 | **0.0004** | -1.0104 | 0.9044 | **0.0070** | **14** |
| CDC42BPA | 1.3401 | **5.24E-07** | 1.1739 | 0.1478 | 0.0000 | 1 |
| CDC42EP1 | 1.9328 | **1.18E-12** | -1.6349 | **0.0400** | 0.0010 | 5 |
| CDC42EP3 | 2.4446 | **5.20E-10** | NA | NA | 0.0000 | 2 |
| CDC45 | -2.3610 | **1.04E-07** | 1.2484 | 0.1808 | 0.0000 | 1 |
| CDC5L | -1.0003 | 0.9967 | 1.2492 | **0.0032** | **0.0034** | **17** |
| CDC7 | -1.6486 | **0.0005** | NA | NA | **0.0069** | **15** |
| CDCA8 | -2.2742 | **1.68E-07** | 1.2646 | 0.1435 | 0.0014 | 8 |
| CDIP1 | 1.1496 | **0.0236** | NA | NA | 0.0001 | 3 |
| CDIPT | 1.4072 | **1.05E-05** | NA | NA | 0.0014 | 13 |
| CDK10 | -1.3128 | **1.89E-06** | 1.2082 | 0.0506 | 0.0001 | 2 |
| CDK12 | -1.1683 | **0.0282** | 1.1135 | 0.1700 | 0.0003 | 2 |
| CDK13 | -1.1984 | **0.0372** | 1.3252 | **0.0313** | 0.0000 | 1 |
| CDK18 | 1.5149 | **1.13E-06** | NA | NA | **0.0040** | **16** |
| CDK19 | -1.2242 | **0.0013** | 1.0407 | 0.6332 | 0.0000 | 1 |
| CDK20 | -1.4415 | **2.53E-07** | -1.0927 | 0.2089 | 0.0000 | 2 |
| CDK5 | -1.5036 | **2.02E-06** | NA | NA | 0.0009 | 8 |
| CDK5RAP2 | 1.2707 | **0.0035** | 1.1673 | 0.2236 | 0.0000 | 1 |
| CDK5RAP3 | -1.2119 | **0.0003** | -1.0186 | 0.8643 | 0.0000 | 1 |
| CDK6 | 1.2812 | **0.0105** | 1.1262 | 0.1571 | 0.0012 | 10 |
| CDKL5 | 1.1769 | **0.0134** | 1.0051 | 0.9675 | 0.0001 | 2 |
| CDKN1A | 1.4723 | **0.0005** | 1.0265 | 0.8465 | **0.0172** | **29** |
| CDKN1B | 1.3095 | **0.0004** | 1.0113 | 0.9188 | 0.0002 | 8 |
| CDKN1C | 1.5524 | **0.0022** | -1.0650 | 0.6951 | 0.0000 | 1 |
| CDKN2C | 1.2629 | **0.0128** | NA | NA | 0.0003 | 8 |
| CDKN2D | -1.2544 | **0.0036** | -1.0639 | 0.6898 | 0.0021 | 12 |
| CDT1 | -2.2059 | **2.85E-07** | NA | NA | 0.0000 | 3 |
| CDYL | -1.5763 | **1.01E-09** | 1.0309 | 0.8437 | 0.0000 | 4 |
| CEBPB | 1.6286 | **2.66E-05** | -1.2359 | 0.1240 | 0.0000 | 2 |
| CEBPE | NA | NA | -1.2734 | **0.0459** | 0.0012 | 6 |
| CELF2 | 2.5611 | **1.60E-12** | 1.2783 | 0.0504 | 0.0000 | 1 |
| CENPJ | -1.4034 | **1.16E-05** | NA | NA | 0.0000 | 1 |
| CENPO | -1.2409 | **0.0032** | NA | NA | 0.0001 | 4 |
| CENPU | -2.1388 | **4.57E-06** | NA | NA | 0.0000 | 2 |
| CENPX | -1.4843 | **9.73E-06** | NA | NA | 0.0000 | 1 |
| CEP170 | 1.3792 | **0.0001** | NA | NA | 0.0004 | 6 |
| CEP63 | 1.2319 | **0.0152** | NA | NA | 0.0007 | 13 |
| CEP68 | 1.3837 | **0.0003** | 1.2142 | 0.1768 | 0.0009 | 5 |
| CEP72 | -1.2549 | **0.0047** | -1.0660 | 0.5653 | 0.0001 | 3 |
| CEP76 | -1.3270 | **0.0006** | 1.0056 | 0.9642 | **0.0122** | **30** |
| CEP85 | -1.3650 | **0.0001** | NA | NA | 0.0000 | 2 |
| CETP | -1.3342 | **0.0010** | NA | NA | 0.0000 | 1 |
| CFAP20 | -1.1351 | **0.0063** | NA | NA | 0.0000 | 1 |
| CFDP1 | -1.1524 | **0.0231** | 1.2437 | 0.0543 | 0.0000 | 1 |
| CFL1 | -1.3821 | **2.30E-07** | 1.1220 | 0.4786 | 0.0008 | 2 |
| CFP | 2.5124 | **7.54E-10** | -1.0607 | 0.6983 | 0.0000 | 4 |
| CHD4 | -1.2320 | **0.0001** | 1.0971 | 0.4730 | **0.0045** | 8 |
| CHEK2 | -1.2090 | **0.0241** | 1.0569 | 0.6237 | 0.0000 | 3 |
| CHFR | -1.2802 | **3.98E-05** | -1.0471 | 0.6920 | 0.0000 | 4 |
| CHIC2 | -1.2130 | **0.0027** | NA | NA | **0.0027** | **14** |
| CHMP1B | -1.1745 | **0.0431** | NA | NA | 0.0000 | 2 |
| CHMP2A | -1.3016 | **9.64E-07** | -1.0273 | 0.8229 | 0.0000 | 2 |
| CHMP4A | -1.2791 | **0.0020** | -1.0363 | 0.6819 | 0.0003 | 3 |
| CHORDC1 | -1.3543 | **0.0097** | NA | NA | 0.0000 | 3 |
| CHRNB1 | 1.2022 | **0.0163** | 1.0361 | 0.7893 | 0.0000 | 1 |
| CHTOP | -1.2701 | **2.75E-09** | 1.0037 | 0.9578 | 0.0001 | 2 |
| CIAPIN1 | -1.4071 | **1.64E-07** | NA | NA | 0.0000 | 1 |
| CIB1 | -1.4631 | **2.30E-06** | 1.1128 | 0.3138 | **0.0039** | 10 |
| CIC | 1.1919 | **0.0100** | -1.0849 | 0.5866 | 0.0011 | 3 |
| CIDEB | 1.2701 | **4.62E-05** | -1.2353 | 0.0576 | **0.0112** | **20** |
| CIR1 | 1.2880 | **0.0035** | NA | NA | 0.0008 | 3 |
| CIRBP | 1.1381 | 0.0871 | 1.3027 | **0.0182** | 0.0001 | 2 |
| CITED2 | 2.1925 | **2.75E-05** | -1.1335 | 0.4996 | 0.0000 | 1 |
| CIZ1 | 1.1669 | **0.0256** | NA | NA | 0.0024 | 2 |
| CLDN1 | 2.1542 | **1.81E-05** | 1.0208 | 0.9229 | 0.0008 | 2 |
| CLDN10 | -7.7759 | **1.11E-11** | 1.0509 | 0.8148 | 0.0000 | 2 |
| CLDN5 | 3.4687 | **9.90E-19** | 1.0178 | 0.9209 | **0.0037** | 11 |
| CLEC1A | 1.8526 | **4.00E-08** | NA | NA | 0.0001 | 2 |
| CLINT1 | -1.2520 | **1.83E-05** | NA | NA | 0.0000 | 1 |
| CLIP3 | 1.7077 | **1.97E-10** | -1.1843 | 0.2208 | 0.0000 | 2 |
| CLK1 | 1.3269 | **0.0321** | NA | NA | 0.0005 | 6 |
| CLK2 | -1.2157 | **3.39E-06** | 1.1005 | 0.3853 | **0.0067** | **19** |
| CLK4 | 1.2633 | **0.0120** | NA | NA | 0.0000 | 1 |
| CLN6 | -1.2664 | **0.0013** | NA | NA | 0.0000 | 3 |
| CLP1 | -1.2117 | **0.0006** | 1.0531 | 0.5007 | 0.0000 | 4 |
| CLSTN3 | 1.0172 | 0.8534 | -1.3363 | **0.0260** | 0.0000 | 2 |
| CLTC | -1.2456 | **0.0009** | NA | NA | **0.0046** | 9 |
| CLU | 3.8094 | **5.11E-11** | 1.1777 | 0.6003 | **0.0033** | 11 |
| CLUAP1 | -1.2922 | **3.76E-05** | 1.1520 | **0.0490** | 0.0000 | 2 |
| CMTM6 | -2.1743 | **5.55E-12** | 1.1873 | 0.1273 | 0.0012 | 7 |
| CNDP2 | -2.5236 | **4.59E-12** | 1.3089 | 0.1573 | 0.0000 | 1 |
| CNNM3 | -1.1888 | **0.0405** | NA | NA | 0.0017 | 10 |
| CNOT1 | -1.3123 | **7.00E-08** | -1.0267 | 0.7785 | 0.0001 | 3 |
| CNOT3 | -1.1520 | **0.0169** | -1.0632 | 0.6003 | 0.0000 | 2 |
| CNOT4 | 1.1182 | **0.0411** | -1.0335 | 0.6735 | 0.0007 | 3 |
| CNOT7 | -1.1981 | **0.0026** | 1.1134 | 0.2302 | 0.0001 | 3 |
| CNOT8 | -1.4557 | **7.92E-08** | -1.0055 | 0.9744 | 0.0008 | 4 |
| CNOT9 | -1.4134 | **3.79E-08** | NA | NA | 0.0000 | 2 |
| CNPY2 | -1.4417 | **4.15E-09** | -1.0023 | 0.9746 | 0.0000 | 1 |
| CNTNAP2 | 2.3902 | **0.0058** | 1.0810 | 0.5262 | 0.0001 | 2 |
| COBLL1 | 1.4579 | **3.19E-06** | NA | NA | 0.0000 | 1 |
| COG2 | -1.1576 | **0.0068** | 1.0029 | 0.9835 | 0.0000 | 1 |
| COG4 | -1.2015 | **0.0002** | -1.1564 | **0.0464** | 0.0002 | 3 |
| COIL | -1.1137 | **0.0174** | 1.1988 | **0.0148** | **0.0042** | **14** |
| COL18A1 | 1.5460 | **6.24E-06** | 1.0264 | 0.8721 | 0.0005 | 2 |
| COL4A1 | -1.2129 | **0.0416** | -1.2002 | 0.1877 | 0.0000 | 1 |
| COMMD4 | -1.4044 | **8.34E-07** | NA | NA | 0.0000 | 1 |
| COPA | -1.6342 | **1.41E-06** | 1.2515 | **0.0253** | **0.0026** | 4 |
| COPB1 | -1.1563 | **0.0128** | NA | NA | 0.0008 | 5 |
| COPB2 | -1.4279 | **1.29E-06** | 1.0266 | 0.7534 | 0.0025 | 4 |
| COPE | -1.4630 | **2.38E-06** | NA | NA | 0.0006 | 3 |
| COPG1 | -1.4529 | **2.64E-09** | NA | NA | 0.0016 | 3 |
| COPS3 | -1.3020 | **4.57E-05** | 1.0668 | 0.4015 | 0.0024 | 5 |
| COPS5 | -1.2145 | **0.0072** | 1.0438 | 0.6463 | **0.0040** | 10 |
| COPZ1 | -1.1377 | **0.0026** | 1.0332 | 0.7157 | 0.0000 | 1 |
| COPZ2 | 1.4686 | **2.06E-06** | NA | NA | 0.0000 | 1 |
| COQ3 | -1.1387 | **0.0370** | -1.0470 | 0.5032 | 0.0000 | 1 |
| COQ8A | 1.5972 | **5.19E-09** | NA | NA | 0.0017 | 7 |
| COQ9 | -1.3664 | **1.81E-08** | NA | NA | 0.0015 | 7 |
| CORIN | 1.8323 | **0.0062** | 1.0684 | 0.7147 | 0.0000 | 1 |
| COX5A | -1.7430 | **1.24E-12** | -1.0126 | 0.9110 | 0.0003 | 2 |
| COX6C | -1.3803 | **2.17E-05** | -1.2924 | **0.0065** | 0.0000 | 2 |
| COX7C | 1.1007 | **0.0353** | NA | NA | 0.0000 | 1 |
| CPNE7 | 1.3104 | **0.0276** | -1.1641 | 0.5748 | 0.0005 | 9 |
| CPSF6 | -1.2868 | **0.0003** | 1.3408 | **0.0112** | 0.0011 | 9 |
| CRBN | 1.3242 | **6.06E-06** | 1.2329 | **0.0491** | 0.0000 | 3 |
| CREB3L1 | -1.3572 | **0.0057** | -1.0684 | 0.7303 | **0.0111** | **58** |
| CREBL2 | NA | NA | 1.1920 | **0.0476** | 0.0001 | 3 |
| CREM | 1.2550 | **0.0335** | -1.0951 | 0.3936 | 0.0006 | 7 |
| CRK | -1.2284 | **0.0001** | NA | NA | **0.0101** | **25** |
| CRKL | -1.0890 | **0.0279** | NA | NA | 0.0016 | 9 |
| CRMP1 | -1.2952 | **0.0058** | NA | NA | 0.0000 | 2 |
| CSDE1 | 1.3723 | **4.38E-05** | NA | NA | 0.0000 | 1 |
| CSE1L | -1.1875 | **0.0254** | NA | NA | 0.0000 | 1 |
| CSF3R | NA | NA | -1.4437 | **0.0247** | 0.0008 | 2 |
| CSGALNACT2 | 1.7136 | **8.18E-07** | NA | NA | 0.0017 | 5 |
| CSN1S1 | NA | NA | -1.2197 | **0.0010** | 0.0000 | 1 |
| CSNK1A1 | -1.1952 | **0.0004** | 1.0476 | 0.5581 | 0.0000 | 1 |
| CSNK1D | -1.2868 | **2.27E-06** | NA | NA | 0.0001 | 3 |
| CSNK1E | -1.2546 | **0.0002** | 1.1051 | 0.3650 | 0.0005 | 4 |
| CSNK1G1 | -1.3647 | **1.46E-09** | 1.1515 | 0.1680 | 0.0000 | 1 |
| CSNK2A1 | -1.1069 | **0.0395** | 1.3565 | **0.0072** | **0.0055** | **17** |
| CSNK2A2 | 1.3807 | **6.68E-10** | 1.0418 | 0.7627 | 0.0000 | 1 |
| CSPG5 | -1.3835 | **0.0429** | 1.0194 | 0.8388 | 0.0000 | 1 |
| CSPP1 | -1.0688 | 0.3812 | 1.3344 | **0.0218** | 0.0001 | 2 |
| CSRNP2 | -1.1843 | **0.0047** | NA | NA | 0.0000 | 1 |
| CSRP1 | 2.3026 | **1.16E-11** | 1.0241 | 0.9119 | 0.0000 | 2 |
| CST1 | -2.3117 | **0.0215** | 1.1921 | 0.4643 | 0.0001 | 2 |
| CSTF1 | -1.2054 | **0.0004** | 1.0225 | 0.8065 | 0.0000 | 1 |
| CSTF2 | -1.3507 | **8.91E-07** | -1.0926 | 0.3509 | 0.0015 | 9 |
| CTBP1 | -1.1281 | **0.0398** | NA | NA | 0.0014 | 9 |
| CTBP2 | -1.3184 | **0.0001** | 1.1021 | 0.1508 | **0.0030** | **17** |
| CTCF | -1.1592 | **0.0095** | 1.2266 | 0.1193 | 0.0000 | 1 |
| CTH | NA | NA | 1.3451 | **0.0332** | 0.0000 | 1 |
| CTNNA1 | -1.2765 | **4.88E-07** | -1.1045 | 0.1485 | 0.0008 | 3 |
| CTNNBL1 | 1.3176 | **2.69E-08** | 1.0716 | 0.5777 | 0.0005 | 5 |
| CTNND1 | -1.3417 | **5.62E-08** | -1.1179 | 0.1851 | 0.0008 | 2 |
| CTNND2 | NA | NA | 1.3311 | **0.0230** | 0.0000 | 1 |
| CTPS1 | -1.4086 | **0.0050** | NA | NA | 0.0000 | 1 |
| CTPS2 | -1.3596 | **8.03E-07** | NA | NA | 0.0000 | 1 |
| CUL4A | -1.3041 | **0.0001** | -1.0112 | 0.9030 | 0.0001 | 2 |
| CUTC | 1.3461 | **1.35E-06** | 1.1713 | 0.1435 | 0.0002 | 5 |
| CX3CL1 | 1.4120 | **0.0025** | -1.0658 | 0.6945 | 0.0000 | 1 |
| CX3CR1 | 1.5794 | **0.0165** | 1.0290 | 0.9170 | 0.0000 | 1 |
| CXCR5 | 1.0164 | 0.8018 | -1.1994 | **0.0409** | 0.0000 | 1 |
| CYB561 | -2.3368 | **7.77E-15** | 1.1471 | 0.1266 | 0.0017 | **17** |
| CYB561D2 | -1.3400 | **2.75E-05** | -1.0997 | 0.3618 | 0.0001 | 4 |
| CYB5R2 | -1.3969 | **0.0016** | 1.0824 | 0.4790 | 0.0012 | 8 |
| CYB5R3 | 1.4788 | **1.28E-10** | 1.0125 | 0.8885 | 0.0014 | **14** |
| CYLD | 1.6655 | **6.34E-13** | 1.0763 | 0.3599 | 0.0002 | 3 |
| CYREN | -1.3394 | **7.38E-06** | NA | NA | 0.0000 | 1 |
| CYSLTR2 | 1.4025 | **0.0310** | 1.0314 | 0.7683 | 0.0000 | 1 |
| CYTH2 | -1.2733 | **0.0010** | -1.1409 | 0.3887 | 0.0008 | 2 |
| DAAM2 | 1.3175 | **0.0022** | -1.0952 | 0.4882 | 0.0001 | 4 |
| DAB1 | -1.5630 | **0.0060** | -1.0171 | 0.9453 | **0.0043** | **14** |
| DAB2 | 1.2323 | **0.0127** | -1.0668 | 0.5579 | 0.0000 | 1 |
| DACH1 | -1.8814 | **2.08E-06** | 1.1058 | 0.6021 | 0.0015 | 9 |
| DAP3 | -1.0906 | **0.0312** | -1.0557 | 0.3084 | 0.0000 | 2 |
| DAPK2 | -1.3164 | **0.0026** | NA | NA | 0.0000 | 1 |
| DARS | -1.2164 | **0.0057** | NA | NA | 0.0001 | 2 |
| DARS2 | -1.4523 | **1.97E-10** | 1.0160 | 0.8881 | 0.0000 | 4 |
| DAXX | -1.3069 | **0.0002** | NA | NA | **0.0204** | **40** |
| DBN1 | -1.3163 | **0.0007** | NA | NA | 0.0000 | 1 |
| DCAF1 | -1.1831 | **0.0001** | NA | NA | 0.0002 | 3 |
| DCAF11 | -1.1718 | **0.0110** | 1.1464 | 0.2230 | 0.0000 | 1 |
| DCAF8 | 1.0901 | **0.0452** | 1.0055 | 0.9644 | 0.0002 | 4 |
| DCLRE1A | -1.1777 | **0.0032** | 1.1632 | 0.0682 | 0.0000 | 1 |
| DCTN5 | -1.3440 | **3.52E-06** | NA | NA | 0.0000 | 1 |
| DDA1 | -1.1784 | **0.0244** | NA | NA | 0.0000 | 1 |
| DDIT3 | 1.2501 | **0.0123** | -1.1524 | 0.4941 | 0.0005 | 8 |
| DDR1 | NA | NA | 1.2535 | **0.0133** | 0.0018 | 3 |
| DDX21 | -1.5697 | **1.13E-06** | 1.1481 | 0.1572 | 0.0011 | 3 |
| DDX39A | -1.6289 | **7.43E-10** | -1.0995 | 0.2528 | 0.0013 | 5 |
| DDX3X | -1.2383 | **0.0009** | 1.2420 | **0.0402** | 0.0000 | 1 |
| DDX41 | -1.2430 | **0.0003** | NA | NA | 0.0000 | 1 |
| DDX46 | -1.3269 | **0.0001** | NA | NA | 0.0000 | 1 |
| DDX56 | -1.2796 | **2.52E-06** | -1.2673 | **0.0058** | 0.0000 | 1 |
| DEAF1 | -1.3081 | **0.0025** | NA | NA | 0.0000 | 1 |
| DEDD | 1.1111 | **0.0351** | NA | NA | 0.0000 | 1 |
| DEF6 | 1.2487 | **0.0342** | -1.1840 | 0.1582 | 0.0011 | 7 |
| DEFB1 | -10.3884 | **8.33E-09** | 1.1279 | 0.7044 | 0.0000 | 1 |
| DENR | -1.2002 | **0.0045** | NA | NA | 0.0000 | 1 |
| DERL2 | -1.2948 | **1.06E-05** | -1.0196 | 0.8177 | 0.0000 | 1 |
| DES | 6.7427 | **9.81E-13** | -1.4551 | 0.2014 | **0.0038** | **14** |
| DESI1 | -1.3321 | **0.0001** | NA | NA | 0.0001 | 5 |
| DESI2 | 1.2017 | **0.0223** | NA | NA | 0.0013 | 5 |
| DEXI | 1.4046 | **9.98E-07** | NA | NA | 0.0000 | 1 |
| DGCR8 | -1.1249 | **0.0473** | -1.1137 | 0.3316 | 0.0000 | 1 |
| DGLUCY | -1.3641 | **9.08E-07** | NA | NA | 0.0000 | 1 |
| DHRS7 | -1.2056 | **0.0153** | -1.0387 | 0.8443 | 0.0000 | 1 |
| DHX15 | -1.5372 | **1.77E-11** | 1.1104 | 0.3347 | 0.0010 | 5 |
| DHX16 | -1.1212 | **0.0090** | 1.1749 | 0.3477 | 0.0000 | 2 |
| DHX9 | -1.4278 | **3.28E-05** | 1.0696 | 0.5090 | 0.0001 | 2 |
| DIDO1 | 1.0105 | 0.8375 | 1.1234 | **0.0358** | 0.0002 | 3 |
| DIMT1 | -1.2769 | **4.81E-05** | 1.0164 | 0.8792 | 0.0000 | 1 |
| DIO2 | -3.6042 | **7.24E-07** | 1.2338 | 0.2913 | 0.0010 | 4 |
| DKC1 | -1.3055 | **9.42E-06** | 1.0168 | 0.8483 | 0.0008 | 2 |
| DLAT | -1.5648 | **1.47E-07** | 1.1067 | 0.4488 | 0.0000 | 1 |
| DLD | -1.2554 | **0.0011** | NA | NA | 0.0001 | 2 |
| DLG1 | -1.4337 | **1.90E-06** | NA | NA | 0.0025 | 6 |
| DLG4 | 1.2539 | **0.0003** | -1.3215 | 0.1560 | **0.0027** | 6 |
| DLK2 | -1.3964 | **0.0001** | -1.1788 | **0.0312** | 0.0000 | 1 |
| DMXL1 | 1.5429 | **0.0006** | NA | NA | 0.0000 | 1 |
| DNAJA2 | -1.1713 | **0.0080** | 1.2820 | **0.0189** | 0.0020 | 3 |
| DNAJA3 | -1.1857 | **0.0008** | NA | NA | **0.0037** | 8 |
| DNAJA4 | -1.1371 | **0.0363** | 1.0263 | 0.7524 | 0.0008 | 2 |
| DNAJB4 | 2.5448 | **3.96E-13** | 1.2822 | **0.0171** | 0.0000 | 1 |
| DNAJC11 | -1.3086 | **1.11E-07** | NA | NA | 0.0001 | 2 |
| DNAJC15 | -3.0752 | **3.00E-14** | 1.0971 | 0.6136 | 0.0000 | 1 |
| DNAJC3 | -1.1435 | **0.0284** | 1.5947 | **0.0167** | 0.0000 | 1 |
| DNAJC8 | 1.0949 | **0.0355** | NA | NA | 0.0000 | 1 |
| DNAJC9 | -1.4019 | **0.0003** | -1.0324 | 0.8347 | 0.0000 | 4 |
| DNAL4 | -1.4839 | **6.37E-09** | -1.0303 | 0.7180 | 0.0017 | 5 |
| DNALI1 | -1.2403 | **0.0119** | 1.1563 | 0.1374 | 0.0000 | 1 |
| DNM1 | -1.4267 | **0.0113** | NA | NA | 0.0000 | 1 |
| DNM2 | -1.1897 | **0.0039** | 1.2561 | 0.2302 | 0.0021 | 7 |
| DNMT3B | -1.6397 | **1.68E-07** | 1.1120 | 0.4337 | 0.0000 | 1 |
| DNPEP | -1.1953 | **0.0034** | NA | NA | 0.0000 | 2 |
| DOC2A | 1.1727 | **0.0247** | -1.1180 | 0.1083 | 0.0000 | 1 |
| DOCK2 | NA | NA | -1.1754 | **0.0374** | **0.0029** | 4 |
| DOCK3 | 1.2513 | **0.0424** | -1.4588 | 0.1226 | 0.0005 | 2 |
| DOCK4 | 2.7179 | **5.46E-15** | -1.0951 | 0.4595 | 0.0008 | 2 |
| DOK2 | 1.6129 | **0.0010** | -1.2057 | 0.0615 | 0.0002 | 5 |
| DOK4 | 1.2692 | **1.77E-05** | -1.2397 | 0.0892 | 0.0000 | 1 |
| DOLK | -1.1225 | **0.0387** | NA | NA | 0.0000 | 3 |
| DOT1L | -1.2976 | **0.0091** | 1.1619 | 0.1878 | 0.0000 | 1 |
| DPH2 | -1.2618 | **0.0002** | 1.0636 | 0.4730 | 0.0000 | 1 |
| DPM3 | -1.1502 | **0.0250** | 1.0983 | 0.3889 | 0.0000 | 1 |
| DPP3 | -1.9207 | **1.79E-11** | NA | NA | 0.0000 | 1 |
| DPYSL2 | 1.5880 | **6.51E-07** | -1.0601 | 0.6283 | 0.0001 | 2 |
| DPYSL4 | -1.6852 | **0.0002** | NA | NA | 0.0000 | 1 |
| DRG2 | -1.1120 | **0.0372** | -1.1463 | 0.1200 | 0.0000 | 1 |
| DSG2 | -1.8120 | **4.11E-05** | 1.5813 | **0.0035** | 0.0000 | 1 |
| DSN1 | -1.6422 | **2.18E-05** | 1.1279 | 0.5068 | 0.0000 | 1 |
| DSTN | 1.5937 | **3.47E-07** | -1.0816 | 0.3927 | 0.0000 | 1 |
| DTNB | -1.1482 | **0.0309** | NA | NA | 0.0010 | 11 |
| DTX2 | 1.1488 | **0.0486** | 1.0208 | 0.8525 | **0.0121** | **33** |
| DTX3 | 1.3891 | **4.00E-05** | -1.0167 | 0.8738 | 0.0008 | 9 |
| DUS1L | -1.2868 | **2.35E-06** | 1.0921 | 0.4882 | 0.0000 | 1 |
| DUSP1 | 2.4296 | **9.67E-06** | -2.3998 | **0.0054** | 0.0000 | 3 |
| DUSP13 | NA | NA | -1.3667 | **0.0260** | 0.0006 | 5 |
| DUSP9 | NA | NA | -1.2474 | **0.0406** | 0.0000 | 1 |
| DVL3 | 1.1283 | **0.0075** | -1.1766 | 0.1658 | **0.0131** | **41** |
| DYNC1LI1 | -1.2626 | **0.0017** | -1.0027 | 0.9785 | 0.0000 | 1 |
| DYNLT1 | -1.5721 | **1.01E-13** | -1.1397 | 0.6905 | **0.0049** | 10 |
| DYNLT3 | -1.8468 | **0.0003** | NA | NA | 0.0000 | 2 |
| DYRK1B | -1.1516 | **0.0117** | -1.3935 | 0.0933 | 0.0000 | 2 |
| DYRK2 | 1.3475 | **0.0003** | 1.1312 | 0.1607 | 0.0002 | 3 |
| DYRK3 | 1.0631 | 0.4068 | 1.1865 | **0.0075** | 0.0002 | 2 |
| DYRK4 | 1.1859 | **0.0005** | -1.0894 | 0.3356 | 0.0000 | 2 |
| DYSF | 1.3405 | **0.0214** | -1.1776 | 0.2136 | 0.0016 | 8 |
| E2F1 | -1.6065 | **0.0001** | NA | NA | 0.0015 | 8 |
| EBAG9 | -1.0765 | 0.3333 | 1.3962 | **0.0022** | 0.0001 | 5 |
| EBNA1BP2 | -1.2497 | **0.0075** | 1.0846 | 0.4426 | 0.0000 | 1 |
| EBP | -1.5537 | **2.28E-06** | 1.0545 | 0.6910 | **0.0132** | **56** |
| ECD | -1.1637 | **0.0016** | 1.0281 | 0.7202 | 0.0000 | 1 |
| ECHDC1 | -1.2742 | **0.0034** | 1.0544 | 0.7444 | 0.0000 | 1 |
| ECI2 | -1.5912 | **0.0003** | -1.1150 | 0.4888 | 0.0000 | 1 |
| ECM1 | -2.9183 | **9.22E-08** | -1.1319 | 0.5634 | 0.0007 | 9 |
| ECPAS | -1.1826 | **0.0269** | NA | NA | 0.0000 | 1 |
| ECSIT | 1.2587 | **0.0006** | NA | NA | 0.0011 | 6 |
| EDA | -1.2411 | **0.0179** | 1.1163 | 0.2494 | 0.0009 | 6 |
| EDC3 | -1.1669 | **0.0070** | NA | NA | 0.0000 | 2 |
| EDN1 | 1.3289 | **0.0215** | 1.1224 | 0.5336 | 0.0000 | 1 |
| EDNRA | -1.6929 | **0.0001** | -1.1801 | 0.4278 | 0.0008 | 2 |
| EEF1AKNMT | -1.2613 | **0.0001** | NA | NA | 0.0000 | 1 |
| EEF1D | 1.2758 | **6.39E-07** | -1.0026 | 0.9754 | **0.0032** | 6 |
| EEF1E1 | -1.9491 | **1.55E-10** | NA | NA | 0.0008 | 2 |
| EEF2KMT | -1.3542 | **4.44E-05** | NA | NA | 0.0000 | 2 |
| EFCAB2 | -1.4727 | **1.37E-06** | 1.2761 | **0.0344** | 0.0000 | 2 |
| EFEMP1 | 5.0213 | **6.57E-13** | -1.2402 | 0.2947 | 0.0002 | 7 |
| EFEMP2 | 1.7167 | **4.27E-11** | NA | NA | **0.0081** | **26** |
| EFHC1 | -1.3953 | **0.0001** | NA | NA | **0.0041** | **18** |
| EFHC2 | -2.1548 | **1.17E-06** | NA | NA | **0.0038** | **29** |
| EFNB2 | -1.5683 | **1.40E-05** | NA | NA | 0.0009 | 3 |
| EFS | -1.2533 | **0.0010** | -1.2429 | **0.0078** | 0.0025 | 13 |
| EGFL7 | 2.0468 | **7.47E-13** | -1.1040 | 0.4325 | 0.0000 | 1 |
| EGLN3 | -1.3990 | **0.0088** | 1.1246 | 0.4191 | **0.0042** | 13 |
| EGR1 | 2.5081 | **3.68E-05** | -3.0794 | **0.0008** | 0.0002 | 2 |
| EHD1 | 1.2363 | **0.0098** | NA | NA | 0.0000 | 2 |
| EHHADH | -1.2536 | **0.0081** | 1.0802 | 0.5439 | **0.0247** | **38** |
| EI24 | -1.2935 | **2.19E-05** | 1.2659 | 0.0665 | 0.0000 | 1 |
| EID1 | 1.3396 | **0.0013** | NA | NA | 0.0000 | 1 |
| EIF2AK2 | 1.1519 | 0.0615 | 1.4702 | **0.0182** | 0.0008 | 5 |
| EIF2B5 | -1.1540 | **0.0014** | 1.1709 | 0.2044 | 0.0000 | 1 |
| EIF3A | -1.0509 | 0.4566 | 1.4422 | **0.0034** | 0.0023 | 8 |
| EIF3B | -1.2060 | **0.0001** | NA | NA | 0.0004 | 5 |
| EIF3F | 1.2134 | **0.0006** | 1.1094 | 0.4274 | 0.0021 | 11 |
| EIF3G | 1.2525 | **9.36E-06** | 1.0200 | 0.8361 | 0.0007 | 6 |
| EIF3I | -1.1610 | **0.0028** | 1.2060 | 0.2774 | 0.0001 | 2 |
| EIF3J | -1.3251 | **0.0002** | NA | NA | 0.0000 | 1 |
| EIF4A2 | 1.0906 | 0.2541 | 1.2715 | **0.0289** | 0.0000 | 2 |
| EIF4A3 | -1.1898 | **0.0054** | -1.0427 | 0.6037 | **0.0036** | 11 |
| EIF4E | -1.1808 | 0.1216 | 1.5345 | **0.0022** | 0.0009 | 5 |
| EIF4E2 | -1.5407 | **1.35E-09** | -1.0233 | 0.8344 | 0.0024 | **16** |
| EIF4EBP1 | 1.1138 | 0.2326 | -1.3615 | **0.0230** | 0.0001 | 4 |
| EIF4G1 | -1.2318 | **0.0009** | NA | NA | **0.0041** | 9 |
| EIF4G3 | -1.0074 | 0.9304 | 1.3097 | **0.0113** | 0.0000 | 1 |
| EIF4H | -1.1009 | **0.0454** | NA | NA | 0.0000 | 2 |
| EIF5A | -1.2231 | **0.0043** | NA | NA | 0.0000 | 1 |
| EIF5A2 | NA | NA | -1.2055 | **0.0333** | 0.0000 | 3 |
| EIF6 | -1.3823 | **1.86E-05** | NA | NA | 0.0000 | 1 |
| ELAVL1 | -1.2135 | **3.06E-05** | -1.0508 | 0.4144 | 0.0015 | 4 |
| ELK1 | 1.5094 | **0.0027** | 1.1099 | 0.3959 | 0.0002 | 3 |
| ELK3 | 1.7740 | **4.60E-07** | -1.0125 | 0.9502 | 0.0002 | 3 |
| ELMO1 | 2.8753 | **5.69E-21** | 1.0026 | 0.9803 | 0.0024 | 4 |
| ELMO2 | -1.1485 | **0.0077** | -1.0253 | 0.8066 | 0.0000 | 1 |
| ELN | 1.4801 | **0.0082** | NA | NA | 0.0000 | 3 |
| ELOB | -1.3141 | **1.19E-06** | NA | NA | 0.0000 | 2 |
| ELOC | -1.1289 | **0.0467** | NA | NA | **0.0041** | 6 |
| ELOVL5 | -1.3630 | **0.0014** | 1.1676 | 0.3247 | 0.0001 | 7 |
| ELOVL6 | -1.5333 | **0.0028** | 1.1101 | 0.4101 | 0.0000 | 1 |
| EMC2 | 1.2317 | **0.0122** | NA | NA | 0.0009 | 3 |
| EMC3 | -1.1402 | **0.0096** | NA | NA | 0.0000 | 1 |
| EMC6 | -1.2186 | **0.0015** | NA | NA | 0.0000 | 4 |
| EMC8 | -1.1908 | **0.0032** | NA | NA | 0.0000 | 1 |
| EMC9 | -1.2429 | **0.0002** | NA | NA | 0.0001 | 2 |
| EMP3 | 1.8324 | **1.05E-11** | -1.2598 | 0.0512 | 0.0015 | 11 |
| ENAH | 1.5380 | **0.0002** | 1.2695 | **0.0258** | 0.0008 | 2 |
| ENDOG | -1.8106 | **5.77E-07** | 1.2130 | 0.1694 | 0.0000 | 1 |
| ENO1 | -1.5681 | **7.33E-06** | 1.1613 | 0.3465 | 0.0013 | 4 |
| ENOX1 | NA | NA | -1.2497 | **0.0239** | 0.0000 | 3 |
| ENOX2 | 1.2029 | **0.0088** | -1.0848 | 0.4415 | 0.0003 | 5 |
| ENPP4 | -1.2972 | **0.0069** | NA | NA | 0.0000 | 2 |
| ENTPD3 | -1.8902 | **0.0004** | 1.2566 | 0.3562 | 0.0001 | 4 |
| ENTR1 | -1.2204 | **0.0001** | NA | NA | 0.0000 | 1 |
| ENY2 | -1.3263 | **4.13E-05** | NA | NA | 0.0001 | 2 |
| EPAS1 | NA | NA | -1.3680 | **0.0202** | 0.0009 | 4 |
| EPB41L1 | 1.2876 | **4.60E-05** | NA | NA | 0.0000 | 1 |
| EPHA4 | 2.5897 | **3.25E-13** | 1.0769 | 0.6290 | 0.0000 | 1 |
| EPN1 | 1.1005 | **0.0309** | -1.2681 | **0.0097** | 0.0002 | 3 |
| EPS15 | 1.4806 | **5.54E-06** | NA | NA | 0.0022 | 6 |
| EPS8 | NA | NA | 1.3842 | **0.0379** | 0.0003 | 6 |
| ERAL1 | -1.1676 | **0.0054** | -1.0519 | 0.6877 | 0.0000 | 1 |
| ERC2 | 1.3148 | **0.0270** | 1.0247 | 0.7863 | 0.0000 | 1 |
| ERCC1 | 1.1517 | **0.0188** | NA | NA | 0.0000 | 3 |
| ERCC2 | 1.1464 | **0.0193** | -1.2187 | 0.2781 | 0.0000 | 2 |
| ERF | -1.1689 | **0.0422** | NA | NA | 0.0000 | 1 |
| ERG28 | -1.1868 | **0.0158** | NA | NA | 0.0016 | **14** |
| ERGIC3 | -1.2022 | **0.0117** | -1.1312 | 0.3085 | **0.0184** | **58** |
| ERH | -1.2787 | **1.83E-05** | 1.1318 | 0.2518 | 0.0001 | 2 |
| ERLIN1 | -1.5127 | **9.02E-13** | 1.0974 | 0.2893 | 0.0001 | 3 |
| ERMP1 | -1.7250 | **1.16E-08** | 1.0250 | 0.8394 | 0.0005 | 6 |
| ERP29 | -1.1057 | 0.0956 | 1.3273 | **0.0473** | 0.0010 | 5 |
| ESR2 | 1.4990 | **0.0096** | -1.0194 | 0.8065 | 0.0010 | 6 |
| ESYT1 | 1.1707 | **0.0050** | NA | NA | 0.0000 | 1 |
| ETF1 | -1.2607 | **0.0008** | 1.1147 | 0.1773 | 0.0016 | 4 |
| ETHE1 | -1.3877 | **0.0008** | NA | NA | 0.0000 | 1 |
| ETNK1 | -1.8317 | **2.25E-09** | 1.2516 | 0.0606 | 0.0002 | 2 |
| ETNK2 | 1.3859 | **0.0007** | NA | NA | 0.0001 | 2 |
| ETS2 | -1.5423 | **0.0001** | 1.0592 | 0.6541 | 0.0001 | 2 |
| ETV6 | 1.1501 | **0.0474** | NA | NA | 0.0006 | 5 |
| EVI5 | 1.3087 | **0.0024** | NA | NA | 0.0001 | 5 |
| EVL | -1.1253 | 0.1126 | -1.2114 | **0.0179** | 0.0000 | 2 |
| EWSR1 | -1.2128 | **9.97E-06** | -1.1838 | 0.1577 | **0.0147** | **30** |
| EXOC3 | 1.1569 | **0.0034** | -1.2261 | **0.0352** | 0.0000 | 1 |
| EXOC6B | 1.1486 | **0.0149** | 1.1653 | **0.0287** | 0.0000 | 1 |
| EXOC7 | -1.1281 | **0.0119** | 1.0218 | 0.7697 | 0.0007 | 8 |
| EXOSC2 | -1.1797 | **0.0095** | 1.0424 | 0.7061 | 0.0000 | 2 |
| EXOSC4 | -1.2980 | **0.0002** | -1.1957 | 0.1761 | 0.0005 | 4 |
| EXOSC5 | -1.1308 | **0.0328** | NA | NA | 0.0011 | **14** |
| EXOSC7 | -1.2060 | **3.33E-05** | NA | NA | 0.0003 | 3 |
| EXOSC8 | -1.1983 | **0.0158** | NA | NA | 0.0024 | **20** |
| EXOSC9 | -1.1550 | **0.0418** | 1.0134 | 0.8942 | 0.0004 | 2 |
| EXT1 | 1.3920 | **0.0014** | NA | NA | 0.0000 | 1 |
| EXT2 | 1.2173 | **1.43E-05** | NA | NA | 0.0000 | 1 |
| EYA3 | -1.2042 | **0.0036** | 1.2590 | **0.0050** | 0.0000 | 1 |
| EZR | -2.0465 | **8.60E-07** | 1.4158 | **0.0045** | 0.0021 | 8 |
| F11R | -1.6847 | **7.52E-09** | 1.0661 | 0.5756 | 0.0010 | 6 |
| F12 | -1.2527 | **0.0090** | -1.0347 | 0.7506 | 0.0000 | 1 |
| F2RL1 | -2.0964 | **2.06E-05** | NA | NA | 0.0000 | 1 |
| F8 | 3.3979 | **2.48E-20** | NA | NA | 0.0000 | 2 |
| FA2H | NA | NA | 1.3285 | **0.0239** | 0.0005 | 10 |
| FABP3 | 2.8040 | **4.93E-12** | -1.0077 | 0.9600 | 0.0002 | 2 |
| FADD | -1.2185 | **0.0052** | 1.0923 | 0.1564 | **0.0038** | 12 |
| FAF2 | -1.1714 | **0.0002** | -1.0243 | 0.7785 | 0.0000 | 1 |
| FAH | 1.4201 | **0.0041** | NA | NA | 0.0001 | 3 |
| FAIM | -1.6253 | **3.51E-08** | NA | NA | 0.0000 | 1 |
| FAM102A | -1.2752 | **0.0007** | -1.1631 | 0.1485 | 0.0016 | 3 |
| FAM110B | 1.3482 | **3.91E-05** | NA | NA | 0.0000 | 2 |
| FAM114A1 | 1.1909 | **0.0129** | -1.0688 | 0.4942 | 0.0000 | 1 |
| FAM136A | -1.3079 | **0.0001** | -1.0315 | 0.7300 | 0.0001 | 4 |
| FAM168B | 1.1422 | **0.0055** | NA | NA | 0.0010 | **18** |
| FAM173A | -1.1964 | **0.0234** | NA | NA | 0.0005 | 5 |
| FAM184A | 1.6180 | **0.0003** | -1.0527 | 0.6285 | 0.0000 | 3 |
| FAM192A | -1.1245 | **0.0045** | 1.0929 | 0.1776 | 0.0000 | 1 |
| FAM214B | 1.1745 | **0.0042** | NA | NA | **0.0028** | **16** |
| FAM222B | -1.2452 | **0.0003** | NA | NA | 0.0012 | 8 |
| FAM3C | -1.4057 | **0.0153** | NA | NA | 0.0010 | 13 |
| FAM49B | 1.3307 | **0.0005** | 1.0688 | 0.6718 | 0.0000 | 2 |
| FAM50B | 1.4346 | **5.40E-07** | 1.0252 | 0.7974 | **0.0031** | **18** |
| FAM8A1 | 1.3550 | **0.0001** | NA | NA | 0.0000 | 1 |
| FANCA | -1.4819 | **1.59E-07** | 1.0770 | 0.5126 | 0.0001 | 4 |
| FANCF | -1.4699 | **2.31E-07** | 1.0266 | 0.7738 | 0.0005 | 3 |
| FANCG | -1.3091 | **0.0002** | NA | NA | **0.0035** | 11 |
| FARP1 | -1.6139 | **1.54E-09** | NA | NA | 0.0000 | 1 |
| FARSA | -1.2852 | **0.0001** | NA | NA | 0.0000 | 2 |
| FAS | 1.6054 | **0.0001** | NA | NA | 0.0008 | 6 |
| FASLG | -1.3230 | **0.0406** | -1.3822 | **0.0120** | 0.0020 | 10 |
| FASTKD5 | -1.2612 | **4.23E-05** | -1.0731 | 0.2327 | 0.0000 | 2 |
| FAXDC2 | 1.8485 | **5.48E-10** | NA | NA | 0.0006 | 9 |
| FBL | -1.1895 | **0.0049** | NA | NA | 0.0000 | 1 |
| FBLN5 | 1.5418 | **0.0031** | NA | NA | 0.0000 | 2 |
| FBXL2 | 1.5450 | **1.46E-07** | 1.1039 | 0.3446 | 0.0000 | 1 |
| FBXL5 | 1.1620 | **0.0078** | -1.0678 | 0.5098 | 0.0000 | 1 |
| FBXL7 | NA | NA | -1.2566 | **0.0222** | 0.0000 | 1 |
| FBXO17 | 1.4811 | **8.92E-08** | -1.2768 | 0.3053 | 0.0007 | 7 |
| FBXO24 | 1.0673 | 0.5092 | -1.3765 | **0.0007** | 0.0000 | 2 |
| FBXO28 | -1.4503 | **1.32E-07** | NA | NA | 0.0011 | 8 |
| FCER1G | 2.2190 | **4.34E-06** | -1.2725 | 0.1213 | **0.0027** | **22** |
| FDFT1 | -1.4354 | **2.03E-07** | -1.0276 | 0.8148 | 0.0000 | 1 |
| FDPS | -1.4861 | **1.55E-09** | 1.0032 | 0.9775 | 0.0002 | 2 |
| FEM1C | -1.4297 | **0.0002** | 1.2898 | **0.0165** | 0.0000 | 2 |
| FER | 1.4750 | **1.05E-08** | 1.0604 | 0.6615 | 0.0000 | 1 |
| FES | 1.6850 | **9.72E-10** | NA | NA | 0.0000 | 1 |
| FEZ1 | 2.1467 | **3.35E-17** | NA | NA | 0.0011 | 5 |
| FEZ2 | 1.3458 | **4.11E-08** | -1.0989 | 0.1134 | 0.0002 | 2 |
| FGF13 | -1.3042 | **0.0181** | 1.2488 | 0.0686 | 0.0013 | 3 |
| FGFR1 | 1.5268 | **9.42E-07** | 1.0731 | 0.4123 | 0.0018 | 7 |
| FGFR1OP | -1.7398 | **1.41E-08** | NA | NA | 0.0000 | 1 |
| FH | -1.6415 | **1.55E-07** | 1.0195 | 0.8844 | 0.0003 | 7 |
| FHL5 | 3.9045 | **8.52E-15** | 1.0842 | 0.4375 | **0.0037** | **23** |
| FHOD1 | 1.3425 | **2.65E-05** | NA | NA | 0.0000 | 1 |
| FILIP1L | 4.4181 | **3.88E-12** | -1.2089 | 0.3068 | 0.0000 | 1 |
| FKBP1B | -1.2381 | **0.0464** | 1.0064 | 0.9485 | 0.0001 | 3 |
| FKBP2 | -1.2510 | **0.0004** | 1.1280 | 0.1764 | 0.0008 | 2 |
| FKBP3 | -1.2145 | **0.0108** | NA | NA | 0.0009 | 3 |
| FKBP4 | -1.6429 | **1.43E-08** | NA | NA | 0.0013 | 3 |
| FKBP8 | 1.2968 | **2.11E-06** | -1.0474 | 0.6387 | **0.0063** | 12 |
| FLAD1 | -1.4039 | **4.13E-08** | -1.1142 | 0.1990 | 0.0016 | 8 |
| FLI1 | 1.9759 | **4.12E-08** | 1.0096 | 0.9494 | 0.0001 | 3 |
| FLNA | 1.7407 | **1.51E-06** | NA | NA | **0.0156** | **20** |
| FLNC | 2.2112 | **1.08E-07** | -1.0817 | 0.5594 | 0.0008 | 5 |
| FLT1 | 1.9069 | **1.50E-10** | -1.0321 | 0.7225 | 0.0006 | 4 |
| FMNL1 | NA | NA | -1.3031 | **0.0032** | 0.0003 | 2 |
| FNBP1 | 1.4162 | **3.80E-06** | -1.0403 | 0.7396 | 0.0000 | 1 |
| FNTA | -1.2407 | **0.0004** | NA | NA | **0.6667** | 2 |
| FOLR1 | -2.1506 | **0.0002** | 1.3833 | 0.1553 | 0.0000 | 1 |
| FOS | 3.9180 | **3.50E-06** | -5.3142 | **0.0010** | **0.0047** | **16** |
| FOSB | 2.4952 | **0.0083** | -5.4065 | **0.0007** | 0.0007 | 10 |
| FOXH1 | -1.2408 | **0.0289** | -1.0011 | 0.9937 | 0.0006 | 11 |
| FOXJ2 | 1.1129 | **0.0210** | NA | NA | 0.0000 | 1 |
| FOXL2 | -1.7160 | **0.0051** | NA | NA | 0.0000 | 1 |
| FOXO4 | 1.4937 | **1.12E-07** | NA | NA | 0.0000 | 1 |
| FRS2 | -1.0434 | 0.5781 | 1.2495 | **0.0100** | 0.0006 | 6 |
| FRS3 | 1.0797 | 0.1801 | -1.2561 | **0.0050** | **0.0049** | **23** |
| FRYL | -1.1052 | 0.2419 | 1.3426 | **0.0200** | 0.0000 | 1 |
| FTH1 | 1.2796 | **0.0107** | 1.0658 | 0.5488 | 0.0002 | 3 |
| FTO | 1.2798 | **3.55E-06** | -1.0634 | 0.5697 | 0.0003 | 5 |
| FUS | -1.2894 | **1.15E-06** | 1.2240 | 0.1456 | **0.0030** | 8 |
| FXR1 | 1.2528 | **0.0003** | NA | NA | 0.0014 | 13 |
| FYCO1 | 1.6404 | **2.06E-07** | -1.0434 | 0.6198 | 0.0000 | 1 |
| FYN | 1.2889 | **0.0004** | -1.1186 | 0.1963 | 0.0017 | 11 |
| FZD7 | 15.0355 | **9.08E-31** | -1.1524 | 0.4075 | 0.0006 | 4 |
| G0S2 | 1.9244 | **0.0123** | -1.0465 | 0.8837 | 0.0000 | 2 |
| G3BP1 | -1.2348 | **0.0001** | NA | NA | 0.0016 | 2 |
| G3BP2 | -1.4173 | **0.0001** | 1.0907 | 0.2948 | 0.0000 | 1 |
| GAB2 | 1.7316 | **4.23E-07** | -1.3800 | **0.0024** | 0.0004 | 5 |
| GABARAPL2 | 1.4467 | **1.65E-11** | NA | NA | **0.0083** | **18** |
| GABPB1 | -1.1780 | **0.0013** | -1.0191 | 0.8536 | 0.0009 | 7 |
| GADD45B | 2.1017 | **3.90E-06** | NA | NA | 0.0010 | 3 |
| GAK | -1.2005 | **0.0001** | 1.0969 | 0.3042 | 0.0000 | 1 |
| GALNT2 | 1.3819 | **0.0002** | NA | NA | 0.0000 | 1 |
| GAN | -1.2209 | **0.0003** | 1.0354 | 0.7834 | 0.0013 | 4 |
| GAPDH | -1.4333 | **9.95E-07** | 1.0707 | 0.6565 | **0.0039** | 6 |
| GARS | -1.1601 | **0.0058** | -1.0693 | 0.6548 | 0.0000 | 1 |
| GATAD2A | -1.3215 | **1.22E-05** | 1.2986 | **0.0092** | 0.0015 | 2 |
| GCDH | -1.2275 | **0.0001** | NA | NA | 0.0000 | 1 |
| GCFC2 | -1.2154 | **0.0052** | NA | NA | 0.0000 | 1 |
| GCH1 | 1.2891 | **0.0304** | 1.0094 | 0.9435 | 0.0002 | 3 |
| GCN1 | -1.1793 | **0.0008** | NA | NA | 0.0000 | 1 |
| GDAP1 | 1.0408 | 0.6696 | 1.3692 | **0.0022** | 0.0008 | 2 |
| GDF15 | -2.0589 | **0.0025** | -1.0995 | 0.7075 | 0.0000 | 1 |
| GDPD5 | 1.2113 | **0.0129** | NA | NA | 0.0008 | 2 |
| GEM | 2.5590 | **0.0001** | -1.3418 | 0.1869 | **0.0053** | **30** |
| GEMIN2 | -1.3907 | **1.59E-06** | 1.0416 | 0.6820 | 0.0001 | 4 |
| GFOD1 | 1.7140 | **3.40E-06** | 1.0254 | 0.8920 | 0.0000 | 1 |
| GGH | -1.8303 | **0.0007** | 1.1652 | 0.4883 | 0.0000 | 1 |
| GHR | 2.8054 | **1.80E-11** | 1.0743 | 0.6441 | **0.0029** | 7 |
| GIGYF2 | -1.1091 | **0.0075** | 1.0228 | 0.7983 | 0.0000 | 1 |
| GIMAP6 | 3.7565 | **9.81E-20** | 1.0214 | 0.8812 | 0.0002 | 3 |
| GINS2 | -2.0314 | **1.86E-06** | 1.1957 | 0.2884 | 0.0000 | 2 |
| GINS4 | -1.7778 | **6.93E-07** | 1.0739 | 0.6081 | 0.0003 | 3 |
| GIPC1 | -1.4376 | **1.18E-07** | NA | NA | 0.0017 | 5 |
| GIT1 | -1.1327 | **0.0298** | -1.0373 | 0.7651 | 0.0010 | 4 |
| GJA1 | -1.5320 | **0.0015** | -1.1208 | 0.5227 | 0.0000 | 1 |
| GJB1 | NA | NA | 1.5405 | **0.0416** | 0.0010 | 12 |
| GJB3 | -1.5860 | **0.0430** | -1.1662 | 0.2859 | 0.0000 | 3 |
| GLE1 | -1.2530 | **6.04E-06** | 1.0938 | 0.2409 | 0.0000 | 2 |
| GLI1 | -1.6250 | **0.0043** | -1.0989 | 0.4429 | 0.0000 | 1 |
| GLI2 | 1.4841 | **0.0002** | -1.1458 | 0.2119 | 0.0000 | 1 |
| GLI3 | 1.8648 | **5.81E-14** | -1.0760 | 0.5986 | 0.0008 | 3 |
| GLMN | -1.2097 | **0.0377** | 1.1239 | 0.2439 | 0.0001 | 3 |
| GLRX3 | -1.2104 | **0.0021** | 1.1565 | 0.2472 | **0.0114** | **21** |
| GLS | 2.1041 | **4.44E-13** | -1.0570 | 0.6290 | 0.0000 | 1 |
| GLUL | 1.8997 | **4.31E-09** | 1.2480 | 0.1303 | 0.0000 | 1 |
| GLYR1 | -1.2714 | **0.0001** | 1.0310 | 0.6547 | 0.0000 | 2 |
| GMCL1 | -1.2054 | **0.0123** | 1.4959 | **0.0007** | **0.0106** | **26** |
| GMDS | -1.3123 | **0.0004** | NA | NA | 0.0000 | 1 |
| GMNN | -2.6470 | **1.14E-10** | 1.2476 | 0.1836 | 0.0001 | 3 |
| GMPPA | -1.2058 | **0.0013** | -1.1413 | 0.1383 | 0.0001 | 4 |
| GMPPB | -1.8627 | **4.19E-10** | NA | NA | 0.0000 | 2 |
| GNA12 | 1.4862 | **4.26E-08** | NA | NA | 0.0000 | 1 |
| GNA14 | NA | NA | -1.2451 | **0.0162** | 0.0000 | 1 |
| GNA15 | 1.6505 | **1.67E-05** | -1.1406 | 0.2373 | 0.0000 | 1 |
| GNAI2 | 1.3658 | **0.0002** | NA | NA | 0.0010 | 4 |
| GNAI3 | -1.2287 | **0.0011** | 1.0398 | 0.6649 | 0.0019 | 5 |
| GNB1 | -1.0995 | **0.0101** | 1.2371 | 0.0627 | **0.0035** | 7 |
| GNB2 | -1.2216 | **0.0008** | NA | NA | 0.0009 | 3 |
| GNB3 | 1.1313 | 0.1700 | -1.2479 | **0.0032** | 0.0010 | 3 |
| GNB5 | -1.3212 | **0.0003** | 1.0093 | 0.9316 | 0.0000 | 1 |
| GNG11 | 1.9648 | **1.50E-07** | 1.0335 | 0.8675 | 0.0000 | 2 |
| GNG12 | 1.8241 | **2.93E-10** | 1.4399 | **0.0021** | 0.0000 | 1 |
| GNG4 | 1.6090 | **0.0174** | NA | NA | **0.0027** | 4 |
| GNL3L | -1.2512 | **0.0001** | 1.0737 | 0.5886 | 0.0002 | 5 |
| GNLY | -12.8858 | **3.20E-13** | -1.9902 | **0.0196** | 0.0000 | 1 |
| GOLGA2 | -1.1570 | **0.0170** | 1.0071 | 0.9624 | **0.0685** | **116** |
| GOLGA5 | -1.1995 | **0.0061** | NA | NA | 0.0000 | 1 |
| GOLPH3 | -1.1900 | **0.0151** | 1.1015 | 0.5593 | 0.0000 | 1 |
| GOLPH3L | -1.1281 | **0.0251** | 1.0890 | 0.3480 | 0.0001 | 2 |
| GOLT1B | -1.3438 | **0.0027** | -1.0977 | 0.3113 | 0.0000 | 3 |
| GORASP1 | -1.3998 | **6.93E-08** | -1.0207 | 0.8369 | 0.0010 | 5 |
| GORASP2 | -1.2603 | **2.19E-06** | NA | NA | **0.0229** | **54** |
| GOSR1 | -1.1796 | **0.0013** | 1.3210 | **0.0234** | 0.0000 | 1 |
| GP1BA | 1.2336 | **0.0175** | -1.1522 | 0.2810 | 0.0015 | 4 |
| GPATCH2 | -1.2907 | **0.0013** | NA | NA | 0.0000 | 2 |
| GPBP1L1 | -1.1022 | 0.1077 | 1.2293 | **0.0139** | 0.0002 | 2 |
| GPHN | -1.3816 | **0.0001** | 1.1284 | 0.2981 | 0.0000 | 1 |
| GPKOW | -1.2111 | **0.0020** | -1.0211 | 0.8078 | **0.0032** | **19** |
| GPM6A | 3.1014 | **1.13E-09** | -1.1309 | 0.2444 | 0.0000 | 1 |
| GPS1 | -1.1425 | **0.0228** | NA | NA | 0.0000 | 1 |
| GPX7 | -1.5727 | **4.11E-06** | -1.2331 | 0.0755 | 0.0012 | 5 |
| GRAP2 | 1.2603 | **0.0438** | -1.1491 | **0.0337** | 0.0008 | 8 |
| GRIN2A | 1.5957 | **0.0186** | -1.0392 | 0.5309 | 0.0002 | 2 |
| GRIP1 | -1.3050 | **0.0055** | 1.1797 | **0.0499** | **0.0027** | 6 |
| GRM3 | NA | NA | -1.1786 | **0.0243** | 0.0008 | 2 |
| GRPEL1 | -1.2810 | **9.63E-06** | NA | NA | 0.0000 | 1 |
| GSE1 | -1.4177 | **2.29E-05** | NA | NA | 0.0002 | 5 |
| GSN | 1.6656 | **3.58E-07** | 1.1857 | 0.1777 | 0.0000 | 1 |
| GSPT1 | -1.4977 | **1.31E-11** | 1.1749 | 0.1885 | 0.0000 | 1 |
| GSPT2 | 1.3195 | **0.0004** | NA | NA | 0.0000 | 1 |
| GSTCD | -1.2241 | **0.0223** | NA | NA | 0.0000 | 1 |
| GSTM3 | 2.2180 | **4.14E-11** | NA | NA | 0.0000 | 1 |
| GSTM5 | 2.3108 | **1.40E-07** | 1.1091 | 0.5090 | 0.0008 | 3 |
| GSTP1 | -1.4897 | **5.11E-07** | NA | NA | 0.0007 | 4 |
| GSTZ1 | -2.0450 | **4.93E-10** | 1.1445 | 0.1836 | 0.0000 | 2 |
| GTF2A1 | 1.0459 | 0.4142 | 1.2033 | **0.0390** | 0.0008 | 4 |
| GTF2A2 | -1.1996 | **0.0022** | -1.0229 | 0.8465 | 0.0000 | 1 |
| GTF2H2C | -1.4344 | **0.0095** | NA | NA | 0.0002 | 2 |
| GTF2H5 | 1.1365 | **0.0318** | -1.1984 | 0.0608 | 0.0000 | 1 |
| GTF2IRD1 | -1.3043 | **0.0001** | -1.0049 | 0.9693 | 0.0001 | 2 |
| GTPBP2 | -1.1289 | **0.0414** | -1.3615 | **0.0020** | 0.0007 | 6 |
| GTPBP3 | -1.1503 | **0.0271** | NA | NA | 0.0008 | 8 |
| GTPBP4 | -1.2962 | **9.53E-06** | -1.0043 | 0.9588 | 0.0008 | 3 |
| GTPBP8 | 1.1464 | **0.0074** | 1.0975 | 0.4102 | 0.0000 | 1 |
| GYPC | 1.9821 | **2.55E-18** | NA | NA | 0.0008 | 2 |
| H1F0 | -1.2594 | **0.0081** | -1.1298 | 0.3559 | 0.0000 | 1 |
| H2AFX | -1.8304 | **2.05E-07** | NA | NA | 0.0008 | 3 |
| H2AFY | -1.4073 | **4.50E-07** | NA | NA | 0.0008 | 2 |
| H3F3B | -1.3064 | **5.76E-08** | 1.0274 | 0.7316 | 0.0009 | 4 |
| HACD2 | -1.2679 | **0.0033** | NA | NA | 0.0000 | 3 |
| HAND1 | NA | NA | -1.1622 | **0.0289** | 0.0000 | 2 |
| HCCS | -1.2204 | **0.0002** | NA | NA | 0.0000 | 1 |
| HCK | NA | NA | -1.5395 | **0.0265** | 0.0019 | 12 |
| HCLS1 | 2.4021 | **7.51E-09** | -1.0673 | 0.7240 | 0.0000 | 1 |
| HDAC1 | -1.3944 | **6.48E-11** | -1.0431 | 0.5187 | 0.0022 | 10 |
| HDAC2 | -1.2710 | **0.0017** | NA | NA | 0.0002 | 4 |
| HDAC5 | 1.3172 | **0.0005** | -1.0087 | 0.9522 | 0.0000 | 2 |
| HDHD3 | -1.3917 | **5.23E-06** | 1.0289 | 0.7602 | 0.0000 | 3 |
| HDLBP | -1.1268 | **0.0200** | NA | NA | 0.0000 | 1 |
| HEBP2 | -1.4325 | **1.72E-08** | 1.1492 | 0.1849 | 0.0000 | 2 |
| HECTD3 | 1.2811 | **0.0007** | NA | NA | 0.0000 | 1 |
| HELLS | NA | NA | 1.3731 | **0.0289** | 0.0000 | 1 |
| HEPH | 1.5173 | **0.0002** | NA | NA | 0.0000 | 1 |
| HERC1 | 1.2023 | **0.0014** | -1.1182 | 0.1312 | 0.0000 | 1 |
| HEXIM1 | -1.4107 | **2.62E-06** | 1.1746 | 0.1260 | 0.0000 | 1 |
| HGF | 1.3334 | **0.0263** | -1.1964 | 0.3561 | 0.0000 | 1 |
| HGS | -1.1622 | **0.0152** | 1.0820 | 0.3757 | **0.0381** | **75** |
| HHLA3 | 1.2393 | **0.0014** | -1.1163 | 0.4680 | 0.0000 | 2 |
| HIF1A | -1.2575 | **0.0246** | 1.0091 | 0.9525 | **0.0051** | 9 |
| HILPDA | -1.4059 | **0.0003** | 1.1910 | 0.1346 | 0.0000 | 1 |
| HINT1 | -1.5983 | **2.06E-14** | 1.0125 | 0.8494 | 0.0000 | 1 |
| HIP1 | -1.2489 | **0.0017** | 1.1693 | 0.0528 | 0.0006 | 5 |
| HIPK1 | 1.1470 | **0.0347** | 1.3696 | **0.0020** | 0.0002 | 2 |
| HIST1H3D | NA | NA | 1.5501 | **0.0046** | 0.0017 | **17** |
| HIST1H3E | -1.2768 | **0.0222** | 1.3182 | **0.0072** | 0.0017 | **17** |
| HIST1H3F | NA | NA | 1.4316 | **0.0101** | 0.0017 | **17** |
| HIST1H3H | -1.5359 | **0.0002** | NA | NA | 0.0017 | **17** |
| HIST1H4E | NA | NA | 1.7067 | **0.0213** | 0.0023 | 10 |
| HIST1H4F | NA | NA | 1.4762 | **0.0036** | 0.0023 | 10 |
| HIST1H4H | -1.2560 | **0.0076** | NA | NA | 0.0023 | 10 |
| HIST1H4L | NA | NA | 1.6706 | **0.0486** | 0.0023 | 10 |
| HLCS | -1.2224 | **2.74E-05** | -1.0594 | 0.3615 | 0.0000 | 1 |
| HLX | 2.4612 | **9.28E-10** | NA | NA | 0.0001 | 3 |
| HMG20B | -1.4218 | **1.15E-07** | NA | NA | 0.0002 | 4 |
| HMGA1 | -2.0930 | **1.06E-07** | -1.1649 | 0.3312 | **0.0066** | **21** |
| HMGB3 | -3.0671 | **1.47E-20** | 1.4495 | **0.0161** | 0.0001 | 2 |
| HMGCR | -3.3786 | **7.47E-13** | NA | NA | 0.0000 | 1 |
| HMGXB4 | -1.1604 | **0.0017** | -1.0177 | 0.8182 | 0.0003 | 3 |
| HMOX2 | -1.1969 | **0.0005** | 1.0736 | 0.4901 | 0.0011 | 9 |
| HNRNPA1 | -1.1755 | **0.0172** | NA | NA | **0.0031** | 7 |
| HNRNPA2B1 | -1.2116 | **0.0177** | 1.2191 | 0.0679 | 0.0007 | 4 |
| HNRNPA3 | -1.2450 | **0.0009** | NA | NA | 0.0000 | 1 |
| HNRNPAB | -1.6121 | **7.22E-10** | NA | NA | 0.0000 | 1 |
| HNRNPD | -1.1958 | **0.0050** | 1.1083 | 0.1742 | **0.0038** | 7 |
| HNRNPF | -1.4619 | **1.43E-11** | NA | NA | **0.0031** | **15** |
| HNRNPH1 | -1.4302 | **0.0018** | 1.4856 | **0.0256** | **0.0043** | **15** |
| HNRNPH2 | -1.1880 | **0.0163** | 1.1498 | 0.1885 | 0.0000 | 1 |
| HNRNPL | -1.1395 | **0.0340** | NA | NA | 0.0001 | 4 |
| HNRNPM | -1.2855 | **3.22E-05** | 1.1144 | 0.3133 | 0.0020 | 9 |
| HNRNPR | -1.4208 | **4.26E-06** | 1.1080 | 0.3419 | 0.0003 | 2 |
| HNRNPU | NA | NA | 1.2027 | **0.0491** | 0.0008 | 4 |
| HOMER1 | -1.8037 | **2.22E-06** | 1.0712 | 0.5892 | 0.0000 | 2 |
| HOMER3 | 1.3151 | **0.0037** | NA | NA | **0.0048** | **20** |
| HPGDS | 1.4978 | **0.0009** | -1.0938 | 0.3475 | 0.0000 | 1 |
| HPRT1 | -1.8196 | **1.85E-07** | 1.0082 | 0.9651 | 0.0021 | 4 |
| HRAS | -1.2076 | **0.0041** | -1.1602 | 0.1729 | **0.0029** | 8 |
| HSBP1 | -1.2460 | **3.92E-05** | 1.1262 | 0.0733 | 0.0001 | 3 |
| HSD17B10 | -1.3507 | **2.06E-06** | NA | NA | 0.0000 | 1 |
| HSD17B11 | 2.0874 | **2.60E-10** | NA | NA | 0.0019 | **18** |
| HSF4 | 1.0956 | 0.1859 | -1.4636 | **0.0023** | 0.0011 | 7 |
| HSP90AA1 | -1.3055 | **0.0010** | NA | NA | **0.0076** | **18** |
| HSP90AB1 | -1.1982 | **0.0056** | 1.0452 | 0.7919 | **0.0242** | **40** |
| HSP90B1 | -1.5614 | **4.72E-09** | NA | NA | 0.0017 | 3 |
| HSPA13 | -1.1967 | **0.0484** | 1.0428 | 0.7721 | 0.0000 | 1 |
| HSPA1A | 1.7488 | **4.91E-05** | -1.1093 | 0.4151 | 0.0003 | 3 |
| HSPA1L | 1.4030 | **1.17E-05** | -1.1075 | 0.1694 | 0.0001 | 2 |
| HSPA2 | -1.4931 | **0.0014** | 1.4027 | **0.0137** | 0.0000 | 1 |
| HSPA4 | -1.3585 | **8.39E-06** | -1.0267 | 0.7463 | 0.0002 | 3 |
| HSPA5 | -1.6232 | **1.46E-09** | NA | NA | 0.0000 | 1 |
| HSPA9 | -1.2914 | **2.36E-05** | 1.0553 | 0.4385 | 0.0016 | 3 |
| HSPB11 | -1.2437 | **0.0001** | NA | NA | 0.0003 | 4 |
| HSPB6 | 1.8740 | **0.0001** | NA | NA | 0.0000 | 1 |
| HSPB8 | 2.9606 | **1.05E-11** | 1.0819 | 0.6436 | 0.0009 | 6 |
| HSPBP1 | -1.2304 | **0.0003** | -1.0230 | 0.8153 | 0.0008 | 2 |
| HSPG2 | 1.6770 | **8.91E-07** | -1.0927 | 0.4503 | 0.0000 | 2 |
| HSPH1 | -1.3611 | **0.0001** | 1.0780 | 0.5977 | 0.0000 | 1 |
| HTRA2 | 1.1642 | **0.0002** | -1.0377 | 0.6771 | 0.0000 | 1 |
| HYAL3 | -1.2981 | **0.0005** | -1.2391 | **0.0120** | 0.0000 | 1 |
| IARS | -1.3351 | **0.0009** | -1.0734 | 0.4497 | 0.0000 | 1 |
| ICAM2 | 2.2376 | **2.04E-15** | NA | NA | 0.0000 | 1 |
| ICK | 1.4100 | **0.0010** | -1.0296 | 0.8732 | 0.0000 | 1 |
| ICMT | -1.2159 | **0.0003** | -1.0668 | 0.4075 | 0.0002 | 3 |
| ID3 | 1.3152 | **0.0339** | 1.1729 | 0.2897 | 0.0001 | 5 |
| IER2 | 1.1569 | 0.2750 | -1.6674 | **0.0077** | 0.0000 | 1 |
| IFI35 | 1.2059 | **0.0170** | NA | NA | 0.0000 | 1 |
| IFITM3 | 1.3784 | **2.71E-05** | -1.1371 | 0.1441 | 0.0001 | 4 |
| IFNAR1 | 1.2455 | **0.0038** | 1.0344 | 0.7956 | 0.0014 | 4 |
| IFNAR2 | 1.4304 | **2.79E-07** | -1.0396 | 0.5682 | 0.0000 | 1 |
| IFRD2 | -1.3284 | **4.37E-05** | NA | NA | 0.0000 | 1 |
| IFT27 | -1.1431 | **0.0325** | NA | NA | 0.0001 | 2 |
| IGBP1 | 1.2367 | **0.0012** | -1.1240 | 0.2125 | 0.0000 | 2 |
| IGF1 | -1.7815 | **0.0008** | 1.0313 | 0.9131 | 0.0008 | 2 |
| IGF1R | -1.4341 | **3.41E-05** | 1.0765 | 0.4899 | 0.0013 | 5 |
| IGFBP4 | 1.5299 | **0.0012** | NA | NA | 0.0000 | 1 |
| IGFBP5 | 2.5914 | **1.69E-08** | -1.5851 | **0.0117** | **0.0028** | 10 |
| IGFBP6 | 2.5295 | **1.09E-06** | NA | NA | **0.0028** | 9 |
| IKZF1 | NA | NA | -1.1364 | **0.0490** | **0.0092** | **38** |
| IL10RA | 2.6702 | **4.53E-09** | -1.2212 | 0.1482 | 0.0002 | 6 |
| IL15 | -2.7572 | **4.74E-07** | -1.0685 | 0.7642 | **0.6667** | 3 |
| IL15RA | -1.1125 | 0.1836 | -1.1774 | **0.0385** | 0.0000 | 1 |
| IL2RB | -2.8512 | **1.71E-08** | -1.2909 | 0.1883 | 0.0000 | 2 |
| IL2RG | 1.4107 | **0.0410** | -1.0802 | 0.4960 | 0.0000 | 2 |
| IL3RA | 1.3861 | **3.66E-07** | 1.0632 | 0.6946 | 0.0000 | 6 |
| ILF2 | -1.7720 | **1.95E-15** | NA | NA | 0.0002 | 2 |
| ILF3 | -1.2911 | **4.29E-05** | 1.0047 | 0.9621 | 0.0008 | 4 |
| ILK | 1.1023 | 0.1300 | -1.2751 | **0.0244** | 0.0019 | 5 |
| IMMT | -1.1523 | **0.0044** | -1.1018 | 0.2398 | 0.0014 | 4 |
| INCENP | -1.4627 | **2.62E-06** | 1.1303 | 0.1932 | 0.0001 | 4 |
| INPPL1 | -1.2331 | **0.0004** | 1.0205 | 0.9286 | 0.0008 | 5 |
| INSIG2 | 1.6857 | **5.39E-06** | NA | NA | 0.0018 | 8 |
| INTS13 | -1.2772 | **0.0064** | NA | NA | 0.0000 | 1 |
| IP6K2 | -1.6210 | **7.62E-12** | NA | NA | 0.0000 | 1 |
| IPCEF1 | 1.6216 | **0.0001** | 1.0191 | 0.8881 | 0.0000 | 1 |
| IPO8 | 1.1848 | **0.0019** | 1.3012 | **0.0013** | 0.0000 | 1 |
| IQSEC1 | 1.4987 | **4.77E-09** | -1.0945 | 0.1364 | 0.0000 | 1 |
| IRAK4 | -1.1759 | **0.0403** | NA | NA | 0.0000 | 3 |
| ISCU | NA | NA | -1.1756 | **0.0450** | 0.0004 | 5 |
| ISOC1 | -1.4432 | **0.0128** | 1.0404 | 0.7807 | 0.0000 | 1 |
| IST1 | -1.0837 | **0.0206** | -1.0764 | 0.2020 | **0.0029** | 5 |
| ITCH | -1.1877 | **0.0069** | 1.2278 | **0.0047** | 0.0004 | 3 |
| ITGA2 | -2.0144 | **4.68E-05** | 1.3310 | 0.0543 | 0.0000 | 1 |
| ITGA2B | -1.2333 | **0.0146** | -1.2352 | 0.2810 | 0.0000 | 1 |
| ITGA4 | NA | NA | 1.3826 | **0.0102** | 0.0000 | 2 |
| ITGA5 | 1.4219 | **0.0002** | -1.3654 | **0.0252** | 0.0001 | 2 |
| ITGAM | 3.3344 | **1.21E-09** | -1.1128 | 0.5050 | 0.0004 | 9 |
| ITGB1 | 1.2193 | **0.0062** | 1.1225 | 0.1346 | **0.0038** | 9 |
| ITGB1BP1 | 1.3058 | **2.26E-05** | 1.0663 | 0.4393 | 0.0004 | 3 |
| ITGB1BP2 | NA | NA | -1.3241 | **0.0425** | 0.0000 | 2 |
| ITM2A | 5.0384 | **1.33E-14** | NA | NA | 0.0000 | 1 |
| ITPKB | 1.2578 | **0.0160** | -1.1897 | 0.1763 | 0.0011 | 5 |
| ITPRID2 | -1.4382 | **0.0005** | NA | NA | 0.0000 | 1 |
| ITSN1 | 1.3062 | **0.0015** | 1.1484 | 0.2108 | 0.0023 | 9 |
| ITSN2 | 1.2076 | **0.0024** | 1.2826 | 0.0568 | 0.0008 | 3 |
| JAK1 | -1.0003 | 0.9963 | 1.4167 | **0.0380** | 0.0002 | 2 |
| JAK2 | 1.1871 | **0.0442** | NA | NA | 0.0013 | 5 |
| JMJD6 | 1.1901 | **0.0063** | 1.1094 | 0.3968 | 0.0008 | 2 |
| JOSD1 | -1.1414 | **0.0411** | -1.0923 | 0.3659 | 0.0000 | 4 |
| JRK | 1.1366 | **0.0346** | -1.0168 | 0.9044 | 0.0023 | 13 |
| JUN | 1.5577 | **0.0059** | 1.0010 | 0.9973 | **0.0103** | **21** |
| JUNB | 2.2058 | **2.19E-05** | -1.9277 | **0.0075** | 0.0003 | 5 |
| KANK2 | 1.1970 | **0.0232** | NA | NA | **0.0114** | **49** |
| KARS | -1.2161 | **1.16E-05** | NA | NA | 0.0000 | 1 |
| KAT2B | 1.8707 | **7.46E-07** | NA | NA | 0.0000 | 1 |
| KAT7 | -1.2400 | **0.0001** | NA | NA | 0.0003 | 5 |
| KAT8 | 1.3501 | **1.13E-07** | 1.1375 | 0.4015 | 0.0000 | 1 |
| KAZN | 1.2306 | **0.0104** | 1.2305 | 0.0524 | 0.0000 | 2 |
| KCNA10 | NA | NA | -1.3804 | **0.0133** | 0.0000 | 2 |
| KCNC1 | NA | NA | 1.2175 | **0.0097** | 0.0000 | 1 |
| KCNE4 | 3.3457 | **2.00E-13** | 1.2679 | 0.3428 | 0.0004 | 3 |
| KCNH2 | 2.4736 | **1.14E-05** | -1.3638 | **0.0178** | 0.0000 | 1 |
| KCNMA1 | 2.0465 | **6.95E-07** | -1.1821 | 0.3666 | 0.0000 | 1 |
| KCNMB1 | 4.6882 | **8.51E-12** | NA | NA | 0.0004 | 2 |
| KCNN3 | 1.8838 | **5.88E-07** | -1.0119 | 0.9309 | 0.0000 | 2 |
| KCTD13 | -1.1142 | **0.0207** | -1.3431 | 0.0980 | **0.0026** | 10 |
| KCTD5 | -1.3281 | **2.46E-05** | NA | NA | 0.0008 | 2 |
| KCTD7 | 1.2716 | **0.0008** | -1.0684 | 0.6093 | 0.0018 | 13 |
| KCTD9 | 1.4064 | **0.0001** | NA | NA | **0.0043** | **22** |
| KDM1A | -1.3767 | **1.52E-08** | -1.2607 | 0.0691 | **0.0109** | **30** |
| KDM4A | -1.1914 | **0.0002** | 1.4301 | 0.0591 | 0.0000 | 8 |
| KDR | 1.6256 | **0.0002** | -1.3238 | 0.0533 | 0.0014 | 4 |
| KEAP1 | -1.1539 | **0.0027** | NA | NA | **0.0067** | **16** |
| KHDC4 | -1.2273 | **0.0029** | NA | NA | 0.0004 | 8 |
| KHDRBS1 | -1.1407 | **0.0371** | -1.0585 | 0.2967 | 0.0015 | 8 |
| KIAA0319L | -1.3253 | **4.00E-07** | 1.2359 | 0.1473 | 0.0000 | 1 |
| KIAA1324 | NA | NA | 1.5722 | **0.0178** | 0.0000 | 1 |
| KIF16B | 1.1802 | **0.0108** | 1.0844 | 0.3524 | 0.0001 | 2 |
| KIF1C | 1.0842 | 0.1584 | -1.2477 | **0.0108** | 0.0000 | 1 |
| KIF3A | -1.0219 | 0.8363 | 1.6480 | **0.0008** | 0.0016 | 2 |
| KIR2DL4 | -2.4335 | **0.0001** | -1.3557 | **0.0418** | 0.0003 | 4 |
| KLF11 | 1.3658 | **0.0003** | -1.1048 | 0.2062 | 0.0000 | 1 |
| KLF2 | 6.8752 | **7.97E-28** | -1.1305 | 0.5698 | 0.0000 | 1 |
| KLF4 | NA | NA | -1.4941 | **0.0268** | 0.0001 | 3 |
| KLF9 | 1.4992 | **0.0148** | NA | NA | 0.0000 | 1 |
| KLHDC3 | 1.1741 | **0.0106** | -1.0207 | 0.8613 | 0.0000 | 1 |
| KLHL12 | -1.2581 | **2.48E-07** | -1.1135 | 0.1393 | **0.0041** | **16** |
| KLHL2 | -1.2163 | **0.0494** | NA | NA | 0.0020 | 9 |
| KLHL24 | 1.2877 | **0.0036** | NA | NA | 0.0000 | 2 |
| KLHL3 | 1.5232 | **5.16E-06** | -1.0936 | 0.2258 | 0.0000 | 3 |
| KMT2A | 1.1650 | **0.0179** | NA | NA | 0.0001 | 5 |
| KMT5B | -1.1856 | **0.0395** | NA | NA | 0.0000 | 1 |
| KPNA3 | -1.0715 | 0.3256 | 1.4576 | **0.0045** | 0.0011 | 11 |
| KPNA6 | -1.1783 | **0.0001** | 1.1675 | 0.1687 | 0.0003 | 6 |
| KRT10 | -1.1959 | **0.0119** | 1.0758 | 0.4944 | 0.0000 | 1 |
| KRT18 | -2.1954 | **5.75E-07** | 1.2672 | 0.0799 | **0.0064** | 11 |
| KRT20 | NA | NA | -1.1683 | **0.0077** | 0.0002 | 9 |
| KRT7 | -3.5595 | **2.72E-06** | 1.2186 | 0.3475 | 0.0000 | 2 |
| KRT75 | NA | NA | 1.1641 | **0.0100** | **0.0069** | **38** |
| LACTB2 | -1.6126 | **0.0003** | 1.1466 | 0.3428 | 0.0000 | 1 |
| LAMTOR2 | -1.2676 | **0.0009** | 1.0890 | 0.3892 | 0.0001 | 2 |
| LAMTOR5 | -1.2080 | **2.26E-05** | NA | NA | 0.0002 | 4 |
| LANCL2 | -1.4334 | **1.28E-06** | NA | NA | 0.0000 | 1 |
| LAPTM4B | -1.6118 | **1.78E-08** | 1.0433 | 0.7103 | 0.0004 | 4 |
| LARP4B | -1.4429 | **2.05E-08** | 1.0555 | 0.4542 | 0.0000 | 1 |
| LATS1 | -1.0508 | 0.4341 | 1.1415 | **0.0494** | 0.0022 | 12 |
| LCP2 | 1.4791 | **0.0026** | 1.0661 | 0.6282 | 0.0021 | 7 |
| LDB1 | 1.3178 | **0.0008** | NA | NA | 0.0001 | 4 |
| LDB2 | 1.3600 | **0.0002** | 1.2020 | 0.2258 | 0.0004 | 9 |
| LDHA | -1.5780 | **1.66E-05** | -1.0470 | 0.6685 | **0.0029** | 4 |
| LDHB | -1.4417 | **7.54E-06** | -1.0444 | 0.6865 | 0.0000 | 2 |
| LDLRAP1 | 1.2633 | **0.0002** | NA | NA | 0.0008 | 3 |
| LDOC1 | 1.3927 | **1.21E-05** | -1.1079 | 0.1916 | **0.0061** | **32** |
| LETM1 | -1.2898 | **0.0001** | -1.0179 | 0.8875 | 0.0000 | 2 |
| LGALS1 | NA | NA | -1.2970 | **0.0057** | **0.0029** | 3 |
| LHX6 | 1.7916 | **1.86E-09** | NA | NA | 0.0000 | 3 |
| LIF | -1.8940 | **0.0171** | -1.3512 | 0.1101 | 0.0000 | 1 |
| LIG3 | -1.3828 | **1.68E-08** | NA | NA | 0.0008 | 3 |
| LIG4 | 1.2104 | **0.0165** | 1.0463 | 0.6925 | **0.0049** | 13 |
| LIMS1 | NA | NA | 1.5418 | **0.0265** | 0.0013 | 10 |
| LIN37 | 1.0770 | 0.4513 | -1.3260 | **0.0016** | 0.0001 | 6 |
| LIN7A | -1.0109 | 0.8735 | -1.2683 | **0.0105** | 0.0011 | 4 |
| LIN7C | 1.0706 | 0.4333 | 1.5490 | **0.0021** | 0.0001 | 2 |
| LMAN1 | -1.6872 | **3.60E-08** | NA | NA | 0.0000 | 1 |
| LMAN2 | -1.4595 | **7.00E-08** | NA | NA | 0.0000 | 1 |
| LMNA | 1.5613 | **9.86E-08** | NA | NA | **0.0082** | **15** |
| LMNB2 | -1.8597 | **2.57E-09** | NA | NA | 0.0005 | 11 |
| LMO1 | NA | NA | -1.5849 | **0.0102** | **0.0078** | **34** |
| LMO4 | -1.3499 | **0.0005** | 1.2606 | 0.1836 | **0.0133** | **40** |
| LMTK2 | NA | NA | 1.1370 | **0.0336** | 0.0001 | 3 |
| LONRF3 | -1.6589 | **1.18E-05** | 1.0886 | 0.6158 | 0.0000 | 2 |
| LOX | 1.2997 | **0.0439** | NA | NA | 0.0008 | 3 |
| LPAR2 | -1.6179 | **7.73E-07** | NA | NA | 0.0000 | 1 |
| LRCH4 | 1.1623 | **0.0182** | NA | NA | 0.0000 | 3 |
| LRP1 | NA | NA | -1.2668 | **0.0179** | **0.0031** | 8 |
| LRP2BP | -1.1611 | 0.0617 | 1.1905 | **0.0445** | 0.0005 | 8 |
| LRP6 | 1.0100 | 0.8823 | 1.2357 | **0.0401** | 0.0018 | 2 |
| LRP8 | -1.4274 | **0.0005** | -1.0031 | 0.9827 | 0.0000 | 1 |
| LRRC59 | -2.0458 | **1.15E-13** | -1.0340 | 0.8528 | 0.0004 | 4 |
| LRRC6 | -1.3689 | **8.61E-06** | NA | NA | 0.0001 | 2 |
| LRRK1 | 1.1432 | **0.0241** | NA | NA | 0.0000 | 1 |
| LSM1 | -1.2935 | **3.90E-05** | 1.0277 | 0.7501 | 0.0002 | 4 |
| LSM12 | -1.3074 | **8.97E-07** | NA | NA | 0.0001 | 2 |
| LSM3 | -1.2747 | **0.0003** | -1.1105 | 0.1867 | 0.0002 | 5 |
| LSM7 | -1.3526 | **1.23E-05** | 1.0385 | 0.6754 | 0.0001 | 4 |
| LSS | -1.2474 | **0.0393** | -1.0311 | 0.8201 | 0.0002 | 2 |
| LUC7L | -1.1405 | **0.0073** | 1.1930 | 0.3927 | 0.0000 | 1 |
| LXN | 3.0519 | **1.53E-08** | -1.0440 | 0.7485 | 0.0013 | 5 |
| LY6E | 1.2900 | **0.0071** | NA | NA | 0.0000 | 1 |
| LYPLA1 | -2.0354 | **2.06E-14** | 1.6389 | **0.0059** | 0.0005 | 2 |
| LYPLA2 | -1.2950 | **1.04E-05** | NA | NA | 0.0000 | 1 |
| LYST | 1.6826 | **2.38E-08** | NA | NA | 0.0000 | 1 |
| LZTR1 | 1.1741 | **0.0088** | -1.1152 | 0.1598 | 0.0000 | 2 |
| MACF1 | 1.4473 | **1.04E-06** | 1.1152 | 0.2528 | 0.0005 | 3 |
| MAD2L1BP | -1.2719 | **0.0007** | NA | NA | 0.0000 | 3 |
| MADD | 1.1641 | **0.0002** | -1.0622 | 0.3452 | 0.0000 | 1 |
| MAF | 1.9969 | **6.93E-09** | -1.2179 | 0.1811 | 0.0000 | 2 |
| MAFG | 1.6013 | **8.73E-12** | -1.0394 | 0.6983 | 0.0001 | 4 |
| MAGED1 | -1.4082 | **0.0010** | -1.1180 | 0.4297 | **0.0053** | **27** |
| MAGI1 | -1.8063 | **4.71E-08** | 1.1393 | 0.3499 | 0.0000 | 1 |
| MAGOH | -1.2942 | **5.78E-06** | NA | NA | 0.0009 | 7 |
| MAGOHB | -1.2512 | **0.0005** | NA | NA | **0.0045** | **25** |
| MAN2B2 | 1.2236 | **0.0050** | 1.1354 | 0.4039 | 0.0000 | 1 |
| MAP1A | 1.6496 | **2.59E-09** | NA | NA | 0.0000 | 1 |
| MAP1B | 1.3252 | **0.0035** | 1.1015 | 0.5476 | 0.0003 | 4 |
| MAP1LC3C | NA | NA | -1.2137 | **0.0333** | 0.0005 | 4 |
| MAP2 | 1.5845 | **0.0035** | -1.0205 | 0.8377 | 0.0000 | 2 |
| MAP2K1 | 1.0051 | 0.9437 | -1.1325 | **0.0489** | 0.0012 | 5 |
| MAP2K5 | 1.3659 | **2.20E-07** | 1.0430 | 0.7107 | 0.0001 | 2 |
| MAP3K1 | -1.9640 | **1.83E-11** | 1.1999 | 0.0616 | 0.0014 | 3 |
| MAP3K10 | -1.0109 | 0.8414 | -1.1871 | **0.0162** | 0.0024 | 2 |
| MAP3K11 | 1.1679 | **0.0162** | -1.0239 | 0.8449 | 0.0000 | 1 |
| MAP3K14 | 1.3301 | **0.0007** | -1.0348 | 0.7778 | 0.0000 | 1 |
| MAP3K3 | NA | NA | -1.2330 | **0.0271** | 0.0002 | 3 |
| MAP3K4 | -1.4434 | **2.80E-06** | 1.0100 | 0.9183 | 0.0001 | 2 |
| MAP3K8 | 1.9397 | **0.0001** | -1.0760 | 0.7075 | 0.0000 | 2 |
| MAPK1 | -1.3152 | **3.49E-06** | NA | NA | **0.0045** | **15** |
| MAPK10 | 1.3495 | **0.0104** | -1.2058 | 0.1402 | 0.0001 | 3 |
| MAPK11 | 1.5820 | **2.13E-09** | -1.1345 | 0.3258 | 0.0000 | 1 |
| MAPK14 | -1.2706 | **0.0019** | -1.0131 | 0.9038 | **0.0041** | 11 |
| MAPK1IP1L | -1.2918 | **9.54E-06** | 1.0568 | 0.4668 | 0.0008 | 9 |
| MAPK3 | 1.3437 | **8.85E-07** | -1.2087 | **0.0111** | 0.0011 | 10 |
| MAPK6 | -1.9944 | **2.18E-08** | NA | NA | 0.0008 | 4 |
| MAPK8IP2 | NA | NA | -1.4128 | **0.0051** | 0.0012 | 4 |
| MAPKAPK5 | -1.2588 | **3.83E-06** | 1.2599 | 0.0648 | 0.0000 | 1 |
| MAPKBP1 | 1.2443 | **0.0016** | 1.0525 | 0.6326 | 0.0020 | 12 |
| MAPRE1 | -1.1444 | **0.0084** | -1.0738 | 0.2406 | **0.0048** | 12 |
| MAPRE3 | 1.2630 | **0.0013** | 1.0038 | 0.9651 | 0.0017 | 13 |
| MARCH2 | 1.3025 | **0.0020** | NA | NA | 0.0014 | 8 |
| MARCH5 | -1.2848 | **4.04E-06** | 1.0237 | 0.7820 | 0.0001 | 6 |
| MARCH6 | -1.0003 | 0.9967 | 1.3615 | **0.0407** | 0.0000 | 1 |
| MARCH8 | 1.4803 | **7.11E-08** | 1.0187 | 0.8177 | 0.0000 | 2 |
| MARK1 | -2.0307 | **7.14E-11** | 1.0895 | 0.5177 | 0.0000 | 1 |
| MARK3 | -1.2131 | **0.0003** | 1.0523 | 0.6946 | 0.0002 | 3 |
| MARS | -1.1811 | **0.0248** | -1.0998 | 0.2667 | 0.0000 | 1 |
| MATN2 | 4.3542 | **1.79E-11** | 1.0347 | 0.9010 | 0.0000 | 1 |
| MATN3 | NA | NA | -1.1463 | **0.0171** | 0.0000 | 1 |
| MAVS | -1.1336 | **0.0341** | 1.1199 | 0.4914 | 0.0000 | 1 |
| MAX | 1.2609 | **0.0001** | 1.0228 | 0.7837 | 0.0017 | 9 |
| MBD1 | 1.0315 | 0.5255 | -1.2207 | **0.0075** | 0.0000 | 3 |
| MBD2 | -1.2407 | **0.0022** | -1.1064 | 0.1316 | 0.0008 | 3 |
| MBD4 | -1.2300 | **0.0057** | NA | NA | 0.0003 | 4 |
| MBNL1 | 1.7547 | **1.87E-09** | 1.2855 | 0.0791 | 0.0004 | 3 |
| MBNL3 | NA | NA | 1.2255 | **0.0330** | 0.0000 | 1 |
| MBP | 1.2934 | **0.0338** | NA | NA | 0.0000 | 1 |
| MCFD2 | 1.3963 | **3.37E-05** | 1.1743 | 0.0644 | 0.0008 | 2 |
| MCM2 | -1.9600 | **3.90E-08** | NA | NA | 0.0009 | 8 |
| MCM3 | -1.4641 | **1.35E-06** | 1.1005 | 0.3499 | 0.0006 | 6 |
| MCM5 | -1.3039 | **0.0009** | NA | NA | 0.0007 | 5 |
| MCM7 | -1.6101 | **4.18E-07** | NA | NA | **0.0056** | **14** |
| MCPH1 | -1.1602 | **0.0003** | 1.0528 | 0.4664 | 0.0000 | 2 |
| MCRS1 | -1.1935 | **0.0016** | -1.0372 | 0.6689 | **0.0059** | **27** |
| MCTS1 | -1.4604 | **1.36E-07** | -1.1066 | 0.1754 | 0.0000 | 1 |
| MDFIC | 2.3560 | **2.59E-09** | -1.0319 | 0.8224 | 0.0000 | 1 |
| MDH2 | -1.3230 | **2.54E-06** | -1.0344 | 0.8068 | 0.0002 | 2 |
| MDK | -1.6675 | **0.0003** | -1.1354 | 0.3258 | 0.0001 | 3 |
| MDM2 | -1.1983 | **0.0064** | -1.0095 | 0.9183 | **0.0124** | **23** |
| MDM4 | -1.5816 | **1.51E-08** | 1.1060 | 0.3864 | 0.0004 | 3 |
| MEAF6 | -1.1295 | **0.0133** | 1.0793 | 0.2831 | 0.0002 | 5 |
| MECOM | -3.9361 | **1.08E-15** | 1.3968 | **0.0139** | 0.0000 | 1 |
| MECP2 | 1.3591 | **6.20E-09** | NA | NA | 0.0000 | 2 |
| MED1 | -1.1388 | **0.0186** | 1.3546 | **0.0312** | 0.0001 | 5 |
| MED12 | 1.1446 | **0.0137** | NA | NA | 0.0000 | 1 |
| MED14 | -1.1763 | **0.0047** | 1.1460 | 0.2098 | 0.0000 | 1 |
| MED18 | -1.1866 | **0.0146** | -1.0182 | 0.9131 | 0.0000 | 1 |
| MED20 | -1.4474 | **6.19E-09** | 1.0773 | 0.2929 | 0.0008 | 4 |
| MED31 | -1.2488 | **0.0018** | -1.0501 | 0.6181 | 0.0006 | 2 |
| MED7 | -1.1906 | **0.0104** | -1.0159 | 0.8980 | 0.0015 | 3 |
| MEF2A | 1.2771 | **0.0028** | 1.5005 | **0.0021** | 0.0000 | 1 |
| MEF2C | 3.5394 | **2.98E-21** | -1.0938 | 0.4243 | 0.0000 | 1 |
| MEF2D | 1.2108 | **0.0011** | -1.2671 | 0.1516 | 0.0001 | 2 |
| MEGF8 | 1.1584 | **0.0434** | NA | NA | 0.0000 | 1 |
| MEN1 | -1.2791 | **1.49E-05** | -1.0174 | 0.8675 | 0.0000 | 1 |
| MEST | -1.5185 | **0.0254** | 1.1735 | 0.3175 | 0.0009 | 3 |
| METTL1 | -1.1290 | **0.0305** | NA | NA | 0.0000 | 1 |
| METTL17 | -1.1713 | **0.0005** | -1.0988 | 0.4101 | 0.0004 | 7 |
| MFSD6 | -1.8951 | **1.96E-06** | 1.4328 | 0.0888 | 0.0011 | 12 |
| MGRN1 | -1.1498 | **0.0051** | -1.0221 | 0.7987 | 0.0000 | 2 |
| MGST2 | -1.2991 | **0.0003** | -1.1112 | 0.3851 | 0.0005 | 5 |
| MGST3 | 1.4061 | **1.98E-08** | -1.0937 | 0.2418 | **0.0062** | **25** |
| MIA2 | -1.6650 | **3.50E-08** | 1.2231 | **0.0287** | 0.0000 | 2 |
| MICAL1 | -1.3709 | **0.0006** | -1.0431 | 0.7919 | 0.0000 | 2 |
| MICALL2 | -1.2976 | **0.0042** | 1.0487 | 0.7350 | 0.0000 | 1 |
| MIEF1 | -1.1548 | **0.0029** | NA | NA | 0.0017 | 7 |
| MIIP | 1.2737 | **2.16E-05** | NA | NA | 0.0011 | 6 |
| MINPP1 | -1.0671 | 0.5332 | 1.4290 | **0.0234** | 0.0000 | 1 |
| MIS18A | -1.4822 | **3.73E-06** | 1.0205 | 0.8975 | 0.0008 | 4 |
| MITF | -1.9811 | **2.00E-06** | -1.1006 | 0.4542 | 0.0024 | 5 |
| MKLN1 | -1.2645 | **0.0007** | 1.0836 | 0.5443 | 0.0000 | 1 |
| MKNK1 | 1.4032 | **5.43E-06** | -1.1061 | 0.2486 | 0.0001 | 3 |
| MKRN1 | -1.1864 | **0.0005** | -1.0606 | 0.3973 | 0.0001 | 4 |
| MLH1 | -1.3075 | **1.22E-05** | 1.0354 | 0.6660 | **0.0028** | **16** |
| MLH3 | -1.1251 | **0.0388** | 1.1321 | 0.1493 | 0.0000 | 1 |
| MLLT3 | -1.3650 | **0.0039** | 1.2292 | **0.0308** | 0.0008 | 3 |
| MLN | NA | NA | -1.2467 | **0.0206** | 0.0002 | 2 |
| MLX | -1.1559 | **0.0188** | 1.0612 | 0.5845 | 0.0019 | 5 |
| MLXIP | 1.4097 | **1.20E-06** | -1.1415 | 0.2011 | 0.0000 | 2 |
| MMP2 | 1.5984 | **0.0001** | NA | NA | 0.0018 | 4 |
| MNAT1 | -1.3044 | **0.0005** | NA | NA | 0.0001 | 4 |
| MNS1 | -1.3784 | **0.0353** | NA | NA | 0.0003 | 7 |
| MOAP1 | -1.7390 | **1.76E-10** | 1.2567 | 0.1768 | 0.0001 | 4 |
| MOB1A | -1.2839 | **0.0011** | 1.3987 | **0.0022** | 0.0001 | 4 |
| MORC3 | 1.4682 | **0.0007** | NA | NA | 0.0000 | 1 |
| MOS | NA | NA | -1.5993 | **0.0183** | **0.0029** | **14** |
| MOXD1 | -1.6253 | **0.0248** | -1.2097 | 0.1695 | 0.0000 | 1 |
| MPDZ | 2.7640 | **4.57E-18** | 1.0732 | 0.5057 | 0.0000 | 1 |
| MPHOSPH8 | 1.3495 | **0.0001** | 1.2372 | 0.1473 | 0.0000 | 4 |
| MPP5 | -1.1847 | **0.0095** | 1.3640 | **0.0032** | 0.0001 | 3 |
| MPPED2 | -2.2349 | **0.0001** | 1.4921 | 0.0683 | 0.0000 | 2 |
| MPRIP | 1.1557 | **0.0058** | NA | NA | 0.0000 | 1 |
| MPST | -1.3072 | **0.0001** | NA | NA | 0.0000 | 1 |
| MRNIP | 1.4512 | **4.60E-07** | NA | NA | 0.0000 | 2 |
| MRPL12 | -1.3989 | **0.0001** | 1.1526 | 0.4907 | 0.0010 | 8 |
| MRPL15 | -1.6626 | **2.96E-10** | 1.0137 | 0.8811 | 0.0000 | 2 |
| MRPL17 | -1.1523 | **0.0353** | NA | NA | 0.0000 | 1 |
| MRPL28 | -1.1472 | **0.0095** | NA | NA | 0.0000 | 4 |
| MRPL44 | -1.2038 | **0.0057** | 1.1722 | 0.1263 | 0.0001 | 3 |
| MRPS22 | -1.1737 | **0.0011** | NA | NA | 0.0000 | 1 |
| MSC | NA | NA | -1.2558 | **0.0088** | 0.0004 | 5 |
| MSH2 | -1.4146 | **0.0015** | NA | NA | 0.0002 | 4 |
| MSL1 | -1.1965 | **0.0051** | NA | NA | **1.0000** | 2 |
| MSL2 | 1.1872 | 0.0503 | 1.3518 | **0.0021** | 0.0000 | 1 |
| MSN | 1.4024 | **0.0002** | -1.1623 | 0.1285 | 0.0006 | 2 |
| MTA1 | -1.1264 | **0.0128** | -1.0110 | 0.9131 | 0.0001 | 4 |
| MTAP | -1.2000 | **0.0001** | 1.1280 | 0.0843 | 0.0000 | 1 |
| MTCH2 | -1.2760 | **9.94E-06** | -1.0413 | 0.6387 | 0.0000 | 1 |
| MTDH | -1.1281 | 0.0849 | 1.3618 | **0.0161** | 0.0000 | 1 |
| MTF2 | 1.0365 | 0.6339 | 1.3100 | **0.0161** | 0.0000 | 1 |
| MTG1 | -1.3396 | **0.0010** | 1.0132 | 0.9013 | 0.0002 | 2 |
| MTHFD2 | -1.5777 | **0.0003** | -1.0622 | 0.8066 | 0.0000 | 1 |
| MTMR6 | 1.2215 | **0.0052** | NA | NA | 0.0000 | 3 |
| MTOR | -1.5895 | **2.50E-09** | NA | NA | 0.0015 | 6 |
| MTX2 | -1.1614 | **0.0071** | 1.0631 | 0.5057 | 0.0000 | 2 |
| MUC1 | -4.0817 | **1.53E-10** | 1.3567 | 0.2018 | **0.0076** | **24** |
| MUSK | NA | NA | -1.2732 | **0.0056** | 0.0001 | 3 |
| MVD | -1.1526 | **0.0128** | NA | NA | 0.0000 | 1 |
| MX1 | 1.9316 | **1.49E-06** | NA | NA | 0.0017 | 6 |
| MXD3 | -1.2440 | **0.0013** | 1.0230 | 0.8506 | 0.0002 | 3 |
| MXI1 | 1.6866 | **4.10E-07** | NA | NA | 0.0001 | 3 |
| MXRA8 | 1.3771 | **0.0011** | NA | NA | 0.0000 | 3 |
| MYB | -1.7181 | **0.0001** | 1.3890 | 0.0568 | 0.0022 | 6 |
| MYCBP | -1.9238 | **8.73E-13** | 1.2190 | **0.0351** | 0.0000 | 1 |
| MYCBP2 | 1.5779 | **7.11E-09** | NA | NA | 0.0000 | 1 |
| MYCL | -1.2299 | **0.0202** | NA | NA | 0.0000 | 1 |
| MYD88 | -1.2004 | **0.0103** | -1.0301 | 0.7844 | 0.0018 | 8 |
| MYDGF | -1.3750 | **9.17E-06** | NA | NA | 0.0001 | 3 |
| MYH10 | 2.1608 | **3.30E-10** | NA | NA | 0.0000 | 1 |
| MYH9 | 1.2110 | **0.0113** | NA | NA | 0.0000 | 2 |
| MYL12B | -1.1430 | **0.0080** | NA | NA | 0.0000 | 1 |
| MYLIP | -1.6528 | **4.01E-08** | -1.0613 | 0.6689 | **0.0027** | 10 |
| MYO15B | 1.2088 | **0.0128** | NA | NA | **0.0101** | **34** |
| MYO6 | -2.2332 | **3.68E-14** | NA | NA | 0.0012 | 5 |
| MYOG | NA | NA | -1.5606 | **0.0337** | 0.0012 | 9 |
| MYOM1 | 2.1208 | **7.46E-09** | -1.0714 | 0.5441 | 0.0002 | 4 |
| MYOM2 | -1.0543 | 0.5966 | -1.3416 | **0.0010** | 0.0000 | 2 |
| MYOZ1 | 1.5180 | **0.0035** | 1.0430 | 0.6709 | 0.0018 | 13 |
| MYOZ2 | 1.3034 | **0.0253** | NA | NA | 0.0006 | 4 |
| MZF1 | -1.1321 | **0.0286** | -1.1164 | 0.2186 | 0.0000 | 1 |
| N4BP2L2 | 1.3257 | **0.0029** | NA | NA | 0.0000 | 2 |
| NAA15 | -1.2541 | **0.0151** | NA | NA | 0.0000 | 1 |
| NAA50 | -1.8196 | **1.29E-08** | NA | NA | 0.0000 | 1 |
| NADK | -1.1477 | **0.0379** | NA | NA | 0.0000 | 1 |
| NADSYN1 | -1.1347 | **0.0350** | -1.1059 | 0.2139 | 0.0002 | 6 |
| NAGK | 1.1592 | **0.0163** | NA | NA | 0.0011 | 8 |
| NAP1L1 | 1.4469 | **0.0004** | NA | NA | 0.0002 | 3 |
| NAP1L2 | 3.2901 | **3.89E-11** | 1.0327 | 0.8379 | 0.0011 | 4 |
| NAPA | -1.1742 | **0.0160** | -1.0618 | 0.5653 | 0.0013 | 4 |
| NAPG | -1.5205 | **2.56E-07** | 1.2181 | **0.0341** | 0.0000 | 1 |
| NARF | -1.2237 | **0.0002** | 1.0115 | 0.9287 | 0.0000 | 1 |
| NASP | -1.3218 | **0.0011** | 1.2333 | 0.1031 | 0.0000 | 5 |
| NAT8 | NA | NA | -1.4721 | **0.0332** | 0.0000 | 5 |
| NAT9 | NA | NA | -1.1414 | **0.0358** | 0.0000 | 1 |
| NAXD | 1.1795 | **0.0074** | NA | NA | 0.0000 | 1 |
| NCBP1 | -1.5086 | **1.44E-05** | 1.0791 | 0.4870 | 0.0008 | 4 |
| NCBP2 | -1.1937 | **0.0003** | -1.0448 | 0.5476 | 0.0000 | 1 |
| NCK1 | 1.3087 | **0.0014** | NA | NA | 0.0023 | **16** |
| NCK2 | 1.2189 | **0.0002** | 1.0788 | 0.4203 | **0.0189** | **36** |
| NCKIPSD | 1.2129 | **0.0075** | 1.1695 | 0.1372 | 0.0006 | 10 |
| NCOA6 | -1.1827 | **0.0031** | -1.0740 | 0.4497 | 0.0008 | 9 |
| NCOR1 | -1.1757 | **0.0017** | 1.3254 | **0.0359** | **0.0027** | **15** |
| NCOR2 | 1.1567 | **0.0061** | 1.0335 | 0.7820 | **0.0033** | **15** |
| NCR1 | -1.7437 | **0.0017** | -1.1621 | 0.2929 | 0.0000 | 2 |
| NCS1 | 1.2665 | **0.0031** | NA | NA | 0.0007 | 6 |
| NDN | 1.4879 | **1.28E-06** | NA | NA | 0.0025 | 8 |
| NDUFA13 | -1.2875 | **0.0001** | 1.1111 | 0.2674 | 0.0000 | 1 |
| NDUFAB1 | -1.1981 | **0.0005** | -1.1739 | **0.0379** | 0.0006 | 8 |
| NDUFAF1 | -1.2262 | **0.0008** | -1.0922 | 0.2081 | 0.0000 | 3 |
| NDUFB11 | -1.4322 | **4.87E-09** | -1.1153 | 0.3676 | 0.0000 | 2 |
| NDUFB2 | -1.3206 | **2.01E-06** | -1.0382 | 0.5283 | 0.0000 | 1 |
| NDUFB5 | 1.1288 | **0.0305** | 1.2965 | **0.0078** | 0.0000 | 1 |
| NDUFB6 | -1.1570 | **0.0035** | -1.1408 | **0.0283** | 0.0000 | 1 |
| NDUFS1 | -1.4270 | **6.22E-07** | 1.2214 | **0.0191** | 0.0000 | 1 |
| NDUFS3 | -1.2055 | **0.0005** | NA | NA | 0.0000 | 1 |
| NDUFV1 | -1.1463 | **0.0031** | -1.0006 | 0.9965 | 0.0000 | 1 |
| NECAB2 | 1.2349 | 0.1985 | -1.5682 | **0.0062** | **0.0030** | **20** |
| NEDD4 | 1.5249 | **2.69E-05** | -1.0031 | 0.9796 | **0.0033** | 9 |
| NEDD9 | -1.2273 | 0.1585 | 1.5657 | **0.0229** | **0.0044** | **16** |
| NEIL1 | -1.1132 | 0.0659 | -1.2465 | **0.0250** | 0.0000 | 1 |
| NEK1 | 1.2546 | **0.0137** | NA | NA | 0.0000 | 2 |
| NELFA | -1.1197 | **0.0220** | NA | NA | 0.0009 | 5 |
| NELFCD | -1.4452 | **1.88E-13** | NA | NA | 0.0000 | 2 |
| NELFE | -1.1780 | **4.34E-05** | NA | NA | 0.0000 | 2 |
| NET1 | 1.7799 | **7.33E-07** | NA | NA | 0.0017 | 2 |
| NEUROG2 | NA | NA | -1.1545 | **0.0478** | 0.0000 | 1 |
| NF2 | -1.1187 | **0.0211** | NA | NA | **0.0034** | 11 |
| NFATC1 | 1.5823 | **1.06E-06** | 1.0053 | 0.9733 | 0.0000 | 1 |
| NFATC2IP | -1.3038 | **1.06E-06** | 1.0461 | 0.4636 | 0.0000 | 1 |
| NFE2L1 | 1.1273 | **0.0217** | -1.0234 | 0.8506 | 0.0000 | 2 |
| NFE2L2 | 1.3321 | **0.0022** | NA | NA | 0.0001 | 3 |
| NFKB1 | 1.0573 | 0.2302 | -1.1457 | **0.0386** | **0.0060** | 13 |
| NFKB2 | 1.3309 | **0.0001** | -1.1534 | 0.4084 | 0.0000 | 2 |
| NFKBIA | 1.5700 | **5.13E-06** | -1.0357 | 0.8117 | **0.0036** | 8 |
| NFKBIE | 1.2679 | **0.0132** | 1.1749 | 0.1402 | 0.0008 | 3 |
| NFYA | -1.5432 | **3.96E-13** | NA | NA | 0.0005 | 6 |
| NGB | NA | NA | -1.1922 | **0.0348** | 0.0000 | 2 |
| NGF | 2.8848 | **8.71E-11** | -1.5129 | 0.1186 | 0.0001 | 3 |
| NGFR | 1.5002 | **0.0143** | NA | NA | 0.0006 | 5 |
| NHP2 | -1.5237 | **2.53E-10** | NA | NA | 0.0008 | 2 |
| NIF3L1 | -1.2477 | **0.0001** | 1.0720 | 0.3475 | **0.0062** | **32** |
| NINJ2 | 1.5436 | **0.0002** | 1.0905 | 0.4325 | 0.0018 | 13 |
| NIP7 | NA | NA | 1.3007 | **0.0401** | 0.0000 | 2 |
| NIPAL3 | -1.1717 | **0.0217** | 1.1445 | 0.1312 | 0.0000 | 1 |
| NIPSNAP1 | -1.2115 | **0.0048** | NA | NA | 0.0000 | 1 |
| NKG7 | -3.2484 | **2.02E-08** | -1.2332 | 0.3194 | 0.0021 | 10 |
| NKX3-1 | 1.3071 | **0.0101** | -1.3344 | 0.3346 | 0.0005 | 2 |
| NLK | -1.3216 | **0.0005** | 1.3996 | **0.0028** | 0.0000 | 1 |
| NMI | 1.3145 | **0.0001** | 1.0547 | 0.7546 | 0.0025 | 10 |
| NOC4L | -1.0666 | 0.2599 | -1.3686 | **0.0469** | 0.0002 | 4 |
| NOL12 | -1.1862 | **0.0338** | NA | NA | 0.0001 | 3 |
| NOL6 | -1.1977 | **0.0056** | NA | NA | 0.0000 | 1 |
| NOLC1 | -1.2999 | **2.69E-06** | NA | NA | 0.0000 | 1 |
| NONO | -1.1626 | **0.0479** | NA | NA | 0.0019 | 6 |
| NOP10 | -1.2084 | **0.0075** | -1.0072 | 0.9316 | 0.0000 | 1 |
| NOP2 | -1.2510 | **0.0001** | 1.0310 | 0.7936 | 0.0001 | 3 |
| NOS1AP | -1.1216 | 0.2075 | -1.2362 | **0.0096** | 0.0000 | 1 |
| NOSIP | -1.2136 | **0.0020** | NA | NA | 0.0000 | 1 |
| NOTCH2 | 1.2466 | **0.0010** | 1.0463 | 0.5567 | 0.0000 | 1 |
| NOTCH4 | 1.7235 | **1.77E-10** | -1.2386 | **0.0032** | 0.0000 | 1 |
| NOX1 | NA | NA | -1.1520 | **0.0113** | 0.0000 | 1 |
| NPEPL1 | 1.2234 | **0.0005** | 1.0136 | 0.9043 | 0.0000 | 1 |
| NPHP1 | -1.2844 | **0.0020** | 1.0127 | 0.9066 | 0.0002 | 2 |
| NPPA | 1.1811 | 0.0778 | -1.1463 | **0.0389** | 0.0000 | 1 |
| NQO2 | 1.6488 | **2.59E-08** | NA | NA | 0.0000 | 2 |
| NR2C1 | -1.2218 | **0.0069** | NA | NA | 0.0000 | 1 |
| NR3C1 | 3.2310 | **2.09E-22** | NA | NA | **0.0056** | **14** |
| NR4A1 | 1.6692 | **0.0132** | NA | NA | 0.0004 | 5 |
| NR4A3 | 1.8767 | **0.0047** | -1.3212 | 0.1562 | 0.0003 | 5 |
| NRAS | -1.7212 | **2.47E-10** | 1.1085 | 0.2551 | 0.0000 | 2 |
| NRG1 | NA | NA | -1.4350 | **0.0443** | 0.0000 | 2 |
| NRIP2 | 1.4526 | **0.0001** | NA | NA | 0.0000 | 1 |
| NSD1 | -1.1691 | **0.0025** | -1.0486 | 0.5829 | 0.0002 | 5 |
| NSD2 | -2.1471 | **2.54E-15** | NA | NA | 0.0000 | 1 |
| NSD3 | -1.1295 | **0.0168** | NA | NA | 0.0001 | 3 |
| NSDHL | -1.4553 | **2.02E-05** | NA | NA | 0.0000 | 1 |
| NSF | -1.2543 | **0.0006** | -1.0361 | 0.6136 | 0.0000 | 1 |
| NTRK1 | -1.0648 | 0.7735 | -1.2090 | **0.0189** | 0.0015 | 7 |
| NTRK2 | 3.6495 | **9.08E-11** | -1.0040 | 0.9754 | 0.0001 | 2 |
| NUCB1 | 1.3934 | **1.83E-08** | NA | NA | 0.0000 | 1 |
| NUDC | -1.1381 | **0.0038** | 1.0551 | 0.6898 | 0.0000 | 2 |
| NUDT15 | -2.0012 | **2.73E-12** | 1.1780 | 0.1618 | 0.0000 | 1 |
| NUDT21 | -1.5979 | **6.77E-09** | 1.0563 | 0.6081 | **0.0034** | 11 |
| NUMA1 | -1.1241 | 0.2194 | 1.4230 | **0.0094** | 0.0009 | 3 |
| NUMB | 1.2235 | **0.0013** | -1.0394 | 0.7524 | 0.0005 | 4 |
| NUP153 | -1.4494 | **0.0006** | 1.0363 | 0.8726 | 0.0000 | 1 |
| NUP155 | -1.1732 | **0.0416** | 1.0115 | 0.9301 | 0.0001 | 2 |
| NUP160 | -1.4446 | **2.38E-06** | -1.0098 | 0.9360 | 0.0000 | 1 |
| NUP188 | -1.2129 | **0.0063** | -1.1861 | 0.0754 | 0.0000 | 1 |
| NUP205 | -1.3095 | **0.0001** | 1.0502 | 0.5730 | 0.0000 | 1 |
| NUP50 | -1.2919 | **0.0010** | 1.1831 | 0.0704 | 0.0001 | 3 |
| NUP62 | -1.1989 | **0.0109** | 1.0264 | 0.8095 | **0.0112** | **28** |
| NUP85 | -1.2799 | **3.92E-06** | -1.0026 | 0.9785 | 0.0000 | 1 |
| NUP88 | -1.3291 | **1.65E-07** | 1.0321 | 0.7542 | 0.0000 | 2 |
| NUP93 | -1.2389 | **0.0010** | NA | NA | 0.0008 | 3 |
| NUP98 | -1.5313 | **6.97E-09** | 1.0961 | 0.3450 | 0.0008 | 2 |
| NVL | -1.1527 | **0.0061** | 1.0448 | 0.5868 | 0.0000 | 2 |
| NXT2 | -1.2981 | **0.0011** | NA | NA | 0.0001 | 5 |
| OARD1 | 1.1510 | **0.0457** | NA | NA | 0.0000 | 1 |
| OAS1 | 1.6841 | **3.43E-06** | -1.0829 | 0.5068 | 0.0002 | 3 |
| ODC1 | -2.0257 | **8.14E-14** | NA | NA | 0.0000 | 1 |
| ODF2 | -1.5839 | **3.77E-12** | NA | NA | 0.0000 | 1 |
| OFD1 | -1.5091 | **0.0001** | NA | NA | 0.0000 | 1 |
| OGFOD1 | -1.6298 | **3.13E-07** | 1.2312 | 0.1041 | 0.0008 | 2 |
| OLA1 | -1.3664 | **0.0001** | 1.2360 | **0.0331** | 0.0000 | 1 |
| OPLAH | -1.8714 | **6.81E-12** | 1.1804 | 0.1683 | 0.0008 | 2 |
| OPTN | 2.5875 | **1.19E-16** | -1.0010 | 0.9954 | **0.0094** | **17** |
| ORC2 | -1.1942 | **0.0017** | 1.2037 | **0.0139** | 0.0000 | 3 |
| ORMDL2 | -1.9027 | **2.10E-11** | 1.3775 | **0.0381** | 0.0000 | 6 |
| OSBPL3 | -1.3035 | **0.0093** | NA | NA | 0.0004 | 5 |
| OSGIN1 | 1.4923 | **0.0001** | -1.1870 | 0.2062 | 0.0011 | 12 |
| OSMR | 1.5461 | **0.0019** | -1.2469 | 0.0545 | 0.0000 | 1 |
| OSTF1 | -1.2241 | **0.0067** | 1.1172 | 0.2944 | 0.0007 | 10 |
| OTUB1 | -1.1661 | **0.0045** | 1.1338 | 0.4208 | 0.0000 | 3 |
| P2RY1 | 1.2455 | 0.0672 | 1.3646 | **0.0258** | 0.0000 | 1 |
| P4HB | -1.5678 | **2.55E-08** | NA | NA | 0.0000 | 1 |
| PA2G4 | -1.1512 | **0.0019** | NA | NA | 0.0000 | 1 |
| PABPC4 | 1.1119 | **0.0295** | 1.1552 | 0.3068 | 0.0000 | 1 |
| PACSIN2 | NA | NA | 1.3148 | **0.0209** | 0.0000 | 1 |
| PAEP | -15.7812 | **1.99E-07** | NA | NA | 0.0000 | 1 |
| PAFAH1B1 | -1.1571 | **0.0046** | NA | NA | 0.0005 | 3 |
| PAFAH1B2 | -1.1973 | **0.0044** | 1.1882 | 0.0805 | 0.0001 | 3 |
| PAFAH1B3 | -1.3401 | **0.0012** | NA | NA | 0.0016 | 4 |
| PAICS | -1.9706 | **1.29E-11** | 1.0399 | 0.7393 | 0.0022 | 6 |
| PAIP1 | -1.3610 | **3.99E-05** | 1.2178 | **0.0258** | 0.0000 | 1 |
| PAK1IP1 | -1.4218 | **0.0001** | 1.2592 | 0.0572 | 0.0000 | 1 |
| PAK2 | -1.1552 | **0.0263** | 1.0509 | 0.5221 | 0.0020 | 9 |
| PALMD | 1.5969 | **0.0001** | 1.0721 | 0.6130 | 0.0000 | 1 |
| PAM | -1.0229 | 0.8510 | 1.2523 | **0.0310** | 0.0000 | 1 |
| PARK7 | -1.1693 | **0.0018** | -1.0559 | 0.4861 | 0.0013 | 3 |
| PARP1 | -1.5446 | **4.72E-11** | -1.0105 | 0.9165 | **0.0027** | 7 |
| PARP16 | 1.1073 | **0.0264** | -1.0799 | 0.3440 | 0.0000 | 1 |
| PARP2 | -1.1729 | **0.0052** | -1.0017 | 0.9828 | 0.0000 | 1 |
| PARVA | 1.3387 | **3.43E-05** | -1.0189 | 0.8705 | 0.0000 | 1 |
| PASK | -1.5635 | **2.19E-07** | 1.0403 | 0.6228 | 0.0000 | 1 |
| PATZ1 | -1.1801 | **0.0106** | -1.0001 | 0.9988 | 0.0019 | **15** |
| PAX5 | NA | NA | -1.3859 | **0.0206** | **0.0042** | 13 |
| PAXIP1 | -1.2745 | **0.0001** | 1.1021 | 0.2191 | **0.0027** | 6 |
| PBLD | -1.3744 | **0.0002** | NA | NA | 0.0000 | 3 |
| PBX2 | 1.1866 | **0.0033** | -1.0383 | 0.7606 | 0.0000 | 4 |
| PBXIP1 | 1.6483 | **6.20E-06** | NA | NA | 0.0010 | 9 |
| PCBD1 | -1.3920 | **1.64E-05** | -1.0085 | 0.9505 | 0.0024 | 8 |
| PCBP1 | -1.2403 | **1.51E-06** | -1.0896 | 0.2991 | 0.0007 | 7 |
| PCBP2 | -1.1120 | **0.0179** | 1.2521 | 0.0607 | 0.0004 | 5 |
| PCBP3 | -1.2942 | **0.0023** | -1.1653 | 0.0647 | 0.0009 | 3 |
| PCGF2 | 1.1501 | **0.0293** | -1.4961 | **0.0153** | 0.0008 | 6 |
| PCGF3 | -1.1754 | **0.0067** | -1.0312 | 0.7976 | 0.0008 | 3 |
| PCK2 | -1.3386 | **0.0033** | 1.0947 | 0.2690 | 0.0000 | 1 |
| PCLAF | -5.8761 | **2.34E-10** | NA | NA | 0.0000 | 2 |
| PCMTD2 | 1.2609 | **0.0194** | NA | NA | 0.0000 | 1 |
| PCNA | -1.9479 | **2.63E-07** | 1.1080 | 0.5044 | **0.0075** | **23** |
| PCNT | -1.1934 | **0.0004** | -1.0776 | 0.3324 | 0.0000 | 1 |
| PDAP1 | -1.0753 | 0.3931 | 1.3177 | **0.0093** | 0.0000 | 1 |
| PDCD11 | -1.2427 | **1.82E-06** | NA | NA | 0.0000 | 1 |
| PDCD6 | -1.3240 | **1.02E-10** | -1.1092 | 0.0896 | 0.0005 | 6 |
| PDCD6IP | -1.1318 | **0.0082** | 1.0619 | 0.4262 | 0.0011 | 7 |
| PDCL | -1.3142 | **3.27E-07** | 1.1458 | 0.1676 | 0.0001 | 3 |
| PDE2A | 2.9801 | **9.62E-15** | -1.0125 | 0.9454 | 0.0000 | 1 |
| PDE4DIP | 1.3212 | **0.0050** | 1.1351 | 0.1788 | **0.0036** | **21** |
| PDGFA | 2.1859 | **8.84E-11** | 1.0471 | 0.7966 | 0.0002 | 2 |
| PDGFB | 1.4001 | **0.0004** | -1.4312 | **0.0034** | 0.0008 | 2 |
| PDGFRA | 1.4317 | **0.0036** | -1.1392 | 0.3802 | 0.0020 | 6 |
| PDHX | -1.2685 | **0.0016** | NA | NA | 0.0019 | 4 |
| PDIA3 | -1.4105 | **0.0004** | 1.6595 | **0.0010** | 0.0003 | 3 |
| PDIA4 | -1.8566 | **3.22E-13** | NA | NA | 0.0000 | 1 |
| PDIA6 | -2.0873 | **5.23E-18** | NA | NA | 0.0002 | 2 |
| PDK4 | 2.6790 | **9.52E-07** | NA | NA | 0.0000 | 1 |
| PDLIM7 | NA | NA | -1.4453 | **0.0054** | 0.0015 | 12 |
| PDPK1 | -1.0082 | 0.8790 | 1.2934 | **0.0382** | 0.0013 | 7 |
| PDZD7 | 1.1701 | **0.0318** | -1.1439 | 0.4882 | 0.0000 | 1 |
| PEA15 | 1.3149 | **0.0003** | -1.0559 | 0.5450 | 0.0000 | 2 |
| PEG10 | 2.4185 | **2.92E-05** | NA | NA | 0.0001 | 4 |
| PELI1 | -1.3530 | **0.0047** | NA | NA | 0.0011 | 5 |
| PELI2 | 1.7619 | **4.44E-07** | NA | NA | 0.0012 | 7 |
| PEMT | -2.7837 | **3.83E-14** | 1.1907 | 0.4344 | 0.0001 | 2 |
| PEX10 | -1.1406 | **0.0220** | NA | NA | 0.0000 | 2 |
| PEX11B | -1.2317 | **1.55E-06** | NA | NA | 0.0000 | 1 |
| PEX12 | 1.3561 | **0.0011** | 1.0402 | 0.7463 | 0.0024 | 13 |
| PEX13 | -1.2809 | **0.0004** | 1.0519 | 0.6494 | 0.0000 | 1 |
| PEX19 | 1.1511 | **0.0049** | -1.0285 | 0.6777 | **0.0041** | 9 |
| PFDN2 | -1.1790 | **0.0002** | NA | NA | 0.0001 | 3 |
| PFDN4 | 1.0851 | 0.1761 | -1.3793 | **0.0040** | 0.0000 | 1 |
| PFDN5 | 1.2620 | **0.0002** | -1.0744 | 0.4016 | **0.0135** | **37** |
| PFKM | 1.3883 | **1.69E-06** | NA | NA | 0.0000 | 1 |
| PFN1 | -1.2403 | **0.0031** | 1.0369 | 0.6861 | 0.0000 | 2 |
| PFN2 | 1.3270 | **0.0007** | -1.0613 | 0.5871 | 0.0000 | 1 |
| PGGHG | -1.4073 | **0.0003** | NA | NA | 0.0000 | 1 |
| PGGT1B | 1.0669 | 0.3334 | 1.3067 | **0.0085** | 0.0000 | 1 |
| PGK1 | -1.5175 | **1.08E-05** | 1.0346 | 0.7505 | 0.0000 | 2 |
| PGP | -1.2699 | **0.0003** | NA | NA | 0.0001 | 2 |
| PGR | -3.6570 | **7.21E-11** | 1.0923 | 0.5864 | 0.0010 | 5 |
| PHB | -1.3028 | **4.31E-05** | NA | NA | 0.0010 | 6 |
| PHB2 | -1.1212 | **0.0361** | 1.0646 | 0.6002 | 0.0008 | 2 |
| PHF1 | 1.6366 | **2.25E-12** | -1.1366 | 0.1116 | 0.0024 | **18** |
| PHF20 | 1.0821 | 0.1987 | 1.1642 | **0.0433** | 0.0000 | 8 |
| PHF21A | 1.3356 | **2.61E-05** | 1.0500 | 0.5653 | 0.0001 | 4 |
| PHKA1 | 1.4224 | **0.0008** | -1.0970 | 0.2071 | 0.0000 | 1 |
| PHKA2 | -1.2242 | **0.0023** | -1.1410 | 0.2899 | 0.0024 | 2 |
| PHKB | -1.2657 | **0.0001** | NA | NA | 0.0016 | 3 |
| PHYH | 1.1769 | **0.0225** | 1.0055 | 0.9620 | 0.0003 | 2 |
| PIAS1 | 1.3153 | **0.0001** | NA | NA | **0.0028** | 13 |
| PIAS3 | -1.6051 | **3.52E-11** | -1.0457 | 0.6560 | 0.0006 | 7 |
| PIAS4 | -1.1201 | **0.0198** | 1.0698 | 0.7209 | 0.0024 | 8 |
| PICALM | 1.3796 | **0.0011** | 1.1286 | 0.2440 | 0.0000 | 1 |
| PID1 | 1.4670 | **0.0070** | -1.3532 | **0.0312** | 0.0010 | 3 |
| PIDD1 | -1.1454 | **0.0152** | NA | NA | 0.0001 | 4 |
| PIH1D1 | -1.0985 | **0.0354** | NA | NA | 0.0025 | 8 |
| PIK3C2B | 1.3780 | **3.56E-05** | -1.1073 | 0.3338 | 0.0000 | 1 |
| PIK3C3 | 1.1284 | **0.0429** | -1.0157 | 0.9005 | 0.0000 | 1 |
| PIK3CA | 1.3230 | **0.0208** | NA | NA | 0.0000 | 1 |
| PILRA | 1.8790 | **0.0002** | -1.2136 | 0.3091 | **0.0040** | 3 |
| PIM1 | 1.5203 | **0.0016** | -1.1810 | 0.2472 | 0.0004 | 6 |
| PIN4 | -1.1345 | **0.0347** | -1.0225 | 0.7845 | 0.0000 | 2 |
| PINK1 | 1.2762 | **3.08E-05** | NA | NA | 0.0001 | 2 |
| PISD | -1.2459 | **0.0022** | NA | NA | 0.0000 | 1 |
| PJA1 | 1.2565 | **0.0002** | 1.0182 | 0.8981 | 0.0000 | 1 |
| PJA2 | 1.5045 | **4.79E-06** | NA | NA | 0.0000 | 1 |
| PKD1 | 1.1214 | **0.0455** | NA | NA | 0.0025 | 3 |
| PKD2 | 1.8469 | **8.00E-07** | NA | NA | 0.0016 | 2 |
| PKN1 | 1.2496 | **0.0005** | 1.0346 | 0.7701 | **0.0045** | **18** |
| PKNOX1 | -1.1892 | **0.0018** | 1.0441 | 0.6128 | 0.0000 | 2 |
| PKNOX2 | NA | NA | -1.4206 | **0.0020** | 0.0023 | 11 |
| PKP2 | -1.2486 | 0.1444 | 1.4253 | **0.0249** | 0.0000 | 4 |
| PKP3 | NA | NA | 1.3394 | **0.0478** | 0.0000 | 1 |
| PKP4 | -1.4518 | **0.0001** | 1.2405 | **0.0436** | 0.0006 | 4 |
| PLAA | -1.0978 | 0.1991 | 1.2070 | **0.0332** | 0.0000 | 2 |
| PLAGL2 | -1.2397 | **0.0003** | -1.0651 | 0.5829 | 0.0008 | 6 |
| PLCD1 | -1.1766 | **0.0141** | NA | NA | 0.0016 | 4 |
| PLCG1 | -1.1211 | **0.0493** | NA | NA | **0.0031** | **14** |
| PLCG2 | 1.0568 | 0.4900 | -1.2531 | **0.0392** | 0.0001 | 2 |
| PLEC | 1.4470 | **1.01E-07** | -1.0336 | 0.6472 | 0.0000 | 2 |
| PLEKHA1 | -1.9651 | **4.23E-11** | NA | NA | 0.0008 | 2 |
| PLEKHA4 | 2.0391 | **2.43E-12** | -1.1919 | 0.1793 | 0.0000 | 1 |
| PLEKHA5 | -1.6663 | **2.16E-07** | NA | NA | 0.0000 | 1 |
| PLEKHB1 | -1.3823 | **0.0049** | 1.0078 | 0.9569 | 0.0000 | 3 |
| PLEKHB2 | -1.3175 | **0.0002** | 1.0586 | 0.4979 | 0.0025 | 10 |
| PLEKHF1 | 1.1922 | **0.0255** | -1.0284 | 0.8563 | 0.0000 | 3 |
| PLEKHF2 | -1.2569 | **0.0337** | NA | NA | **0.0263** | **51** |
| PLEKHO1 | 1.5910 | **1.86E-06** | -1.1912 | **0.0424** | 0.0007 | 3 |
| PLG | NA | NA | -1.1901 | **0.0458** | 0.0000 | 2 |
| PLK3 | 1.3206 | **0.0234** | -1.1197 | 0.4442 | 0.0009 | 3 |
| PLLP | NA | NA | 1.4944 | **0.0171** | 0.0007 | 5 |
| PLP2 | 1.3842 | **1.08E-06** | 1.0402 | 0.7708 | 0.0023 | **17** |
| PLPP3 | 1.4321 | **0.0058** | NA | NA | 0.0000 | 1 |
| PLPPR2 | 1.2249 | **0.0001** | NA | NA | **0.0033** | 7 |
| PLSCR4 | 6.5280 | **1.12E-24** | 1.1649 | 0.3277 | 0.0003 | 7 |
| PMAIP1 | -2.9579 | **3.68E-06** | 1.1234 | 0.4960 | 0.0000 | 2 |
| PMF1 | -1.2468 | **0.0003** | -1.1116 | 0.1949 | 0.0000 | 4 |
| PML | 1.1871 | **0.0019** | -1.0915 | 0.1509 | **0.0045** | 8 |
| PMP22 | 2.5983 | **1.12E-19** | -1.2913 | **0.0186** | 0.0022 | 12 |
| PMPCA | -1.2923 | **1.32E-06** | 1.1267 | 0.4919 | 0.0000 | 1 |
| PNISR | 1.2346 | **0.0281** | NA | NA | 0.0000 | 1 |
| PNKP | -1.4311 | **9.19E-07** | -1.1080 | 0.3318 | **0.0041** | **16** |
| PNMA1 | 1.2016 | **0.0428** | -1.0664 | 0.4875 | **0.0081** | **41** |
| PNN | -1.3237 | **0.0031** | NA | NA | 0.0008 | 3 |
| POGK | -1.2465 | **0.0022** | -1.1653 | 0.1967 | 0.0000 | 1 |
| POGZ | -1.0199 | 0.7465 | 1.2915 | **0.0490** | **0.0052** | **24** |
| POLB | -1.0141 | 0.8192 | -1.2590 | **0.0183** | 0.0004 | 3 |
| POLD1 | -1.2718 | **0.0055** | NA | NA | 0.0003 | 3 |
| POLD2 | -1.4755 | **1.87E-05** | NA | NA | 0.0000 | 2 |
| POLD3 | -1.1837 | **0.0042** | NA | NA | 0.0000 | 1 |
| POLD4 | -1.0139 | 0.8958 | -1.1906 | **0.0226** | 0.0000 | 1 |
| POLDIP2 | -1.2748 | **1.87E-05** | -1.1262 | 0.1673 | 0.0001 | 2 |
| POLE | -1.5460 | **8.72E-08** | NA | NA | 0.0000 | 1 |
| POLE2 | -2.0597 | **3.46E-09** | 1.1945 | 0.1768 | 0.0025 | 9 |
| POLH | -1.0897 | 0.1836 | 1.1702 | **0.0470** | 0.0001 | 4 |
| POLL | 1.1317 | **0.0335** | -1.1192 | 0.2080 | 0.0004 | 6 |
| POLR1C | -1.3841 | **2.08E-09** | 1.0051 | 0.9682 | **0.0059** | **29** |
| POLR2A | -1.2247 | **0.0038** | -1.1839 | 0.5527 | 0.0009 | 4 |
| POLR2G | -1.2853 | **6.70E-06** | -1.0127 | 0.8849 | 0.0011 | 9 |
| POMGNT1 | -1.3110 | **0.0003** | 1.2368 | 0.1679 | 0.0000 | 1 |
| POP5 | -1.2610 | **0.0015** | 1.1479 | 0.1834 | 0.0000 | 3 |
| PPARD | 1.1199 | **0.0363** | -1.1231 | 0.2531 | 0.0005 | 6 |
| PPARG | 2.0942 | **7.51E-07** | 1.0556 | 0.7613 | **0.0047** | **15** |
| PPIA | -1.3996 | **6.48E-06** | -1.0037 | 0.9689 | **0.0049** | 7 |
| PPIB | -1.5727 | **1.14E-10** | NA | NA | 0.0010 | 3 |
| PPID | -1.3104 | **0.0012** | NA | NA | 0.0000 | 1 |
| PPIG | 1.2766 | **0.0037** | NA | NA | 0.0007 | 6 |
| PPIH | -1.2940 | **2.37E-05** | NA | NA | 0.0015 | 5 |
| PPIL2 | -1.1992 | **3.57E-05** | -1.1043 | 0.2390 | 0.0000 | 1 |
| PPM1A | -1.2724 | **0.0005** | 1.1538 | 0.0764 | 0.0000 | 1 |
| PPP1CA | -1.7170 | **1.38E-11** | NA | NA | **0.0052** | 12 |
| PPP1R12A | 1.3781 | **0.0009** | NA | NA | 0.0000 | 2 |
| PPP1R16B | 2.2095 | **1.54E-10** | NA | NA | 0.0011 | 10 |
| PPP1R3C | 4.2099 | **4.32E-17** | -1.1514 | 0.4025 | 0.0001 | 2 |
| PPP1R3D | 1.1610 | **0.0133** | NA | NA | 0.0000 | 2 |
| PPP1R8 | -1.3005 | **5.34E-09** | -1.1097 | 0.1849 | 0.0002 | 3 |
| PPP2CA | -1.4044 | **1.17E-06** | 1.0174 | 0.8676 | 0.0016 | 5 |
| PPP2R1A | -1.3560 | **3.63E-07** | -1.3328 | **0.0235** | 0.0013 | 5 |
| PPP2R5D | -1.1473 | **0.0022** | 1.1044 | 0.4949 | 0.0000 | 2 |
| PPP2R5E | -1.1934 | **0.0224** | 1.1690 | 0.0681 | 0.0002 | 3 |
| PPP3CA | 1.6987 | **4.41E-11** | 1.2576 | **0.0212** | 0.0010 | 4 |
| PPP3CB | 1.3197 | **7.98E-07** | NA | NA | 0.0000 | 1 |
| PPP4C | -1.3137 | **1.06E-07** | 1.0924 | 0.4276 | 0.0004 | 2 |
| PPP5C | -1.1550 | **0.0015** | NA | NA | 0.0001 | 3 |
| PRAME | -3.2370 | **1.78E-08** | 1.2136 | 0.3434 | 0.0000 | 1 |
| PRCC | -1.1510 | **0.0117** | NA | NA | 0.0000 | 1 |
| PRDM4 | -1.3844 | **5.19E-11** | NA | NA | 0.0000 | 2 |
| PRDX2 | -1.2744 | **0.0002** | 1.0120 | 0.9462 | 0.0000 | 1 |
| PRDX3 | -1.2821 | **0.0002** | 1.0649 | 0.5558 | 0.0014 | 3 |
| PRDX4 | -1.1977 | **0.0071** | 1.0830 | 0.4150 | 0.0000 | 2 |
| PREP | -1.6757 | **7.47E-13** | 1.1563 | 0.2468 | 0.0000 | 1 |
| PRKAA1 | -1.3313 | **0.0008** | 1.0097 | 0.9400 | **0.0026** | 13 |
| PRKACA | 1.5338 | **1.12E-10** | -1.0681 | 0.5989 | 0.0003 | 5 |
| PRKACB | 1.3250 | **0.0006** | 1.2759 | **0.0161** | 0.0001 | 3 |
| PRKAG1 | -1.1862 | **0.0024** | 1.1942 | 0.1332 | 0.0003 | 5 |
| PRKAR1A | 1.1829 | **0.0267** | 1.3695 | **0.0139** | 0.0013 | 9 |
| PRKAR1B | 1.6504 | **3.43E-08** | -1.1052 | 0.2098 | **0.0063** | **22** |
| PRKCD | -1.2621 | **0.0046** | 1.1003 | 0.3764 | 0.0000 | 2 |
| PRKCI | 1.2015 | **0.0238** | 1.6655 | **0.0040** | 0.0001 | 3 |
| PRKCQ | -1.7751 | **7.75E-06** | 1.3273 | 0.1211 | 0.0003 | 4 |
| PRKCZ | -1.6938 | **8.04E-12** | NA | NA | **0.0027** | 10 |
| PRKDC | -1.4717 | **4.09E-06** | 1.0792 | 0.5416 | 0.0005 | 3 |
| PRKRA | -1.1447 | **0.0044** | 1.0005 | 0.9960 | 0.0006 | 6 |
| PRKX | -3.0974 | **1.72E-08** | 1.0711 | 0.8065 | 0.0000 | 1 |
| PRLR | -1.9725 | **1.71E-05** | -1.0812 | 0.6081 | 0.0000 | 1 |
| PRM1 | NA | NA | -1.5255 | **0.0486** | 0.0000 | 1 |
| PRMT1 | -1.2623 | **4.88E-05** | NA | NA | **0.0062** | 12 |
| PRMT2 | 1.3027 | **1.44E-05** | 1.1787 | 0.0871 | 0.0000 | 1 |
| PRMT3 | -1.3371 | **0.0009** | 1.2471 | 0.0841 | 0.0000 | 1 |
| PRMT5 | -1.3822 | **5.86E-07** | NA | NA | **0.0050** | 11 |
| PRNP | 2.1323 | **7.29E-14** | 1.1020 | 0.3885 | **0.0076** | 12 |
| PRPF19 | -1.1908 | **0.0017** | 1.2844 | **0.0355** | 0.0004 | 7 |
| PRPF3 | -1.0968 | 0.1885 | -1.2807 | **0.0107** | **0.0028** | 13 |
| PRPF31 | -1.1549 | **0.0022** | NA | NA | **0.0076** | **31** |
| PRPF4 | -1.3328 | **3.04E-11** | NA | NA | 0.0004 | 6 |
| PRPF40A | -1.2017 | **0.0025** | NA | NA | 0.0007 | 5 |
| PRPS2 | -1.5294 | **6.08E-06** | 1.1318 | 0.3853 | 0.0003 | 3 |
| PRPSAP1 | -1.2478 | **2.48E-06** | -1.1770 | 0.0533 | 0.0010 | 5 |
| PRPSAP2 | -1.2343 | **0.0022** | 1.1268 | 0.1804 | 0.0000 | 2 |
| PRR4 | 1.0876 | 0.3059 | -1.3972 | **0.0432** | 0.0001 | 3 |
| PRR5 | 1.1618 | **0.0371** | NA | NA | 0.0000 | 2 |
| PRRC1 | -1.2193 | **0.0004** | 1.2707 | **0.0402** | 0.0000 | 1 |
| PSAP | NA | NA | 1.2948 | **0.0385** | 0.0000 | 1 |
| PSD4 | -1.5203 | **2.68E-05** | 1.0787 | 0.5558 | 0.0000 | 1 |
| PSEN1 | -1.4103 | **3.47E-06** | 1.0594 | 0.5842 | **0.0053** | 12 |
| PSEN2 | 1.2465 | **0.0002** | NA | NA | 0.0011 | 6 |
| PSENEN | -1.3108 | **0.0004** | 1.0454 | 0.7737 | 0.0000 | 1 |
| PSIP1 | 1.4490 | **0.0005** | 1.1202 | 0.4015 | 0.0001 | 2 |
| PSMA3 | -1.4608 | **2.14E-07** | NA | NA | **0.0074** | **19** |
| PSMA4 | -1.4291 | **5.82E-06** | 1.1235 | 0.1999 | 0.0004 | 7 |
| PSMA5 | -1.3832 | **1.25E-07** | 1.1660 | 0.0841 | 0.0003 | 2 |
| PSMA6 | -1.4927 | **0.0002** | -1.0404 | 0.6205 | **0.0035** | 11 |
| PSMA7 | -1.1586 | **0.0029** | NA | NA | 0.0006 | 7 |
| PSMB1 | -1.2167 | **5.82E-06** | 1.0123 | 0.8667 | **0.0107** | **29** |
| PSMB10 | -1.2017 | **0.0258** | -1.1687 | **0.0477** | 0.0000 | 1 |
| PSMB3 | -1.3470 | **4.69E-06** | NA | NA | 0.0009 | 7 |
| PSMB4 | -1.1267 | **0.0127** | -1.0427 | 0.6158 | 0.0012 | 13 |
| PSMB5 | -1.3183 | **1.89E-06** | NA | NA | 0.0012 | 10 |
| PSMB6 | -1.2609 | **0.0001** | NA | NA | 0.0003 | 5 |
| PSMB7 | -1.2257 | **1.64E-05** | -1.1027 | 0.0812 | 0.0000 | 6 |
| PSMB8 | -1.3061 | **0.0001** | NA | NA | 0.0014 | 8 |
| PSMC3 | -1.4116 | **2.95E-06** | NA | NA | 0.0017 | 9 |
| PSMC4 | -1.4694 | **1.53E-07** | NA | NA | **0.0034** | 5 |
| PSMC5 | -1.2134 | **1.20E-06** | NA | NA | **0.0095** | **16** |
| PSMC6 | -1.1810 | **0.0408** | NA | NA | **0.0040** | 11 |
| PSMD10 | -1.2277 | **0.0007** | -1.0042 | 0.9692 | 0.0000 | 1 |
| PSMD14 | -1.5409 | **3.16E-07** | NA | NA | 0.0008 | 2 |
| PSMD2 | -1.1578 | **0.0029** | NA | NA | **0.0040** | 6 |
| PSMD3 | -1.3374 | **4.39E-05** | NA | NA | 0.0015 | 5 |
| PSMD6 | -1.2638 | **0.0001** | 1.1346 | 0.2009 | 0.0002 | 2 |
| PSMD7 | -1.3440 | **1.64E-05** | -1.0063 | 0.9533 | 0.0000 | 1 |
| PSMD9 | -1.1849 | **0.0022** | 1.0010 | 0.9916 | 0.0002 | 5 |
| PSME3 | -1.7324 | **4.04E-13** | -1.1242 | 0.3023 | **0.0045** | 12 |
| PSME4 | -1.6056 | **3.77E-11** | 1.2338 | **0.0281** | 0.0000 | 1 |
| PSMG1 | -1.6847 | **2.03E-06** | -1.0977 | 0.4718 | 0.0000 | 1 |
| PSORS1C2 | -1.4197 | **0.0082** | 1.0853 | 0.7469 | 0.0023 | 8 |
| PSPC1 | -1.1771 | **0.0291** | 1.1576 | 0.1226 | 0.0000 | 2 |
| PSTPIP1 | 1.6160 | **0.0007** | 1.0188 | 0.9135 | **0.0054** | **21** |
| PTBP1 | -1.3753 | **5.87E-12** | -1.0131 | 0.9131 | 0.0011 | 7 |
| PTGES | 1.3716 | **0.0030** | -1.1147 | 0.0645 | 0.0001 | 4 |
| PTK2B | 1.4976 | **1.10E-06** | -1.1464 | 0.1043 | **0.0032** | 5 |
| PTPN1 | 1.1590 | **0.0211** | -1.0904 | 0.3953 | 0.0001 | 3 |
| PTPN14 | -1.1738 | **0.0297** | 1.2294 | 0.0717 | 0.0000 | 1 |
| PTPN2 | 1.3474 | **0.0002** | NA | NA | 0.0000 | 1 |
| PTPN4 | -1.2037 | **0.0071** | NA | NA | 0.0000 | 1 |
| PTPN7 | 1.0408 | 0.7860 | -1.2299 | **0.0388** | 0.0002 | 2 |
| PTPN9 | -1.1742 | **0.0019** | -1.0127 | 0.8980 | **0.0041** | 13 |
| PTPRB | 1.8987 | **6.48E-11** | 1.0500 | 0.6963 | 0.0000 | 1 |
| PTPRJ | -1.0559 | 0.5914 | 1.3792 | **0.0112** | 0.0000 | 2 |
| PTPRK | NA | NA | 1.2619 | **0.0462** | 0.0000 | 1 |
| PUDP | -1.1438 | **0.0190** | NA | NA | 0.0000 | 1 |
| PUS7L | -1.0948 | 0.3043 | 1.2181 | **0.0453** | 0.0000 | 1 |
| PYGM | NA | NA | -1.4579 | **0.0043** | 0.0000 | 1 |
| PYGO1 | 1.2640 | **0.0012** | -1.0071 | 0.9511 | 0.0000 | 1 |
| QARS | -1.2367 | **1.39E-06** | 1.0057 | 0.9611 | **0.0032** | **21** |
| QPCTL | -1.2487 | **0.0029** | 1.0583 | 0.6198 | 0.0000 | 1 |
| QPRT | -1.5412 | **4.57E-05** | NA | NA | 0.0000 | 1 |
| QRICH1 | -1.1082 | **0.0043** | NA | NA | **0.0041** | **15** |
| RAB11B | -1.1762 | **0.0012** | 1.0358 | 0.6756 | 0.0016 | 4 |
| RAB11FIP1 | -1.6482 | **0.0005** | 1.3526 | **0.0465** | 0.0000 | 2 |
| RAB11FIP2 | 1.6766 | **1.88E-09** | NA | NA | 0.0020 | 5 |
| RAB11FIP5 | 1.3657 | **1.64E-06** | NA | NA | 0.0008 | 3 |
| RAB14 | -1.2489 | **0.0008** | 1.1541 | 0.1997 | 0.0008 | 3 |
| RAB1A | -1.2107 | **0.0041** | -1.0267 | 0.8115 | 0.0022 | 5 |
| RAB21 | -1.1055 | 0.3473 | 1.2714 | **0.0134** | 0.0000 | 2 |
| RAB33B | 1.6877 | **9.28E-08** | 1.3722 | **0.0182** | 0.0000 | 1 |
| RAB3A | 1.6690 | **2.55E-06** | -1.1191 | 0.1534 | 0.0000 | 1 |
| RAB5A | -1.2376 | **0.0012** | 1.0315 | 0.7863 | 0.0005 | 3 |
| RAB5C | 1.1467 | **0.0426** | -1.0397 | 0.6767 | 0.0000 | 1 |
| RABGAP1L | 1.5836 | **0.0001** | NA | NA | 0.0025 | 4 |
| RABGEF1 | 1.2735 | **0.0054** | -1.0396 | 0.6560 | 0.0004 | 4 |
| RABIF | -1.2234 | **0.0006** | 1.0662 | 0.4307 | 0.0013 | 6 |
| RAC2 | 1.3699 | **0.0278** | -1.1413 | 0.3917 | 0.0011 | 3 |
| RAD23A | 1.1362 | **0.0063** | 1.2774 | **0.0428** | 0.0017 | 9 |
| RAD51 | -1.8830 | **5.45E-07** | 1.1852 | 0.3561 | 0.0004 | 4 |
| RAD51B | 1.1655 | **0.0223** | 1.0742 | 0.5113 | 0.0000 | 1 |
| RAD52 | -1.1641 | **0.0202** | -1.1345 | 0.1517 | 0.0006 | 6 |
| RAD54L | -1.8853 | **2.30E-08** | NA | NA | 0.0002 | 2 |
| RAD9A | -1.2636 | **1.19E-05** | -1.1855 | 0.1452 | 0.0007 | 4 |
| RAE1 | -1.1247 | **0.0009** | NA | NA | 0.0000 | 1 |
| RAF1 | -1.0833 | **0.0230** | -1.0397 | 0.6765 | **0.0056** | **15** |
| RALA | -1.1374 | 0.1596 | 1.2409 | **0.0148** | 0.0010 | 3 |
| RALB | 1.1157 | **0.0239** | NA | NA | 0.0003 | 2 |
| RALGDS | 1.6694 | **3.74E-12** | 1.0445 | 0.7810 | 0.0001 | 5 |
| RAN | -1.2674 | **0.0045** | 1.1056 | 0.3728 | 0.0021 | 8 |
| RANBP1 | -1.4647 | **2.04E-07** | NA | NA | 0.0000 | 2 |
| RANBP10 | -1.1527 | **0.0036** | -1.1059 | 0.1005 | 0.0000 | 1 |
| RANBP2 | -1.3756 | **2.55E-05** | NA | NA | 0.0004 | 5 |
| RANGAP1 | -1.3451 | **3.82E-07** | NA | NA | 0.0000 | 2 |
| RANGRF | -1.1913 | **0.0047** | -1.0592 | 0.4015 | 0.0000 | 1 |
| RAP1A | 1.5947 | **1.54E-07** | NA | NA | 0.0002 | 3 |
| RAP2A | 1.4452 | **1.27E-07** | 1.2362 | 0.0576 | 0.0001 | 2 |
| RAPGEF1 | 1.3238 | **4.30E-05** | -1.1513 | 0.1347 | 0.0000 | 2 |
| RAPGEF4 | 1.6502 | **0.0003** | NA | NA | 0.0000 | 1 |
| RARA | 1.1868 | **0.0128** | -1.0010 | 0.9960 | **0.0049** | **15** |
| RARG | 1.1746 | **0.0074** | -1.1272 | 0.2341 | 0.0002 | 3 |
| RARS | -1.4828 | **0.0001** | NA | NA | 0.0002 | 2 |
| RASIP1 | 1.7415 | **3.22E-09** | NA | NA | 0.0000 | 1 |
| RASSF4 | 1.6658 | **0.0006** | 1.2195 | 0.3119 | 0.0000 | 1 |
| RB1CC1 | 1.2293 | **0.0312** | NA | NA | 0.0008 | 2 |
| RBBP7 | -1.1470 | **0.0436** | 1.0214 | 0.8154 | 0.0000 | 4 |
| RBBP8 | -2.2285 | **2.48E-11** | NA | NA | 0.0016 | 9 |
| RBFOX2 | 1.1717 | **0.0366** | NA | NA | 0.0011 | 12 |
| RBM10 | -1.2443 | **0.0002** | 1.0620 | 0.5732 | 0.0014 | 12 |
| RBM12 | -1.3013 | **7.82E-06** | -1.1387 | 0.1514 | 0.0008 | 2 |
| RBM14 | -1.2586 | **0.0012** | NA | NA | 0.0005 | 4 |
| RBM15 | -1.1384 | **0.0292** | -1.0263 | 0.8671 | 0.0001 | 2 |
| RBM15B | -1.2294 | **0.0001** | NA | NA | 0.0000 | 1 |
| RBM23 | -1.2031 | **6.84E-06** | -1.0428 | 0.6852 | 0.0004 | 5 |
| RBM25 | -1.1444 | **0.0249** | NA | NA | 0.0000 | 1 |
| RBM38 | 1.2973 | **0.0002** | -1.3641 | 0.1303 | 0.0000 | 1 |
| RBM4 | -1.2138 | **0.0027** | 1.1790 | **0.0449** | 0.0000 | 1 |
| RBM47 | -2.7437 | **5.22E-10** | NA | NA | 0.0000 | 1 |
| RBM5 | -1.1628 | **0.0181** | -1.0442 | 0.6407 | 0.0000 | 2 |
| RBM8A | -1.3378 | **2.85E-06** | 1.0206 | 0.8032 | 0.0000 | 5 |
| RBPJ | 1.3035 | **0.0002** | NA | NA | 0.0016 | 4 |
| RBPMS | 1.3520 | **0.0012** | -1.0759 | 0.6136 | **0.0142** | **48** |
| RBX1 | -1.3299 | **9.29E-08** | 1.0242 | 0.7593 | 0.0001 | 4 |
| RC3H2 | -1.6180 | **8.19E-11** | -1.0227 | 0.8382 | 0.0000 | 1 |
| RCAN3 | NA | NA | 1.2841 | **0.0097** | 0.0000 | 1 |
| RCC1 | -1.4892 | **5.62E-08** | 1.1504 | 0.2328 | 0.0002 | 3 |
| RCN1 | -1.6484 | **8.01E-06** | 1.1358 | 0.5054 | 0.0024 | 10 |
| RCN2 | -1.3118 | **0.0057** | 1.4424 | **0.0077** | 0.0000 | 1 |
| RCN3 | 1.7073 | **8.26E-10** | NA | NA | 0.0000 | 1 |
| RCOR1 | -1.7630 | **1.18E-11** | 1.0399 | 0.6889 | 0.0002 | 3 |
| REEP1 | 2.5744 | **1.25E-09** | NA | NA | 0.0004 | 5 |
| REEP4 | -1.2333 | **0.0005** | 1.0091 | 0.9429 | **0.0067** | **27** |
| REL | 1.2022 | **0.0400** | NA | NA | **0.0440** | **91** |
| REM1 | 1.2212 | 0.0821 | -1.2916 | **0.0057** | 0.0000 | 1 |
| REPIN1 | -1.1738 | **0.0034** | 1.0830 | 0.3137 | 0.0000 | 1 |
| RER1 | -1.2076 | **0.0011** | 1.1342 | 0.1186 | 0.0000 | 2 |
| RETREG1 | -2.8719 | **3.95E-08** | NA | NA | 0.0000 | 1 |
| RFC1 | 1.1481 | **0.0211** | 1.2263 | 0.0748 | 0.0000 | 1 |
| RFC2 | -1.3141 | **0.0001** | 1.1648 | 0.3561 | 0.0000 | 1 |
| RFC5 | -1.4629 | **1.91E-08** | 1.3274 | **0.0174** | 0.0000 | 5 |
| RFWD3 | -1.4300 | **2.32E-05** | 1.1348 | 0.4418 | 0.0000 | 3 |
| RFX1 | -1.1559 | **0.0073** | NA | NA | 0.0000 | 1 |
| RFX5 | NA | NA | -1.2785 | **0.0032** | 0.0000 | 1 |
| RFXANK | -1.3494 | **3.16E-07** | 1.0608 | 0.6965 | 0.0000 | 1 |
| RGS14 | 1.0378 | 0.6497 | -1.2137 | **0.0235** | 0.0000 | 1 |
| RGS17 | NA | NA | 1.2319 | **0.0171** | 0.0014 | 4 |
| RGS2 | 6.5739 | **6.58E-19** | -1.3847 | 0.1243 | 0.0016 | 4 |
| RGS3 | 1.2016 | **0.0142** | NA | NA | 0.0003 | 2 |
| RHOB | 1.5087 | **0.0003** | NA | NA | 0.0000 | 1 |
| RHOG | 1.3463 | **1.91E-05** | -1.1766 | 0.0942 | 0.0000 | 1 |
| RHOT2 | -1.1029 | **0.0367** | 1.1169 | 0.2613 | 0.0000 | 1 |
| RIF1 | -1.1816 | **0.0393** | NA | NA | 0.0000 | 2 |
| RIN1 | -1.0627 | 0.4646 | -1.7280 | **0.0020** | **0.0065** | **17** |
| RIN3 | NA | NA | -1.2476 | **0.0065** | 0.0002 | 3 |
| RING1 | -1.1046 | **0.0126** | NA | NA | 0.0007 | 6 |
| RIPK1 | -1.1334 | **0.0147** | NA | NA | 0.0006 | 5 |
| RIPK2 | -1.5464 | **5.35E-07** | 1.1229 | 0.1802 | 0.0000 | 1 |
| RMDN3 | -1.1404 | **0.0036** | NA | NA | 0.0021 | 12 |
| RMI1 | -1.8868 | **7.56E-07** | 1.5338 | **0.0093** | 0.0000 | 1 |
| RNASE1 | 4.6270 | **2.00E-23** | -1.1964 | 0.1836 | 0.0000 | 1 |
| RNASEH1 | -1.1969 | **0.0006** | 1.0039 | 0.9785 | 0.0001 | 3 |
| RNF11 | 1.2738 | **0.0039** | 1.0091 | 0.9317 | 0.0016 | 11 |
| RNF111 | 1.1342 | **0.0438** | NA | NA | 0.0009 | 9 |
| RNF114 | -1.1615 | **0.0003** | NA | NA | 0.0000 | 3 |
| RNF122 | 1.3127 | **0.0015** | 1.1812 | 0.1607 | 0.0001 | 3 |
| RNF125 | 1.3615 | **0.0072** | 1.0517 | 0.7456 | 0.0001 | 3 |
| RNF13 | 1.6378 | **2.04E-09** | NA | NA | 0.0000 | 2 |
| RNF138 | -1.1916 | **0.0133** | NA | NA | 0.0002 | 4 |
| RNF14 | -1.3306 | **0.0001** | 1.1597 | 0.1078 | 0.0002 | 5 |
| RNF141 | -1.2015 | **0.0158** | -1.1526 | 0.0742 | 0.0000 | 1 |
| RNF170 | -1.2572 | **0.0102** | 1.2554 | 0.0521 | 0.0010 | 4 |
| RNF19B | 1.5100 | **2.91E-06** | 1.1462 | 0.1409 | 0.0013 | 11 |
| RNF2 | 1.2265 | **0.0085** | 1.0808 | 0.5566 | **0.0056** | **15** |
| RNF208 | -1.3378 | **0.0016** | 1.1289 | 0.1587 | 0.0001 | 3 |
| RNF216 | 1.2707 | **1.98E-06** | NA | NA | 0.0012 | 7 |
| RNF32 | -1.1821 | **0.0019** | 1.0058 | 0.9471 | 0.0000 | 1 |
| RNF34 | -1.1916 | **3.58E-05** | -1.0842 | 0.2035 | 0.0000 | 1 |
| RNF38 | 1.5451 | **2.56E-07** | 1.3155 | **0.0032** | 0.0000 | 2 |
| RNF4 | -1.1820 | **1.76E-05** | -1.0883 | 0.2466 | **0.0202** | **38** |
| RNF40 | -1.1055 | **0.0449** | NA | NA | 0.0003 | 4 |
| RNF8 | -1.4673 | **1.63E-07** | 1.2425 | 0.1041 | **0.0065** | **20** |
| RNH1 | 1.1836 | **0.0172** | NA | NA | 0.0000 | 1 |
| ROBO1 | 1.3182 | **0.0355** | 1.2905 | 0.0644 | 0.0000 | 1 |
| ROR2 | 1.2133 | **0.0208** | NA | NA | 0.0013 | 10 |
| RPA2 | 1.1666 | **0.0100** | 1.0574 | 0.4637 | **0.0038** | 10 |
| RPA3 | -1.3425 | **0.0001** | -1.0289 | 0.7880 | 0.0000 | 1 |
| RPGRIP1L | -1.4125 | **0.0022** | NA | NA | 0.0000 | 1 |
| RPIA | -1.5664 | **5.63E-12** | 1.0289 | 0.7644 | 0.0005 | 6 |
| RPL23 | 1.1753 | **0.0303** | -1.1408 | 0.0550 | 0.0004 | 2 |
| RPL30 | 1.1712 | **0.0005** | -1.1292 | 0.0596 | 0.0000 | 1 |
| RPL34 | 1.1795 | **0.0013** | -1.1457 | 0.0568 | 0.0000 | 2 |
| RPL36 | 1.2066 | **0.0012** | -1.0895 | 0.5829 | 0.0000 | 1 |
| RPL4 | -1.1238 | **0.0230** | 1.0316 | 0.8321 | 0.0000 | 1 |
| RPN1 | -1.4979 | **4.59E-12** | NA | NA | 0.0005 | 2 |
| RPRD1A | -1.8508 | **1.26E-10** | 1.0699 | 0.5021 | 0.0003 | 5 |
| RPS14 | 1.1254 | 0.2482 | 1.3314 | **0.0012** | 0.0004 | 2 |
| RPS20 | 1.2647 | **0.0001** | -1.0486 | 0.3101 | 0.0000 | 1 |
| RPS23 | 1.1429 | **0.0035** | -1.0906 | 0.2020 | 0.0000 | 1 |
| RPS25 | 1.2199 | **0.0008** | -1.1727 | **0.0289** | 0.0007 | 3 |
| RPS6KA1 | -1.3531 | **0.0045** | NA | NA | 0.0000 | 1 |
| RPS6KA2 | 1.4422 | **2.48E-08** | 1.0833 | 0.4904 | 0.0000 | 2 |
| RPS6KA3 | 1.3544 | **0.0004** | -1.0419 | 0.7286 | 0.0002 | 4 |
| RPS9 | 1.1950 | **0.0022** | 1.1564 | 0.1598 | 0.0000 | 1 |
| RRAGC | 1.5158 | **7.42E-10** | 1.1151 | 0.1837 | 0.0000 | 1 |
| RRAS | 1.8777 | **1.46E-08** | -1.1521 | 0.2485 | 0.0002 | 3 |
| RRAS2 | -1.6272 | **1.37E-06** | 1.1174 | 0.2155 | 0.0002 | 3 |
| RSRP1 | -1.3350 | **0.0078** | NA | NA | 0.0000 | 1 |
| RSU1 | 1.2434 | **0.0012** | NA | NA | 0.0000 | 1 |
| RTN4 | 1.4080 | **2.06E-06** | 1.1003 | 0.4889 | 0.0009 | 5 |
| RUBCN | 1.2606 | **1.92E-05** | NA | NA | 0.0000 | 2 |
| RUFY3 | 1.3272 | **0.0032** | 1.1508 | 0.2069 | 0.0000 | 1 |
| RUNDC3A | 1.1913 | **0.0352** | -1.2223 | 0.1842 | 0.0022 | 11 |
| RUNX3 | 1.3271 | **0.0498** | -1.0086 | 0.9682 | 0.0000 | 1 |
| RUSC1 | -1.2521 | **0.0001** | NA | NA | **0.0042** | **24** |
| RUVBL1 | -1.6835 | **2.55E-10** | NA | NA | 0.0008 | 2 |
| RUVBL2 | -1.6093 | **5.50E-11** | NA | NA | **0.0029** | 4 |
| RXRA | 1.3643 | **2.34E-06** | NA | NA | 0.0007 | 11 |
| S100A4 | 2.0916 | **1.17E-05** | -1.0630 | 0.7590 | 0.0008 | 2 |
| S100A8 | 5.2611 | **4.74E-09** | -1.1066 | 0.8424 | 0.0008 | 2 |
| S100A9 | 4.4624 | **3.07E-11** | 1.0301 | 0.9435 | 0.0000 | 1 |
| SACM1L | -1.1641 | **0.0438** | NA | NA | 0.0009 | 5 |
| SAE1 | -1.6648 | **2.15E-10** | NA | NA | 0.0000 | 1 |
| SAFB | -1.0893 | **0.0447** | NA | NA | 0.0016 | 4 |
| SAFB2 | NA | NA | 1.2758 | **0.0280** | 0.0000 | 1 |
| SALL1 | -7.9243 | **1.22E-21** | 1.0325 | 0.8865 | 0.0001 | 2 |
| SALL2 | 1.3146 | **0.0104** | NA | NA | 0.0008 | 5 |
| SAMHD1 | NA | NA | 1.3086 | **0.0258** | 0.0000 | 2 |
| SAP18 | 1.3066 | **4.19E-05** | NA | NA | 0.0001 | 2 |
| SAR1A | -1.3917 | **0.0001** | 1.1326 | 0.1478 | **0.0041** | **26** |
| SAR1B | -1.3241 | **0.0018** | 1.0158 | 0.9135 | 0.0000 | 1 |
| SARS2 | -1.3339 | **0.0011** | NA | NA | 0.0000 | 1 |
| SART3 | -1.0574 | 0.2666 | 1.3039 | **0.0133** | 0.0001 | 5 |
| SAT1 | -1.8680 | **3.79E-07** | NA | NA | 0.0021 | 13 |
| SATB1 | 1.2523 | **0.0034** | NA | NA | 0.0000 | 5 |
| SCAF8 | -1.1702 | **0.0178** | 1.1050 | 0.1868 | 0.0000 | 1 |
| SCAI | 1.1880 | **0.0211** | NA | NA | 0.0000 | 2 |
| SCAMP1 | -1.1140 | 0.1519 | 1.5288 | **0.0002** | 0.0014 | 7 |
| SCAMP3 | -1.2116 | **0.0002** | NA | NA | 0.0000 | 1 |
| SCAMP4 | -1.1866 | **0.0040** | 1.0343 | 0.7418 | 0.0008 | 5 |
| SCAND1 | -1.2168 | **0.0010** | NA | NA | **0.0030** | 10 |
| SCARB2 | 1.1767 | **0.0048** | 1.2266 | 0.1208 | 0.0002 | 2 |
| SCARF1 | 1.4121 | **2.24E-05** | 1.0888 | 0.5203 | 0.0000 | 2 |
| SCCPDH | -1.7307 | **9.78E-07** | -1.0093 | 0.9378 | 0.0000 | 1 |
| SCN5A | -1.6321 | **5.00E-05** | -1.2792 | 0.1409 | 0.0017 | 4 |
| SCNM1 | 1.1098 | **0.0441** | NA | NA | **0.0146** | **46** |
| SCO2 | -1.2218 | **0.0309** | 1.2222 | 0.0919 | 0.0000 | 2 |
| SCRIB | -1.4138 | **2.04E-06** | 1.1823 | **0.0341** | 0.0005 | 4 |
| SCYL3 | -1.1893 | **0.0053** | NA | NA | 0.0001 | 2 |
| SEC13 | -1.4041 | **7.44E-08** | NA | NA | **0.0057** | 4 |
| SEC23A | 1.2123 | **0.0383** | -1.1144 | 0.2897 | **0.0030** | 11 |
| SEC23B | -1.7887 | **4.49E-10** | 1.2145 | 0.1433 | 0.0010 | 7 |
| SEC23IP | -1.2589 | **0.0006** | 1.0748 | 0.4325 | 0.0001 | 3 |
| SEC24C | -1.2782 | **0.0001** | NA | NA | 0.0001 | 4 |
| SEC24D | -1.3118 | **0.0065** | NA | NA | 0.0001 | 4 |
| SEC31A | 1.1199 | **0.0241** | -1.1487 | 0.2207 | 0.0008 | 4 |
| SEC61A1 | -1.6426 | **1.61E-10** | 1.2607 | 0.0887 | 0.0000 | 1 |
| SEC61B | -1.4186 | **2.73E-07** | 1.0250 | 0.7884 | 0.0000 | 1 |
| SEC61G | -1.5188 | **1.32E-07** | -1.2046 | **0.0287** | 0.0000 | 2 |
| SEH1L | -1.2728 | **4.59E-05** | 1.1634 | 0.1068 | **0.0040** | 2 |
| SELENBP1 | 1.2850 | **0.0424** | -1.0503 | 0.6951 | 0.0000 | 2 |
| SEM1 | -1.4474 | **5.06E-07** | NA | NA | 0.0000 | 2 |
| SEMA3G | 2.2026 | **4.67E-07** | 1.1349 | 0.4553 | 0.0000 | 1 |
| SEMA4D | -1.4014 | **0.0014** | -1.0108 | 0.9249 | 0.0000 | 1 |
| SEPT2 | -1.2703 | **1.34E-05** | -1.0590 | 0.5358 | 0.0001 | 2 |
| SEPT7 | 1.6717 | **2.37E-10** | NA | NA | 0.0000 | 1 |
| SERP1 | -1.1515 | **0.0455** | NA | NA | 0.0015 | 10 |
| SERPINE1 | 2.3547 | **0.0004** | -1.2185 | 0.2034 | 0.0003 | 4 |
| SERTAD2 | 1.5130 | **3.79E-06** | -1.0512 | 0.6718 | 0.0024 | 4 |
| SERTAD3 | -1.2254 | **0.0053** | NA | NA | 0.0025 | **14** |
| SET | -1.3483 | **8.92E-06** | 1.0594 | 0.6714 | 0.0024 | 10 |
| SETBP1 | 1.3175 | **0.0058** | 1.0530 | 0.6648 | 0.0005 | 3 |
| SETD2 | -1.1251 | **0.0247** | 1.3967 | **0.0461** | 0.0000 | 2 |
| SETD5 | -1.1369 | **0.0172** | 1.0127 | 0.9250 | 0.0000 | 4 |
| SETDB1 | -1.1578 | **0.0021** | 1.0186 | 0.8162 | 0.0006 | 5 |
| SEZ6L2 | -1.3704 | **0.0052** | NA | NA | 0.0000 | 1 |
| SF3A2 | 1.1981 | **0.0106** | -1.1046 | 0.6978 | 0.0006 | 6 |
| SF3B2 | -1.1165 | **0.0121** | 1.1874 | 0.2055 | 0.0008 | 5 |
| SF3B4 | -1.2055 | **0.0012** | -1.1195 | 0.1122 | 0.0011 | 12 |
| SF3B5 | -1.2161 | **0.0005** | NA | NA | 0.0000 | 1 |
| SFMBT1 | -1.2376 | **0.0004** | -1.0378 | 0.6569 | 0.0000 | 2 |
| SFPQ | -1.3270 | **1.15E-05** | 1.3207 | **0.0322** | 0.0003 | 3 |
| SFXN1 | -1.0696 | 0.3287 | 1.2726 | **0.0239** | 0.0013 | 6 |
| SFXN3 | 1.6107 | **2.90E-10** | -1.2879 | 0.0626 | 0.0000 | 6 |
| SGF29 | 1.2341 | **0.0006** | NA | NA | **0.0026** | 8 |
| SGPL1 | -1.4832 | **5.70E-09** | 1.0677 | 0.5441 | 0.0005 | 12 |
| SGSM3 | -1.2091 | **0.0003** | 1.1286 | 0.2316 | 0.0000 | 2 |
| SH2B1 | 1.1460 | **0.0018** | NA | NA | 0.0000 | 2 |
| SH3BP1 | 1.0798 | 0.4049 | -1.2617 | **0.0379** | 0.0000 | 2 |
| SH3BP4 | 1.2628 | **0.0023** | NA | NA | 0.0016 | 2 |
| SH3BP5 | 2.6258 | **1.26E-18** | -1.1706 | 0.1817 | 0.0000 | 1 |
| SH3YL1 | -2.4977 | **6.16E-14** | NA | NA | 0.0000 | 1 |
| SHC2 | 1.2298 | **0.0460** | -1.1636 | 0.1257 | 0.0001 | 2 |
| SHMT1 | -1.2995 | **0.0032** | 1.1224 | 0.4340 | 0.0000 | 1 |
| SHMT2 | -1.2692 | **0.0003** | NA | NA | 0.0000 | 2 |
| SHOC2 | 1.0828 | 0.2544 | 1.2887 | **0.0201** | 0.0001 | 2 |
| SHQ1 | -1.3633 | **2.56E-07** | 1.1890 | **0.0484** | 0.0000 | 1 |
| SHTN1 | -1.5345 | **4.42E-05** | NA | NA | 0.0008 | 3 |
| SIAH1 | -1.0392 | 0.6619 | 1.2574 | **0.0404** | **0.0315** | **53** |
| SIAH2 | -1.3358 | **1.16E-05** | 1.1161 | 0.3154 | 0.0011 | 6 |
| SINHCAF | -1.8966 | **1.16E-09** | NA | NA | 0.0000 | 3 |
| SIPA1L1 | 1.2148 | **0.0111** | 1.1262 | 0.1482 | 0.0000 | 1 |
| SIRPA | 2.9719 | **6.15E-17** | -1.2174 | 0.0533 | 0.0003 | 4 |
| SIRT6 | -1.2473 | **0.0001** | -1.0977 | 0.1436 | 0.0000 | 1 |
| SKA1 | -1.9818 | **0.0001** | 1.1065 | 0.5653 | 0.0000 | 1 |
| SKAP1 | -1.7687 | **1.68E-06** | 1.0976 | 0.5328 | 0.0008 | 2 |
| SKI | 1.4477 | **2.03E-06** | NA | NA | 0.0011 | 3 |
| SLA | 1.6618 | **0.0008** | -1.0997 | 0.6256 | 0.0000 | 3 |
| SLC12A7 | -1.2036 | **0.0117** | 1.0429 | 0.7501 | 0.0000 | 4 |
| SLC15A2 | -3.1687 | **2.85E-06** | 1.8882 | **0.0092** | 0.0005 | 6 |
| SLC16A2 | 1.6459 | **2.58E-07** | NA | NA | 0.0001 | 9 |
| SLC16A3 | -1.4620 | **0.0299** | 1.2820 | 0.0943 | 0.0000 | 1 |
| SLC16A7 | 1.3574 | **0.0109** | 1.2189 | 0.0916 | 0.0000 | 4 |
| SLC17A9 | 1.0690 | 0.5344 | -1.3216 | **0.0330** | 0.0000 | 2 |
| SLC26A6 | -1.1791 | **0.0411** | -1.2994 | **0.0051** | 0.0000 | 4 |
| SLC2A1 | -2.2820 | **1.10E-07** | 1.0142 | 0.9235 | 0.0016 | 2 |
| SLC2A3 | 2.1531 | **2.23E-05** | -1.1198 | 0.4247 | 0.0000 | 1 |
| SLC34A2 | NA | NA | 1.6183 | **0.0350** | 0.0000 | 3 |
| SLC35A1 | 1.2776 | **0.0038** | NA | NA | 0.0023 | 10 |
| SLC35B1 | -1.7807 | **8.52E-15** | NA | NA | 0.0000 | 2 |
| SLC35F6 | -1.1883 | **0.0327** | NA | NA | 0.0000 | 1 |
| SLC35G2 | 2.0375 | **9.02E-13** | NA | NA | 0.0000 | 1 |
| SLC37A4 | -1.5606 | **5.52E-09** | 1.0767 | 0.5610 | 0.0000 | 1 |
| SLC39A2 | NA | NA | -1.2464 | **0.0241** | **0.0031** | **15** |
| SLC39A6 | -2.4223 | **1.31E-12** | 1.1639 | 0.2544 | 0.0000 | 1 |
| SLC39A7 | -1.1022 | 0.1385 | 1.6458 | **0.0347** | 0.0006 | 4 |
| SLC39A9 | -1.4036 | **4.22E-07** | NA | NA | 0.0002 | 8 |
| SLC52A2 | -1.5242 | **1.74E-07** | NA | NA | 0.0000 | 1 |
| SLC6A12 | -1.4641 | **0.0470** | -1.0135 | 0.9462 | 0.0000 | 2 |
| SLC6A20 | -1.5216 | **0.0382** | -1.0029 | 0.9887 | 0.0000 | 1 |
| SLC9A3R1 | -1.7651 | **0.0003** | 1.1164 | 0.5290 | 0.0011 | 5 |
| SLC9A3R2 | -1.2092 | **0.0068** | -1.1589 | 0.2052 | 0.0002 | 2 |
| SLIRP | -1.3688 | **0.0001** | -1.1997 | 0.1994 | 0.0000 | 3 |
| SLIT2 | 2.7856 | **4.28E-12** | 1.1015 | 0.6350 | 0.0000 | 1 |
| SLPI | -6.4518 | **1.65E-07** | 1.0586 | 0.8345 | 0.0002 | 3 |
| SMAD3 | -1.0856 | 0.3013 | -1.2551 | **0.0040** | **0.0070** | **19** |
| SMAD5 | -1.0849 | 0.2435 | 1.4224 | **0.0310** | 0.0006 | 4 |
| SMAD6 | 1.1995 | **0.0201** | -1.4429 | **0.0092** | 0.0000 | 2 |
| SMAD9 | -1.7290 | **8.71E-06** | 1.2844 | 0.1101 | 0.0000 | 1 |
| SMAP1 | -1.1833 | **0.0082** | 1.1087 | 0.3923 | 0.0001 | 4 |
| SMARCA2 | 1.5574 | **1.39E-05** | 1.2489 | **0.0340** | 0.0004 | 2 |
| SMARCA4 | -1.7465 | **4.28E-12** | NA | NA | 0.0018 | 6 |
| SMARCB1 | -1.3179 | **1.92E-05** | NA | NA | 0.0001 | 5 |
| SMARCC1 | -1.5997 | **2.00E-13** | NA | NA | 0.0024 | 7 |
| SMARCD1 | -1.1348 | **0.0059** | -1.1091 | 0.1673 | **0.0052** | **29** |
| SMARCD2 | -1.1884 | **0.0108** | 1.3010 | 0.0580 | 0.0000 | 1 |
| SMARCD3 | 1.4225 | **2.85E-06** | NA | NA | 0.0000 | 1 |
| SMARCE1 | -1.3014 | **1.34E-05** | 1.0194 | 0.8178 | 0.0013 | 13 |
| SMC2 | -1.5292 | **0.0004** | NA | NA | 0.0000 | 1 |
| SMNDC1 | -1.1362 | **0.0435** | 1.0295 | 0.8078 | 0.0000 | 1 |
| SMPD2 | -1.2972 | **0.0004** | NA | NA | 0.0000 | 3 |
| SMTN | 1.1640 | 0.2700 | -1.5028 | **0.0024** | 0.0000 | 1 |
| SMYD2 | -1.9354 | **9.21E-15** | 1.1131 | 0.2868 | 0.0000 | 1 |
| SMYD3 | -1.3718 | **0.0001** | -1.0461 | 0.6900 | 0.0000 | 1 |
| SMYD5 | -1.0189 | 0.7912 | -1.1877 | **0.0386** | 0.0000 | 1 |
| SNAI2 | 1.4245 | **0.0047** | -1.1919 | 0.2258 | 0.0001 | 4 |
| SNAPC3 | -1.2943 | **0.0040** | 1.2214 | 0.1886 | 0.0000 | 2 |
| SNCAIP | -1.4176 | **0.0006** | -1.0858 | 0.5061 | 0.0000 | 1 |
| SND1 | -1.3243 | **5.57E-07** | NA | NA | 0.0021 | 4 |
| SNF8 | -1.2457 | **6.44E-06** | NA | NA | 0.0007 | 8 |
| SNPH | 1.4798 | **8.85E-08** | -1.2041 | 0.1707 | 0.0000 | 1 |
| SNRNP25 | -1.5195 | **3.34E-08** | NA | NA | 0.0001 | 4 |
| SNRNP35 | -1.1663 | **0.0041** | 1.1619 | 0.0973 | 0.0002 | 2 |
| SNRPA1 | -1.3935 | **1.82E-08** | NA | NA | 0.0003 | 3 |
| SNRPB | -1.4542 | **1.66E-08** | NA | NA | **0.0028** | **18** |
| SNRPB2 | -1.1587 | **0.0179** | -1.0676 | 0.5095 | 0.0007 | 9 |
| SNRPC | -1.1694 | **0.0003** | -1.0764 | 0.2624 | 0.0017 | **14** |
| SNRPD2 | -1.2095 | **0.0004** | -1.0693 | 0.3525 | 0.0004 | 3 |
| SNRPF | -1.5048 | **1.46E-07** | -1.0511 | 0.6136 | 0.0008 | 7 |
| SNTA1 | 1.3357 | **2.43E-05** | -1.3350 | **0.0474** | **0.6667** | 2 |
| SNTB2 | 1.1716 | **0.0043** | 1.3188 | 0.0659 | 0.0000 | 1 |
| SNU13 | -1.1319 | **0.0018** | NA | NA | 0.0001 | 2 |
| SNX10 | 1.7439 | **0.0044** | -1.0543 | 0.8224 | 0.0000 | 2 |
| SNX11 | 1.1254 | **0.0145** | -1.0579 | 0.5409 | 0.0009 | 4 |
| SNX2 | 1.3114 | **0.0056** | NA | NA | 0.0013 | 5 |
| SNX4 | -1.2989 | **0.0001** | NA | NA | 0.0004 | 3 |
| SNX5 | 1.1563 | **0.0256** | -1.0083 | 0.9245 | 0.0000 | 2 |
| SNX6 | 1.1780 | **0.0038** | 1.0035 | 0.9790 | 0.0002 | 3 |
| SNX7 | -1.4172 | **0.0001** | NA | NA | 0.0003 | 3 |
| SOCS3 | 2.0808 | **0.0003** | -1.9138 | **0.0024** | **0.0043** | 7 |
| SOD1 | 1.2004 | **0.0001** | -1.1170 | **0.0416** | 0.0000 | 2 |
| SOD3 | 2.4538 | **1.48E-09** | 1.1410 | 0.3660 | 0.0001 | 2 |
| SORBS1 | 2.1979 | **1.68E-07** | -1.2641 | 0.0738 | 0.0002 | 4 |
| SORBS2 | 1.7489 | **2.19E-05** | 1.2173 | 0.1273 | **0.0031** | 11 |
| SORBS3 | 1.3363 | **9.02E-06** | -1.0499 | 0.7785 | **0.0073** | **37** |
| SORT1 | -1.2713 | **0.0143** | 1.0268 | 0.8837 | 0.0002 | 4 |
| SOX4 | -1.5523 | **0.0012** | NA | NA | 0.0006 | 3 |
| SOX5 | -1.2302 | **0.0285** | 1.3169 | **0.0092** | 0.0002 | 5 |
| SP1 | 1.0140 | 0.8414 | 1.1549 | **0.0500** | **0.0026** | 11 |
| SP100 | 1.8777 | **9.53E-12** | 1.3001 | **0.0097** | 0.0014 | 7 |
| SP3 | -1.1128 | 0.1649 | 1.3528 | **0.0254** | 0.0000 | 4 |
| SP4 | 1.2027 | **0.0406** | 1.1048 | 0.1607 | 0.0005 | 6 |
| SPA17 | NA | NA | 1.3884 | **0.0099** | 0.0000 | 1 |
| SPACA9 | 1.1882 | **0.0362** | NA | NA | 0.0000 | 1 |
| SPAG4 | -1.4064 | **0.0021** | 1.3077 | 0.1175 | 0.0004 | 6 |
| SPARC | NA | NA | -1.2668 | **0.0355** | 0.0000 | 1 |
| SPATA2L | -1.4010 | **0.0004** | -1.0231 | 0.8905 | 0.0000 | 1 |
| SPC25 | -2.7180 | **3.28E-07** | 1.3384 | 0.1965 | 0.0000 | 1 |
| SPDEF | -3.3107 | **0.0002** | 1.4088 | **0.0456** | 0.0000 | 2 |
| SPEG | 2.4475 | **2.43E-10** | -1.2572 | 0.2314 | 0.0000 | 1 |
| SPG7 | -1.2145 | **8.61E-06** | -1.0388 | 0.7354 | 0.0001 | 4 |
| SPICE1 | -1.2057 | **0.0055** | 1.1624 | 0.1159 | 0.0000 | 1 |
| SPINK2 | -1.4682 | 0.0578 | -1.5832 | **0.0156** | 0.0000 | 1 |
| SPOCK1 | -1.4264 | **0.0302** | -1.4018 | **0.0227** | 0.0000 | 1 |
| SPOP | 1.2436 | **0.0014** | 1.2024 | 0.1473 | 0.0014 | 4 |
| SPRED2 | 1.1930 | **0.0105** | 1.1004 | 0.2379 | 0.0005 | 7 |
| SPSB1 | 1.7200 | **0.0001** | -1.0418 | 0.7915 | 0.0025 | 5 |
| SPTAN1 | 1.7508 | **2.14E-17** | NA | NA | 0.0001 | 4 |
| SPTBN1 | 1.7972 | **1.17E-13** | NA | NA | 0.0001 | 3 |
| SQLE | -1.7378 | **1.41E-05** | 1.0898 | 0.6612 | 0.0000 | 2 |
| SREK1IP1 | 1.2159 | **0.0288** | NA | NA | 0.0020 | 4 |
| SRGN | 2.2608 | **1.12E-06** | -1.1617 | 0.3658 | 0.0004 | 2 |
| SRI | -1.3434 | **3.71E-07** | 1.0947 | 0.1810 | 0.0006 | 3 |
| SRPK1 | -1.5077 | **3.77E-11** | 1.1277 | 0.3780 | 0.0008 | 3 |
| SRPK2 | 1.0229 | 0.8004 | 1.2289 | **0.0217** | **0.0036** | **16** |
| SRPRB | -1.5094 | **2.05E-08** | NA | NA | 0.0000 | 1 |
| SRRM1 | -1.0365 | 0.5406 | 1.3973 | **0.0109** | 0.0000 | 2 |
| SRSF1 | -1.4759 | **1.09E-06** | 1.0660 | 0.5346 | 0.0013 | 8 |
| SRSF10 | -1.3409 | **7.64E-06** | NA | NA | 0.0000 | 2 |
| SRSF11 | 1.2466 | **0.0191** | NA | NA | 0.0000 | 3 |
| SRSF2 | -1.5474 | **1.87E-09** | -1.0982 | 0.4204 | 0.0001 | 2 |
| SRSF3 | -1.3146 | **1.41E-05** | -1.0553 | 0.6738 | 0.0012 | 9 |
| SRSF5 | 1.3819 | **0.0012** | NA | NA | 0.0000 | 1 |
| SRSF6 | -1.2686 | **0.0012** | 1.3150 | **0.0391** | 0.0000 | 1 |
| SRSF8 | 1.1005 | **0.0279** | -1.0515 | 0.5410 | 0.0000 | 3 |
| SRSF9 | -1.3058 | **3.61E-08** | NA | NA | 0.0002 | 2 |
| SS18L1 | -1.2394 | **0.0076** | 1.0241 | 0.8181 | 0.0004 | 10 |
| SSNA1 | -1.2051 | **0.0040** | NA | NA | 0.0000 | 3 |
| SSRP1 | -1.2641 | **1.12E-06** | 1.0410 | 0.6446 | 0.0000 | 1 |
| SSX2IP | -1.5450 | **0.0007** | NA | NA | **0.0060** | **35** |
| ST13 | 1.5114 | **2.62E-05** | 1.1885 | 0.0616 | 0.0000 | 1 |
| STAM2 | -1.2077 | **0.0284** | NA | NA | 0.0016 | 9 |
| STAMBP | -1.2910 | **1.26E-05** | 1.1512 | 0.1534 | 0.0008 | 7 |
| STAP2 | -1.3163 | **0.0005** | 1.1691 | 0.1731 | 0.0001 | 3 |
| STARD3 | 1.2447 | **2.10E-05** | -1.1546 | 0.1260 | 0.0000 | 1 |
| STAT5B | 1.4023 | **4.42E-06** | -1.3640 | **0.0021** | **0.0058** | 11 |
| STAT6 | 1.1729 | **0.0230** | 1.0078 | 0.9435 | 0.0000 | 1 |
| STEAP3 | -1.3010 | **0.0314** | 1.0723 | 0.6088 | 0.0000 | 1 |
| STIM1 | -1.4341 | **1.24E-06** | -1.2476 | 0.1078 | 0.0001 | 2 |
| STIP1 | -1.3722 | **4.13E-06** | NA | NA | 0.0005 | 3 |
| STK16 | -1.2132 | **0.0047** | NA | NA | **0.0040** | **23** |
| STK24 | -1.1957 | **0.0083** | -1.0349 | 0.7336 | 0.0000 | 1 |
| STK25 | -1.1953 | **0.0013** | NA | NA | 0.0000 | 2 |
| STK38 | 1.2286 | **0.0132** | 1.0388 | 0.6913 | 0.0000 | 1 |
| STK39 | -1.6472 | **0.0002** | 1.0315 | 0.8723 | 0.0000 | 1 |
| STMN1 | -1.6296 | **5.65E-07** | 1.0448 | 0.7202 | 0.0000 | 1 |
| STMN3 | 1.1972 | **0.0264** | NA | NA | 0.0002 | 8 |
| STOM | 2.0410 | **9.68E-13** | -1.1854 | **0.0366** | **0.0191** | **21** |
| STRAP | -1.4761 | **1.55E-08** | -1.0114 | 0.9254 | 0.0000 | 1 |
| STRN | 1.1484 | **0.0380** | NA | NA | 0.0003 | 3 |
| STRN4 | -1.1530 | **0.0118** | -1.4078 | **0.0353** | 0.0000 | 1 |
| STUB1 | 1.1531 | **0.0010** | 1.0747 | 0.6003 | **0.0045** | 13 |
| STX11 | NA | NA | -1.5315 | **0.0144** | **0.0064** | **24** |
| STX12 | 1.2898 | **2.48E-05** | 1.0354 | 0.7045 | **0.0031** | 10 |
| STX17 | -1.1407 | **0.0271** | NA | NA | 0.0000 | 1 |
| STX1A | -1.3037 | **0.0044** | -1.1304 | 0.1820 | **0.0112** | **32** |
| STX2 | 1.3009 | **2.06E-06** | 1.3049 | **0.0444** | **0.0066** | **21** |
| STX3 | 1.0227 | 0.8135 | -1.2709 | **0.0481** | 0.0006 | **14** |
| STX4 | 1.1292 | **0.0303** | NA | NA | **0.0064** | **20** |
| STX5 | -1.1571 | **0.0051** | -1.0347 | 0.7068 | 0.0024 | 13 |
| STX7 | 1.5215 | **2.54E-08** | NA | NA | 0.0011 | 12 |
| STX8 | 1.1207 | **0.0116** | 1.1252 | 0.2755 | 0.0020 | **18** |
| SUCLA2 | -1.2135 | **0.0233** | NA | NA | 0.0000 | 2 |
| SUGP2 | -1.3004 | **2.75E-05** | 1.1492 | 0.3258 | 0.0000 | 1 |
| SUMO3 | NA | NA | -1.2146 | **0.0290** | 0.0009 | 3 |
| SUN2 | 1.2101 | **0.0030** | 1.0582 | 0.6689 | 0.0001 | 3 |
| SUOX | -1.1369 | **0.0209** | NA | NA | **0.0057** | **17** |
| SUPT20H | -1.2202 | **0.0001** | NA | NA | 0.0000 | 1 |
| SUPT4H1 | 1.2091 | **0.0004** | 1.0603 | 0.3883 | 0.0000 | 1 |
| SUPT5H | 1.0388 | 0.4513 | -1.1925 | **0.0381** | 0.0011 | 3 |
| SUPT6H | 1.1059 | **0.0280** | NA | NA | 0.0000 | 1 |
| SUSD6 | 1.5010 | **2.42E-05** | NA | NA | 0.0001 | 2 |
| SUV39H1 | -1.4537 | **1.51E-05** | 1.0551 | 0.6912 | 0.0008 | 6 |
| SWAP70 | 1.5243 | **0.0001** | NA | NA | 0.0000 | 1 |
| SYBU | NA | NA | 1.5749 | **0.0351** | 0.0000 | 1 |
| SYF2 | 1.3992 | **8.45E-06** | NA | NA | 0.0000 | 3 |
| SYNCRIP | -1.4648 | **1.35E-07** | 1.1709 | 0.0712 | 0.0005 | 5 |
| SYNE1 | 1.6755 | **5.84E-07** | 1.1314 | 0.1853 | 0.0004 | 3 |
| SYNE2 | NA | NA | 1.4472 | **0.0260** | 0.0001 | 3 |
| SYNGR1 | 1.5765 | **6.94E-07** | -1.1976 | 0.0966 | 0.0011 | 7 |
| SYNGR3 | 1.2826 | **0.0388** | -1.3757 | 0.1435 | 0.0002 | 4 |
| SYNJ1 | 1.2787 | **0.0002** | 1.1308 | 0.2089 | 0.0000 | 1 |
| SYNJ2 | 1.4373 | **4.76E-05** | 1.0056 | 0.9592 | 0.0000 | 1 |
| SYNPO2 | 2.1665 | **7.15E-10** | NA | NA | 0.0000 | 2 |
| SYNRG | -1.1412 | **0.0154** | -1.1733 | **0.0201** | 0.0001 | 2 |
| SYT2 | -1.5333 | **0.0049** | -1.1434 | 0.5309 | 0.0001 | 6 |
| SZT2 | 1.0020 | 0.9770 | -1.2501 | **0.0040** | 0.0000 | 2 |
| TACC1 | 2.0458 | **4.50E-15** | -1.0351 | 0.8645 | 0.0019 | 9 |
| TACSTD2 | -2.5208 | **4.59E-06** | 1.1176 | 0.6003 | 0.0000 | 1 |
| TADA2A | -1.3243 | **1.51E-06** | 1.0020 | 0.9843 | 0.0007 | 13 |
| TAF1 | -1.1370 | **0.0367** | 1.2144 | **0.0371** | 0.0005 | 10 |
| TAF12 | -1.1032 | 0.0608 | -1.1661 | **0.0493** | 0.0000 | 1 |
| TAF1B | -1.4811 | **2.08E-07** | NA | NA | 0.0009 | 4 |
| TAF1D | -1.2560 | **0.0036** | NA | NA | 0.0000 | 3 |
| TAF4 | -1.1518 | **0.0152** | -1.0489 | 0.5883 | 0.0009 | 3 |
| TAF6L | -1.1505 | **0.0090** | -1.1856 | **0.0336** | 0.0001 | 3 |
| TAF9 | -1.3611 | **1.75E-05** | NA | NA | 0.0017 | 8 |
| TAGLN | 6.3363 | **4.63E-16** | NA | NA | 0.0001 | 2 |
| TAL1 | NA | NA | -1.3211 | **0.0032** | 0.0003 | 5 |
| TALDO1 | -1.3227 | **0.0001** | 1.0989 | 0.3780 | 0.0000 | 1 |
| TAP1 | -1.5443 | **6.76E-06** | 1.0805 | 0.6396 | 0.0000 | 2 |
| TARBP2 | -1.2493 | **0.0007** | NA | NA | 0.0004 | 7 |
| TARS | -1.2356 | **0.0029** | NA | NA | 0.0000 | 1 |
| TARS2 | -1.1237 | **0.0365** | 1.1644 | 0.2880 | 0.0002 | 3 |
| TBC1D1 | 1.1747 | **0.0143** | -1.0841 | 0.4262 | 0.0023 | 11 |
| TBC1D30 | -1.6850 | **1.49E-09** | 1.0415 | 0.6686 | 0.0000 | 2 |
| TBC1D5 | 1.1676 | **0.0030** | 1.1556 | 0.3192 | 0.0008 | 3 |
| TBCB | -1.0993 | **0.0378** | -1.0791 | 0.4879 | 0.0000 | 1 |
| TBK1 | 1.1898 | **0.0114** | NA | NA | 0.0004 | 3 |
| TBL1X | -1.4716 | **2.91E-06** | -1.2863 | **0.0401** | 0.0000 | 1 |
| TBL1XR1 | 1.0823 | 0.5327 | 1.2588 | **0.0360** | 0.0000 | 1 |
| TBP | -1.1849 | **0.0015** | -1.0146 | 0.8613 | 0.0011 | 7 |
| TBRG4 | -1.2178 | **0.0020** | 1.0991 | 0.3798 | 0.0004 | 4 |
| TBX19 | 1.2742 | **0.0027** | NA | NA | 0.0000 | 4 |
| TBX3 | -2.3866 | **3.89E-11** | -1.3198 | **0.0270** | 0.0002 | 3 |
| TBXA2R | 1.4457 | **0.0001** | -1.2406 | **0.0273** | **0.0037** | 10 |
| TCAP | 1.1855 | **0.0251** | -1.1218 | 0.4182 | **0.0026** | 8 |
| TCEA2 | 1.2118 | **0.0006** | NA | NA | **0.0090** | **33** |
| TCERG1 | -1.3422 | **0.0067** | 1.4061 | **0.0440** | 0.0003 | 3 |
| TCF12 | -1.7876 | **6.46E-09** | NA | NA | 0.0015 | **20** |
| TCF20 | -1.2461 | **0.0001** | NA | NA | 0.0000 | 1 |
| TCF3 | -1.1910 | **0.0009** | NA | NA | 0.0005 | 6 |
| TCF4 | 1.4306 | **0.0032** | 1.2650 | 0.1932 | **0.0212** | **53** |
| TCF7L2 | 1.3587 | **0.0007** | -1.0197 | 0.8922 | 0.0009 | 8 |
| TCP1 | -1.2414 | **9.98E-06** | -1.0374 | 0.5439 | 0.0000 | 1 |
| TCTA | -1.1869 | **0.0191** | 1.0785 | 0.5680 | 0.0000 | 2 |
| TDG | -1.2802 | **0.0006** | NA | NA | 0.0000 | 1 |
| TDRD3 | 1.3032 | **0.0101** | NA | NA | 0.0000 | 1 |
| TDRD7 | -1.1718 | **0.0140** | 1.1050 | 0.2302 | 0.0000 | 2 |
| TEAD3 | 1.2769 | **0.0016** | -1.6472 | **0.0007** | 0.0008 | 3 |
| TEK | 1.2929 | **0.0013** | -1.1049 | 0.4354 | 0.0002 | 2 |
| TELO2 | -1.2381 | **0.0001** | -1.0417 | 0.6149 | 0.0000 | 1 |
| TENT5C | 1.4085 | **0.0111** | NA | NA | 0.0000 | 2 |
| TERF1 | 1.3915 | **0.0004** | NA | NA | 0.0010 | 9 |
| TERF2 | -1.1262 | **0.0248** | -1.0206 | 0.8974 | 0.0006 | 6 |
| TERF2IP | 1.1992 | **0.0004** | NA | NA | 0.0000 | 3 |
| TF | 1.2695 | **0.0444** | -1.1584 | **0.0247** | 0.0003 | 7 |
| TFAP4 | -1.1767 | **0.0244** | -1.0809 | 0.2063 | 0.0000 | 4 |
| TFCP2 | -1.2076 | **7.14E-06** | 1.1205 | 0.0999 | **0.0082** | **17** |
| TFDP1 | -1.3720 | **6.29E-06** | 1.0144 | 0.8944 | 0.0001 | 2 |
| TFG | -1.2323 | **0.0008** | 1.1174 | 0.3551 | **0.0033** | **18** |
| TFIP11 | -1.1335 | **0.0106** | NA | NA | **0.0234** | **68** |
| TGFA | -1.2323 | **0.0353** | 1.0729 | 0.3192 | 0.0000 | 2 |
| TGFB1I1 | 1.5686 | **1.69E-05** | NA | NA | 0.0000 | 1 |
| TGFBR2 | 2.5560 | **6.72E-16** | 1.1455 | 0.3666 | 0.0001 | 3 |
| TGIF1 | -1.5517 | **3.25E-06** | 1.0774 | 0.4007 | 0.0000 | 2 |
| TGM2 | -1.7220 | **0.0002** | 1.0452 | 0.8147 | 0.0001 | 2 |
| TGS1 | -1.1571 | **0.0010** | 1.0759 | 0.5178 | 0.0000 | 1 |
| THAP11 | 1.2205 | **0.0007** | 1.0299 | 0.7485 | 0.0016 | 3 |
| THAP4 | -1.3368 | **2.25E-05** | 1.1469 | 0.3707 | 0.0011 | 8 |
| THAP7 | 1.2059 | **0.0009** | 1.1577 | 0.2055 | **0.0052** | **14** |
| THBD | 2.2409 | **1.83E-05** | -1.2446 | 0.2792 | 0.0008 | 7 |
| THBS2 | 4.8280 | **5.77E-13** | NA | NA | 0.0000 | 1 |
| THRA | 1.4622 | **1.52E-06** | NA | NA | 0.0014 | 10 |
| THYN1 | 1.1208 | **0.0376** | 1.0555 | 0.4385 | 0.0000 | 1 |
| TIA1 | -1.2179 | **0.0129** | NA | NA | 0.0000 | 1 |
| TIE1 | 2.0219 | **3.88E-08** | NA | NA | 0.0000 | 2 |
| TIMM17B | -1.1981 | **0.0001** | -1.1040 | 0.4007 | 0.0000 | 2 |
| TIMM23 | -1.3504 | **2.73E-05** | NA | NA | 0.0003 | 4 |
| TIMM50 | -1.2462 | **0.0001** | 1.0430 | 0.6252 | 0.0000 | 1 |
| TIMM8A | -1.2212 | **0.0339** | 1.0825 | 0.5032 | 0.0001 | 3 |
| TIMP4 | 1.9131 | **0.0003** | -1.2150 | 0.1179 | 0.0000 | 1 |
| TINAGL1 | 1.3971 | **0.0007** | -1.1174 | 0.1713 | 0.0000 | 1 |
| TIPIN | -1.3710 | **0.0063** | 1.1000 | 0.5349 | 0.0000 | 1 |
| TLE3 | 1.4358 | **0.0004** | NA | NA | 0.0000 | 1 |
| TLE4 | 1.2447 | **0.0204** | -1.0158 | 0.8986 | 0.0005 | 3 |
| TLK1 | 1.0273 | 0.7685 | 1.3808 | **0.0034** | 0.0008 | 2 |
| TLN1 | 1.6604 | **3.55E-10** | NA | NA | 0.0000 | 1 |
| TLX3 | NA | NA | 1.1550 | **0.0409** | 0.0017 | **14** |
| TM2D1 | 1.1222 | **0.0089** | -1.0091 | 0.9486 | 0.0000 | 1 |
| TMBIM1 | 1.3731 | **0.0039** | 1.3277 | 0.0669 | 0.0000 | 1 |
| TMBIM6 | -1.3030 | **7.16E-06** | 1.0769 | 0.2610 | **0.0046** | 12 |
| TMCC2 | 1.5828 | **9.41E-06** | NA | NA | 0.0000 | 5 |
| TMED2 | -1.7181 | **5.98E-07** | NA | NA | 0.0000 | 2 |
| TMED3 | -1.2812 | **0.0029** | NA | NA | 0.0000 | 2 |
| TMED9 | -1.3673 | **5.86E-06** | 1.0569 | 0.7524 | 0.0008 | 2 |
| TMEM100 | 3.2037 | **4.67E-09** | 1.0831 | 0.7400 | 0.0011 | 8 |
| TMEM109 | 1.4916 | **1.38E-11** | NA | NA | 0.0003 | 5 |
| TMEM147 | -1.1373 | **0.0219** | NA | NA | 0.0016 | 11 |
| TMEM159 | 2.3036 | **1.54E-10** | 1.2105 | 0.2509 | 0.0000 | 1 |
| TMEM208 | -1.3166 | **2.83E-05** | -1.1233 | 0.0545 | 0.0006 | 6 |
| TMEM243 | 1.3748 | **2.80E-05** | NA | NA | 0.0002 | 8 |
| TMEM254 | -1.4968 | **1.65E-08** | NA | NA | 0.0009 | 10 |
| TMEM258 | -1.2174 | **0.0027** | NA | NA | 0.0001 | 2 |
| TMEM267 | -1.2799 | **0.0294** | NA | NA | 0.0000 | 2 |
| TMEM35A | -1.4414 | **0.0112** | NA | NA | 0.0002 | 6 |
| TMEM43 | 1.1720 | **0.0054** | -1.1111 | 0.2292 | 0.0006 | 2 |
| TMEM45A | -1.2499 | **0.0473** | -1.0939 | 0.4682 | 0.0000 | 2 |
| TMEM50A | -1.1023 | 0.1615 | 1.2092 | **0.0239** | 0.0000 | 1 |
| TMEM50B | 1.1482 | **0.0233** | 1.0031 | 0.9701 | 0.0008 | 5 |
| TMEM8A | -1.7569 | **2.76E-07** | -1.0305 | 0.8064 | 0.0000 | 1 |
| TMEM97 | -1.3944 | **0.0017** | 1.1975 | 0.2446 | 0.0014 | 13 |
| TMF1 | -1.1388 | 0.0916 | 1.2932 | **0.0337** | 0.0008 | 2 |
| TMPO | -1.4989 | **3.98E-05** | 1.1256 | 0.3929 | 0.0003 | 3 |
| TMX1 | -1.3010 | **0.0001** | NA | NA | 0.0002 | 4 |
| TMX2 | -1.1120 | **0.0150** | -1.0719 | 0.4889 | **0.0102** | **52** |
| TNFAIP1 | -1.1149 | **0.0454** | NA | NA | 0.0010 | 9 |
| TNFAIP3 | 2.1549 | **6.93E-07** | -1.0432 | 0.8016 | **0.0029** | 10 |
| TNFAIP8 | 2.1927 | **1.99E-07** | -1.1954 | 0.4842 | 0.0001 | 3 |
| TNFRSF14 | 1.4344 | **2.34E-05** | -1.0212 | 0.8921 | 0.0000 | 1 |
| TNFRSF1A | 1.5560 | **2.35E-09** | NA | NA | 0.0002 | 5 |
| TNFRSF21 | -1.5758 | **0.0003** | NA | NA | 0.0001 | 2 |
| TNIP2 | -1.1596 | **0.0039** | NA | NA | 0.0010 | 3 |
| TNKS2 | 1.3166 | **0.0112** | NA | NA | 0.0001 | 5 |
| TNNC2 | 1.4366 | **0.0004** | -1.1496 | 0.1643 | 0.0000 | 1 |
| TNPO1 | 1.2228 | **0.0002** | NA | NA | 0.0000 | 3 |
| TNPO3 | -1.1177 | **0.0272** | NA | NA | 0.0010 | 4 |
| TNRC6B | 1.1950 | **0.0040** | 1.3847 | **0.0287** | 0.0000 | 2 |
| TOB1 | NA | NA | 1.6286 | **0.0144** | 0.0001 | 2 |
| TOB2 | NA | NA | 1.1987 | **0.0274** | 0.0007 | 3 |
| TOLLIP | 1.1614 | **0.0127** | -1.3052 | 0.1444 | 0.0022 | 10 |
| TOM1L1 | -3.2366 | **1.22E-17** | NA | NA | 0.0002 | 2 |
| TOM1L2 | 1.0307 | 0.6349 | -1.2947 | **0.0011** | 0.0000 | 2 |
| TOP1 | -1.2451 | **0.0002** | 1.3508 | **0.0057** | 0.0008 | 4 |
| TOP2B | -1.2490 | **0.0042** | NA | NA | 0.0000 | 2 |
| TOP3A | -1.1772 | **0.0134** | -1.1436 | 0.2168 | 0.0008 | 2 |
| TOP3B | -1.2033 | **0.0029** | NA | NA | 0.0001 | 5 |
| TOPBP1 | -1.1911 | **0.0346** | 1.5058 | **0.0114** | 0.0001 | 3 |
| TOPORS | 1.1560 | 0.0980 | 1.2904 | **0.0410** | 0.0010 | 5 |
| TOR1AIP1 | 1.3745 | **3.90E-05** | 1.1754 | 0.1516 | 0.0000 | 1 |
| TOR1AIP2 | -1.1536 | **0.0259** | 1.0017 | 0.9849 | 0.0000 | 1 |
| TOX3 | -1.9402 | **0.0011** | -1.3791 | 0.2544 | 0.0008 | 3 |
| TOX4 | 1.0397 | 0.4170 | -1.1121 | **0.0489** | 0.0000 | 1 |
| TPBG | -1.6736 | **0.0001** | 1.0939 | 0.4642 | 0.0000 | 1 |
| TPD52 | -6.0407 | **2.00E-23** | 1.3982 | 0.1248 | 0.0001 | 2 |
| TPI1 | -1.7171 | **2.76E-09** | 1.1452 | 0.2646 | 0.0001 | 2 |
| TPK1 | 1.5921 | **1.11E-05** | -1.0724 | 0.5057 | 0.0000 | 1 |
| TPM1 | 2.1128 | **6.81E-08** | -1.3144 | **0.0227** | 0.0019 | 4 |
| TPM2 | 2.6953 | **5.88E-10** | -1.3662 | **0.0184** | 0.0000 | 1 |
| TPM3 | -1.1876 | **0.0139** | 1.0596 | 0.5551 | 0.0010 | 6 |
| TPR | 1.1377 | 0.0828 | 1.3798 | **0.0083** | 0.0000 | 1 |
| TPT1 | 1.1622 | **0.0015** | -1.0495 | 0.3316 | 0.0000 | 1 |
| TRA2A | -1.3068 | **1.92E-06** | 1.2074 | 0.2040 | 0.0000 | 1 |
| TRA2B | -1.3696 | **2.47E-07** | -1.1410 | 0.0698 | 0.0005 | 6 |
| TRAF1 | 1.1841 | **0.0410** | -1.0767 | 0.4636 | **0.0127** | **48** |
| TRAF3IP1 | -1.1281 | **0.0239** | 1.1439 | 0.1707 | 0.0000 | 1 |
| TRAF3IP2 | NA | NA | 1.2802 | **0.0476** | 0.0001 | 4 |
| TRAF5 | 1.3925 | **0.0008** | NA | NA | **0.0063** | **15** |
| TRAIP | -1.3840 | **0.0003** | NA | NA | 0.0005 | 6 |
| TRAK1 | -1.7190 | **2.72E-09** | NA | NA | 0.0000 | 1 |
| TRAM1 | -1.3369 | **0.0008** | 1.0472 | 0.6332 | 0.0000 | 1 |
| TRAP1 | -1.4543 | **1.35E-09** | NA | NA | 0.0016 | 3 |
| TRAPPC4 | -1.2666 | **3.69E-06** | -1.1177 | 0.1154 | 0.0000 | 1 |
| TRAPPC6A | -1.4895 | **4.76E-08** | NA | NA | 0.0025 | 11 |
| TRIAP1 | -1.2382 | **0.0003** | -1.0146 | 0.8849 | 0.0000 | 1 |
| TRIM13 | -1.1467 | **0.0167** | 1.0986 | 0.3393 | 0.0000 | 1 |
| TRIM2 | NA | NA | 1.4845 | **0.0086** | 0.0001 | 4 |
| TRIM21 | 1.3302 | **9.36E-06** | -1.1387 | 0.3123 | **0.0026** | 10 |
| TRIM22 | 1.2607 | **0.0350** | -1.0967 | 0.5206 | 0.0000 | 1 |
| TRIM23 | 1.1707 | 0.0574 | 1.1989 | **0.0291** | **0.0124** | **45** |
| TRIM26 | -1.3510 | **3.19E-07** | NA | NA | 0.0000 | 1 |
| TRIM27 | -1.4822 | **8.26E-10** | NA | NA | **0.0489** | **99** |
| TRIM32 | -1.1745 | **0.0065** | -1.0372 | 0.6895 | **0.0061** | 13 |
| TRIM45 | -1.3941 | **2.89E-05** | 1.0468 | 0.5717 | 0.0002 | 2 |
| TRIM5 | -1.5459 | **3.48E-05** | 1.2407 | 0.1316 | 0.0007 | 7 |
| TRIO | 1.3902 | **2.64E-06** | 1.2277 | 0.1374 | 0.0000 | 1 |
| TRIOBP | -1.1619 | **0.0390** | -1.1938 | 0.1528 | 0.0000 | 1 |
| TRIP10 | 1.3529 | **0.0001** | NA | NA | 0.0022 | 11 |
| TRIP4 | -1.1593 | **0.0125** | NA | NA | 0.0002 | 3 |
| TRIP6 | -1.2232 | **0.0006** | NA | NA | **0.0089** | **36** |
| TROAP | -2.3822 | **6.93E-07** | 1.1096 | 0.4786 | 0.0003 | 5 |
| TRPC1 | 2.8812 | **8.51E-14** | NA | NA | 0.0008 | 2 |
| TRPC4 | NA | NA | -1.4833 | **0.0097** | 0.0000 | 1 |
| TRPC6 | -1.3627 | **0.0290** | -1.0784 | 0.6109 | 0.0000 | 1 |
| TSC1 | 1.1338 | **0.0172** | NA | NA | **0.0057** | **18** |
| TSC2 | -1.4245 | **0.0001** | 1.0194 | 0.8991 | 0.0013 | 5 |
| TSC22D3 | 3.3872 | **8.96E-17** | 1.2494 | 0.0928 | 0.0001 | 2 |
| TSG101 | -1.1420 | **0.0039** | -1.0106 | 0.8936 | **0.0034** | **16** |
| TSHZ2 | 1.5154 | **0.0001** | 1.0763 | 0.6596 | 0.0001 | 5 |
| TSKS | 1.2617 | **0.0453** | -1.1848 | 0.1184 | 0.0000 | 1 |
| TSN | -1.2344 | **0.0005** | 1.0165 | 0.8710 | 0.0007 | 6 |
| TSNAX | 1.3877 | **0.0007** | 1.4394 | **0.0085** | 0.0011 | 13 |
| TSNAXIP1 | -1.1952 | **0.0085** | 1.1168 | 0.2294 | 0.0000 | 1 |
| TSPAN12 | -2.5113 | **1.18E-08** | NA | NA | 0.0015 | 9 |
| TSPAN14 | -1.4268 | **4.93E-08** | NA | NA | 0.0000 | 1 |
| TSPAN15 | -1.9211 | **0.0001** | 1.1049 | 0.5610 | 0.0000 | 1 |
| TSPAN4 | 3.0239 | **5.65E-22** | -1.0260 | 0.8521 | 0.0002 | 5 |
| TSPYL1 | 1.3809 | **4.65E-06** | -1.0111 | 0.9106 | 0.0003 | 5 |
| TSPYL2 | 1.5359 | **1.20E-07** | NA | NA | 0.0017 | 10 |
| TSR1 | -1.5069 | **8.85E-08** | -1.0027 | 0.9785 | 0.0000 | 1 |
| TSTD2 | -1.1743 | **0.0143** | NA | NA | 0.0011 | 8 |
| TTC1 | 1.1082 | **0.0126** | 1.1426 | 0.2892 | 0.0008 | 2 |
| TTC12 | -1.2873 | **0.0001** | 1.0520 | 0.5732 | 0.0000 | 1 |
| TTC19 | -1.4049 | **4.21E-06** | NA | NA | 0.0017 | 13 |
| TTLL12 | -1.4343 | **0.0003** | 1.0111 | 0.9435 | 0.0000 | 1 |
| TTLL7 | 1.5738 | **0.0001** | NA | NA | 0.0000 | 1 |
| TTN | 1.1970 | **0.0092** | NA | NA | 0.0013 | 7 |
| TUBB | -1.4213 | **3.71E-05** | NA | NA | 0.0023 | 4 |
| TUBGCP4 | -1.3294 | **5.38E-08** | 1.1031 | 0.3084 | 0.0018 | 10 |
| TXK | -2.0805 | **6.76E-07** | 1.0090 | 0.9511 | **0.0026** | 9 |
| TXN | -2.4676 | **6.54E-16** | -1.0673 | 0.6119 | 0.0018 | 8 |
| TXNDC15 | 1.2202 | **0.0005** | NA | NA | 0.0000 | 1 |
| TXNDC9 | -1.5487 | **1.47E-07** | 1.1344 | 0.2586 | 0.0015 | 9 |
| TXNRD2 | 1.0874 | 0.0777 | -1.1254 | **0.0407** | 0.0000 | 2 |
| U2AF2 | -1.1267 | **0.0240** | -1.4791 | **0.0265** | **0.0030** | 13 |
| UBA1 | -1.3833 | **4.38E-08** | -1.0678 | 0.6075 | 0.0002 | 2 |
| UBAP2L | -1.2275 | **7.11E-06** | -1.0543 | 0.3917 | 0.0001 | 2 |
| UBE2A | -1.3649 | **2.12E-07** | 1.0287 | 0.6983 | 0.0000 | 1 |
| UBE2B | 1.2934 | **0.0019** | 1.2722 | **0.0206** | 0.0000 | 1 |
| UBE2D1 | -1.3818 | **0.0016** | 1.1158 | 0.4450 | **0.0071** | **25** |
| UBE2D2 | -1.5706 | **1.06E-10** | -1.0301 | 0.8522 | **0.0077** | **21** |
| UBE2E3 | -1.3637 | **0.0001** | 1.1462 | 0.1625 | 0.0011 | 10 |
| UBE2G1 | -1.2553 | **0.0001** | 1.1403 | 0.1101 | 0.0013 | 2 |
| UBE2H | -1.4818 | **3.18E-11** | 1.2240 | **0.0262** | 0.0000 | 1 |
| UBE2I | 1.2871 | **0.0001** | NA | NA | **0.0825** | **95** |
| UBE2J1 | -1.3738 | **1.18E-05** | 1.0923 | 0.3832 | 0.0008 | 9 |
| UBE2K | -1.3303 | **6.85E-06** | 1.1881 | **0.0235** | **0.0037** | 13 |
| UBE2M | -1.2441 | **0.0020** | NA | NA | 0.0003 | 2 |
| UBE2N | -1.5161 | **1.16E-11** | -1.0279 | 0.7192 | **0.0043** | 13 |
| UBE2Q1 | 1.1439 | **0.0082** | 1.0685 | 0.4553 | 0.0000 | 1 |
| UBE2Z | -1.3233 | **4.67E-09** | NA | NA | 0.0005 | 4 |
| UBE3A | -1.1796 | **0.0199** | NA | NA | **0.0028** | 9 |
| UBE3C | -1.2161 | **0.0003** | 1.2149 | 0.0748 | 0.0000 | 1 |
| UBIAD1 | -1.1662 | **0.0064** | NA | NA | 0.0009 | 10 |
| UBL5 | -1.1926 | **0.0010** | -1.0879 | 0.1731 | 0.0015 | 10 |
| UBQLN2 | 1.3194 | **4.64E-05** | -1.0899 | 0.4435 | **0.0350** | **67** |
| UBR2 | 1.1018 | **0.0443** | NA | NA | 0.0000 | 2 |
| UBR5 | 1.0338 | 0.6802 | 1.3304 | **0.0451** | 0.0001 | 2 |
| UBTD1 | 1.3983 | **0.0001** | -1.3918 | 0.1261 | 0.0000 | 2 |
| UBTF | -1.1769 | **0.0065** | NA | NA | 0.0002 | 3 |
| UBXN1 | 1.1413 | **0.0051** | NA | NA | 0.0008 | 6 |
| UBXN4 | -1.2044 | **0.0197** | NA | NA | 0.0000 | 1 |
| UBXN6 | 1.1885 | **0.0132** | NA | NA | 0.0001 | 3 |
| UCP3 | -1.1432 | **0.0347** | 1.1047 | 0.6560 | 0.0000 | 1 |
| UFD1 | -1.1764 | **0.0011** | NA | NA | 0.0003 | 2 |
| UFSP2 | 1.4004 | **9.44E-08** | NA | NA | 0.0006 | 2 |
| UGP2 | 1.5614 | **3.69E-06** | NA | NA | 0.0002 | 3 |
| UNC13B | -1.3554 | **8.99E-08** | -1.0408 | 0.6560 | 0.0000 | 1 |
| UNG | -1.2003 | **0.0115** | NA | NA | 0.0000 | 1 |
| UNKL | 1.4319 | **1.68E-07** | -1.0798 | 0.4001 | 0.0010 | 9 |
| UPF1 | -1.2324 | **0.0003** | 1.0527 | 0.6089 | 0.0000 | 2 |
| UQCRB | 1.3036 | **0.0001** | NA | NA | 0.0000 | 1 |
| UQCRQ | -1.4103 | **6.38E-07** | -1.0141 | 0.8483 | 0.0000 | 1 |
| URB2 | -1.3617 | **3.00E-06** | NA | NA | 0.0000 | 1 |
| UROD | 1.3343 | **1.13E-07** | -1.2161 | 0.0791 | **0.0032** | 2 |
| USP10 | -1.4563 | **9.08E-07** | 1.1865 | 0.1482 | 0.0008 | 2 |
| USP15 | 1.3688 | **8.29E-07** | NA | NA | 0.0003 | 4 |
| USP2 | -1.0813 | 0.5577 | -1.2291 | **0.0322** | **0.0079** | **30** |
| USP33 | -1.0664 | 0.3001 | 1.2999 | **0.0212** | 0.0005 | 4 |
| USP4 | -1.1743 | **0.0001** | NA | NA | 0.0000 | 2 |
| USP47 | -1.2110 | **0.0404** | NA | NA | 0.0000 | 1 |
| USP5 | -1.1604 | **0.0057** | NA | NA | 0.0000 | 2 |
| UTP6 | -1.2341 | **0.0001** | 1.0222 | 0.8390 | 0.0000 | 1 |
| UVRAG | 1.5172 | **2.19E-09** | -1.0036 | 0.9694 | 0.0000 | 1 |
| UXT | 1.1255 | **0.0235** | -1.1307 | 0.0718 | 0.0010 | 4 |
| VAMP1 | 1.4352 | **6.65E-06** | 1.0744 | 0.2929 | 0.0002 | 10 |
| VAMP2 | 1.5196 | **1.18E-05** | 1.0674 | 0.7697 | **0.0026** | 9 |
| VAMP5 | 2.0877 | **7.43E-16** | NA | NA | 0.0021 | **18** |
| VAMP8 | -1.6820 | **0.0005** | 1.2152 | 0.1041 | 0.0000 | 1 |
| VANGL1 | -1.5883 | **3.75E-08** | NA | NA | 0.0000 | 1 |
| VARS | -1.2056 | **0.0079** | NA | NA | 0.0004 | 2 |
| VASP | 1.2294 | **0.0138** | NA | NA | **0.0035** | 9 |
| VAV1 | 1.5915 | **0.0002** | -1.0759 | 0.5457 | 0.0012 | 6 |
| VAV2 | NA | NA | 1.1736 | **0.0402** | 0.0000 | 2 |
| VBP1 | -1.4060 | **2.00E-05** | 1.1661 | 0.1316 | 0.0012 | 10 |
| VCAM1 | 13.5012 | **4.69E-24** | 1.0256 | 0.9250 | **0.1815** | **204** |
| VCP | -1.1267 | **0.0196** | NA | NA | **0.0066** | **15** |
| VDAC1 | -1.6297 | **1.17E-09** | 1.0504 | 0.6541 | 0.0008 | 2 |
| VEGFC | 1.4069 | **0.0003** | NA | NA | 0.0000 | 1 |
| VGLL3 | 2.5288 | **4.28E-07** | -1.0958 | 0.6859 | 0.0019 | 13 |
| VGLL4 | 1.4275 | **4.25E-07** | 1.0868 | 0.4240 | 0.0000 | 1 |
| VPS11 | 1.2947 | **7.79E-06** | -1.0895 | 0.4015 | 0.0000 | 2 |
| VPS37B | -1.7914 | **9.97E-08** | -1.0672 | 0.6996 | 0.0024 | **14** |
| VPS51 | 1.1694 | **0.0398** | NA | NA | 0.0000 | 1 |
| VPS52 | -1.1103 | **0.0186** | 1.0581 | 0.6718 | **0.0071** | **32** |
| VPS72 | -1.1312 | **0.0142** | NA | NA | 0.0002 | 3 |
| VRK1 | -1.4004 | **0.0008** | 1.1557 | 0.3140 | 0.0000 | 1 |
| VSIG4 | 4.0184 | **7.77E-15** | -1.1330 | 0.3237 | 0.0000 | 1 |
| VSTM4 | 1.4559 | **0.0024** | NA | NA | 0.0000 | 1 |
| VTN | 2.6600 | **1.35E-06** | -1.4334 | 0.0596 | 0.0000 | 2 |
| VWA5A | 1.3502 | **0.0001** | 1.2478 | **0.0372** | 0.0000 | 1 |
| VWF | 2.9188 | **7.41E-17** | NA | NA | 0.0014 | 4 |
| WARS | -1.5309 | **3.98E-06** | -1.0865 | 0.5611 | 0.0000 | 1 |
| WASHC3 | 1.4182 | **1.10E-05** | NA | NA | **0.0029** | **16** |
| WASL | 1.6066 | **6.43E-07** | 1.3069 | **0.0032** | **0.0028** | 12 |
| WBP2 | 1.3493 | **0.0001** | 1.0549 | 0.6293 | 0.0002 | 5 |
| WDFY3 | -1.4765 | **6.40E-07** | 1.0773 | 0.4618 | **0.0030** | 4 |
| WDR4 | -1.3909 | **1.74E-05** | 1.0381 | 0.8067 | 0.0000 | 1 |
| WDR5B | -1.4179 | **6.24E-06** | 1.0591 | 0.4636 | 0.0000 | 1 |
| WDR74 | -1.1367 | **0.0280** | NA | NA | 0.0000 | 1 |
| WDYHV1 | 1.2073 | **0.0039** | NA | NA | **0.0569** | **97** |
| WEE1 | -2.1517 | **3.48E-11** | 1.4149 | **0.0049** | 0.0000 | 2 |
| WIPF1 | 1.5661 | **0.0001** | -1.0148 | 0.8948 | 0.0008 | 8 |
| WRAP53 | -1.2511 | **1.49E-05** | -1.0287 | 0.6754 | 0.0000 | 1 |
| WRN | -1.1933 | **0.0230** | NA | NA | 0.0002 | 5 |
| WSB1 | 1.2432 | **0.0113** | NA | NA | 0.0018 | 3 |
| WSB2 | -1.4603 | **8.00E-07** | 1.1446 | 0.2044 | 0.0000 | 1 |
| WTAP | -1.2200 | **0.0005** | -1.0728 | 0.3993 | 0.0004 | 6 |
| WWC1 | NA | NA | 1.4413 | **0.0386** | 0.0011 | 7 |
| WWC2 | 1.4255 | **3.85E-05** | 1.1974 | 0.1444 | 0.0000 | 1 |
| WWP1 | 1.1733 | **0.0448** | NA | NA | 0.0013 | 7 |
| XDH | -2.5311 | **0.0064** | NA | NA | 0.0000 | 1 |
| XPA | 1.3412 | **1.06E-05** | NA | NA | 0.0022 | 8 |
| XPNPEP1 | -1.3542 | **2.00E-11** | 1.0086 | 0.9263 | 0.0000 | 1 |
| XPO6 | -1.2776 | **4.97E-06** | 1.0580 | 0.6169 | 0.0000 | 1 |
| XPO7 | -1.1481 | **0.0053** | 1.0567 | 0.5558 | 0.0000 | 1 |
| XRCC3 | -1.1385 | **0.0500** | -1.0792 | 0.6198 | 0.0000 | 1 |
| XRCC5 | -1.4524 | **3.92E-11** | 1.0396 | 0.8055 | 0.0000 | 3 |
| XRCC6 | -1.3162 | **4.05E-09** | NA | NA | **0.0044** | 10 |
| YARS | 1.1587 | **0.0308** | 1.0616 | 0.6819 | 0.0000 | 1 |
| YEATS4 | -1.4097 | **0.0017** | -1.0268 | 0.8645 | 0.0000 | 3 |
| YES1 | -1.2006 | **0.0244** | NA | NA | **0.0052** | **19** |
| YIF1A | -1.4271 | **4.05E-06** | NA | NA | **0.0084** | **18** |
| YIPF1 | -1.1522 | **0.0488** | -1.0725 | 0.4266 | 0.0000 | 5 |
| YIPF6 | -1.1252 | **0.0449** | 1.3248 | **0.0040** | 0.0024 | **15** |
| YJU2 | 1.1271 | **0.0212** | NA | NA | 0.0015 | 6 |
| YKT6 | -1.2746 | **0.0001** | NA | NA | 0.0000 | 1 |
| YPEL5 | 1.2821 | **0.0003** | NA | NA | 0.0001 | 4 |
| YTHDF1 | -1.1543 | **0.0001** | -1.0161 | 0.8638 | 0.0000 | 3 |
| YWHAB | -1.2197 | **0.0009** | 1.1737 | **0.0233** | **0.0045** | **14** |
| YWHAE | -1.6931 | **1.28E-15** | -1.0782 | 0.4543 | **0.0061** | **14** |
| YWHAQ | -1.1799 | **0.0110** | -1.0820 | 0.2589 | **0.0028** | 8 |
| YWHAZ | -1.2899 | **0.0001** | 1.1283 | 0.0891 | **0.0147** | **28** |
| ZBED1 | -1.2109 | **0.0129** | NA | NA | **0.0053** | **20** |
| ZBTB1 | -1.1619 | **0.0442** | NA | NA | 0.0002 | 4 |
| ZBTB10 | -2.3026 | **3.24E-10** | 1.2169 | 0.1133 | 0.0002 | 6 |
| ZBTB14 | 1.1966 | **0.0061** | NA | NA | 0.0006 | 8 |
| ZBTB24 | -1.1630 | **0.0096** | NA | NA | 0.0011 | 12 |
| ZBTB25 | 1.1374 | **0.0338** | 1.0319 | 0.7524 | 0.0011 | 7 |
| ZBTB3 | -1.2309 | **0.0129** | NA | NA | 0.0000 | 1 |
| ZBTB39 | -1.0280 | 0.7363 | -1.1866 | **0.0330** | 0.0003 | 4 |
| ZBTB5 | -1.1976 | **0.0001** | -1.0738 | 0.3366 | 0.0008 | 2 |
| ZBTB6 | -1.0583 | 0.3853 | 1.2852 | **0.0189** | 0.0001 | 2 |
| ZC2HC1A | 1.1566 | **0.0304** | NA | NA | 0.0002 | 5 |
| ZC2HC1C | -1.2846 | **0.0003** | NA | NA | **0.0030** | **26** |
| ZC4H2 | 1.2299 | **0.0039** | -1.0869 | 0.3972 | 0.0001 | 5 |
| ZDHHC17 | 1.3492 | **0.0003** | NA | NA | **0.0060** | **15** |
| ZDHHC24 | -1.1408 | **0.0449** | NA | NA | 0.0024 | 6 |
| ZEB2 | NA | NA | 1.3550 | **0.0489** | 0.0000 | 2 |
| ZFHX3 | -1.4423 | **5.84E-06** | 1.1261 | 0.3317 | 0.0008 | 7 |
| ZFP36L1 | 1.4121 | **0.0259** | -1.3110 | 0.0919 | 0.0000 | 1 |
| ZFP64 | 1.2431 | **2.94E-05** | NA | NA | 0.0000 | 3 |
| ZFPM2 | 9.4136 | **3.17E-28** | -1.0578 | 0.7068 | 0.0000 | 2 |
| ZFYVE26 | 1.2309 | **0.0004** | -1.0864 | 0.4016 | 0.0009 | 9 |
| ZG16 | NA | NA | -1.6889 | **0.0044** | 0.0004 | 3 |
| ZHX2 | 1.3956 | **2.63E-05** | -1.1073 | 0.1994 | 0.0000 | 1 |
| ZMAT3 | 1.5011 | **0.0001** | 1.1976 | 0.0568 | 0.0001 | 3 |
| ZMIZ2 | -1.1223 | **0.0261** | -1.1921 | **0.0332** | 0.0004 | 7 |
| ZMYM2 | -1.2733 | **0.0065** | NA | NA | 0.0017 | 6 |
| ZMYND10 | NA | NA | 1.5044 | **0.0113** | 0.0002 | 2 |
| ZNF124 | -1.2391 | **0.0459** | 1.2777 | **0.0482** | 0.0007 | 11 |
| ZNF131 | -1.2671 | **0.0036** | NA | NA | 0.0000 | 1 |
| ZNF135 | -1.1020 | 0.1790 | -1.1772 | **0.0331** | 0.0000 | 2 |
| ZNF136 | 1.3047 | **0.0002** | NA | NA | 0.0001 | 2 |
| ZNF140 | -1.2324 | **0.0170** | 1.3079 | **0.0173** | 0.0000 | 1 |
| ZNF148 | 1.2891 | **0.0002** | 1.3280 | **0.0224** | 0.0009 | 7 |
| ZNF16 | 1.2032 | **0.0003** | -1.0056 | 0.9533 | 0.0008 | 2 |
| ZNF175 | 1.2759 | **0.0011** | -1.0654 | 0.4364 | 0.0001 | 3 |
| ZNF185 | -1.5990 | **0.0001** | 1.0823 | 0.6560 | 0.0000 | 1 |
| ZNF202 | -1.1059 | **0.0475** | -1.0616 | 0.4307 | 0.0003 | 5 |
| ZNF205 | 1.2211 | **0.0007** | -1.0957 | 0.5254 | 0.0002 | 3 |
| ZNF207 | -1.1274 | **0.0115** | 1.0187 | 0.8676 | 0.0000 | 1 |
| ZNF212 | -1.1518 | **0.0117** | -1.1004 | 0.1869 | 0.0001 | 4 |
| ZNF219 | 1.4478 | **0.0002** | NA | NA | 0.0000 | 2 |
| ZNF224 | 1.2649 | **0.0026** | NA | NA | 0.0000 | 1 |
| ZNF239 | -1.4627 | **0.0001** | 1.0224 | 0.8515 | 0.0000 | 2 |
| ZNF253 | -1.6195 | **4.74E-07** | 1.3511 | **0.0093** | 0.0000 | 1 |
| ZNF264 | 1.1610 | **0.0380** | 1.1656 | 0.2392 | 0.0009 | 3 |
| ZNF280D | 1.4498 | **0.0002** | NA | NA | 0.0000 | 2 |
| ZNF3 | -1.2582 | **1.09E-05** | 1.2025 | 0.1887 | 0.0002 | 5 |
| ZNF34 | 1.2699 | **0.0009** | 1.0216 | 0.8291 | 0.0010 | **15** |
| ZNF410 | -1.1748 | **0.0347** | -1.0245 | 0.7575 | **0.0037** | **16** |
| ZNF446 | -1.1145 | **0.0334** | 1.0475 | 0.6772 | 0.0014 | 10 |
| ZNF451 | 1.2540 | **0.0016** | NA | NA | 0.0018 | 4 |
| ZNF473 | -1.2215 | **2.65E-05** | -1.0214 | 0.8272 | 0.0000 | 4 |
| ZNF512B | -1.1931 | **0.0183** | -1.0138 | 0.9165 | 0.0004 | 4 |
| ZNF544 | -1.2867 | **7.58E-07** | 1.0721 | 0.2491 | 0.0000 | 2 |
| ZNF552 | -2.0270 | **9.37E-09** | 1.0557 | 0.6553 | 0.0001 | 5 |
| ZNF580 | 1.2169 | **0.0008** | -1.1575 | 0.2971 | 0.0001 | 4 |
| ZNF587 | -1.3707 | **1.18E-05** | 1.0664 | 0.5732 | **0.0062** | **29** |
| ZNF613 | -1.1563 | **0.0059** | 1.0890 | 0.4385 | 0.0000 | 1 |
| ZNF629 | -1.1372 | **0.0076** | NA | NA | 0.0000 | 4 |
| ZNF669 | -1.1672 | **0.0302** | 1.1291 | 0.0625 | 0.0000 | 4 |
| ZNF688 | 1.1401 | **0.0348** | -1.2261 | **0.0312** | **0.0077** | **38** |
| ZNF711 | -1.3106 | **0.0044** | 1.1660 | 0.2452 | 0.0000 | 1 |
| ZNF76 | -1.0981 | **0.0474** | 1.0438 | 0.6772 | **0.0064** | **27** |
| ZNF764 | -1.1239 | **0.0130** | NA | NA | 0.0004 | 8 |
| ZNF79 | -1.2470 | **0.0025** | -1.0623 | 0.5240 | 0.0001 | 3 |
| ZNF8 | -1.1974 | **0.0002** | NA | NA | 0.0000 | 1 |
| ZNF821 | NA | NA | -1.1377 | **0.0483** | 0.0000 | 3 |
| ZNHIT1 | -1.1628 | **0.0024** | -1.0876 | 0.3962 | 0.0006 | 3 |
| ZRSR2 | 1.2906 | **2.36E-06** | 1.1231 | 0.2040 | 0.0010 | 6 |
| ZSCAN16 | -1.4980 | **9.86E-08** | 1.1321 | 0.1898 | 0.0001 | 3 |
| ZSCAN26 | 1.3629 | **0.0001** | NA | NA | 0.0000 | 1 |
| ZSCAN9 | -1.1860 | **0.0012** | NA | NA | 0.0002 | 5 |
