## Supplemental Table 4 for "Systems Pharmacology Model Predicts Zinc and Copper Can Be Repurposed as Endometriosis Therapies"

**Supplemental Table 4: Drugs parameters**

| **Drug ID** | **Drug name** | **Degree** | **Betweenness** | **Promiscuity** | **Progression targets** | **Infertility targets** |
| --- | --- | --- | --- | --- | --- | --- |
| DB00025 | Antihemophilic factor, human recombinant | 0 | 1 | 7 | 6 | 1 |
| DB00031 | Tenecteplase | 0 | 1 | 5 | 3 | 2 |
| DB00042 | Botulinum Toxin Type B | 0 | 1 | 0 | 0 | 0 |
| DB00052 | Somatotropin | 0 | 1 | 0 | 0 | 0 |
| DB00082 | Pegvisomant | 0 | 1 | 0 | 0 | 0 |
| DB00091 | Ciclosporin | 0 | 1 | 0 | 0 | 0 |
| DB00098 | Antithymocyte immunoglobulin (rabbit) | 0 | 1 | 0 | 0 | 0 |
| DB00100 | Coagulation Factor IX (Recombinant) | 0 | 1 | 0 | 0 | 0 |
| DB00114 | Pyridoxal phosphate | 0 | 0 | 8 | 5 | 3 |
| DB00126 | Ascorbic acid | 0 | 1 | 4 | 3 | 1 |
| DB00128 | Aspartic acid | 0 | 1 | 5 | 5 | 0 |
| DB00131 | Adenosine phosphate | 0 | 1 | 0 | 0 | 0 |
| DB00132 | Alpha-Linolenic Acid | 1 | 1 | 0 | 0 | 0 |
| DB00139 | Succinic acid | 0 | 1 | 0 | 0 | 0 |
| DB00142 | Glutamic Acid | 0 | 0 | 6 | 6 | 0 |
| DB00143 | Glutathione | 1 | 1 | 8 | 8 | 0 |
| DB00145 | Glycine | 0 | 0 | 6 | 5 | 1 |
| DB00157 | NADH | 2 | 3 | 22 | 22 | 4 |
| DB00159 | Icosapent | 1 | 1 | 0 | 0 | 0 |
| DB00172 | Proline | 0 | 1 | 4 | 4 | 0 |
| DB00173 | Adenine | 1 | 1 | 5 | 4 | 1 |
| DB00180 | Flunisolide | 1 | 1 | 0 | 0 | 0 |
| DB00188 | Bortezomib | 1 | 1 | 0 | 0 | 0 |
| DB00210 | Adapalene | 1 | 1 | 0 | 0 | 0 |
| DB00223 | Diflorasone | 1 | 1 | 0 | 0 | 0 |
| DB00240 | Alclometasone | 1 | 1 | 0 | 0 | 0 |
| DB00244 | Mesalazine | 1 | 1 | 0 | 0 | 0 |
| DB00253 | Medrysone | 1 | 1 | 0 | 0 | 0 |
| DB00277 | Theophylline | 0 | 1 | 0 | 0 | 0 |
| DB00279 | Liothyronine | 1 | 1 | 0 | 0 | 0 |
| DB00288 | Amcinonide | 1 | 1 | 0 | 0 | 0 |
| DB00313 | Valproic Acid | 1 | 1 | 0 | 0 | 0 |
| DB00324 | Fluorometholone | 1 | 1 | 0 | 0 | 0 |
| DB00328 | Indometacin | 1 | 1 | 0 | 0 | 0 |
| DB00351 | Megestrol acetate | 1 | 1 | 0 | 0 | 0 |
| DB00394 | Beclomethasone dipropionate | 1 | 1 | 0 | 0 | 0 |
| DB00396 | Progesterone | 1 | 1 | 0 | 0 | 0 |
| DB00398 | Sorafenib | 1 | 1 | 5 | 5 | 0 |
| DB00412 | Rosiglitazone | 1 | 1 | 0 | 0 | 0 |
| DB00421 | Spironolactone | 1 | 1 | 0 | 0 | 0 |
| DB00443 | Betamethasone | 1 | 1 | 0 | 0 | 0 |
| DB00459 | Acitretin | 1 | 1 | 0 | 0 | 0 |
| DB00477 | Chlorpromazine | 0 | 1 | 0 | 0 | 0 |
| DB00523 | Alitretinoin | 1 | 1 | 0 | 0 | 0 |
| DB00527 | Cinchocaine | 0 | 1 | 0 | 0 | 0 |
| DB00536 | Guanidine | 0 | 1 | 0 | 0 | 0 |
| DB00547 | Desoximetasone | 1 | 1 | 0 | 0 | 0 |
| DB00570 | Vinblastine | 1 | 1 | 0 | 0 | 0 |
| DB00573 | Fenoprofen | 1 | 1 | 0 | 0 | 0 |
| DB00588 | Fluticasone propionate | 1 | 1 | 0 | 0 | 0 |
| DB00591 | Fluocinolone acetonide | 1 | 1 | 0 | 0 | 0 |
| DB00596 | Ulobetasol | 1 | 1 | 0 | 0 | 0 |
| DB00615 | Rifabutin | 1 | 1 | 0 | 0 | 0 |
| DB00620 | Triamcinolone | 1 | 1 | 0 | 0 | 0 |
| DB00622 | Nicardipine | 0 | 1 | 0 | 0 | 0 |
| DB00623 | Fluphenazine | 0 | 1 | 0 | 0 | 0 |
| DB00635 | Prednisone | 1 | 1 | 0 | 0 | 0 |
| DB00663 | Flumethasone | 1 | 1 | 0 | 0 | 0 |
| DB00675 | Tamoxifen | 1 | 1 | 0 | 0 | 0 |
| DB00687 | Fludrocortisone | 1 | 1 | 0 | 0 | 0 |
| DB00716 | Nedocromil | 1 | 1 | 0 | 0 | 0 |
| DB00731 | Nateglinide | 1 | 1 | 0 | 0 | 0 |
| DB00741 | Hydrocortisone | 1 | 1 | 0 | 0 | 0 |
| DB00746 | Deferoxamine | 1 | 1 | 0 | 0 | 0 |
| DB00753 | Isoflurane | 0 | 1 | 0 | 0 | 0 |
| DB00755 | Tretinoin | 1 | 1 | 5 | 5 | 0 |
| DB00759 | Tetracycline | 0 | 1 | 0 | 0 | 0 |
| DB00764 | Mometasone | 1 | 1 | 0 | 0 | 0 |
| DB00769 | Hydrocortamate | 1 | 1 | 0 | 0 | 0 |
| DB00795 | Sulfasalazine | 1 | 1 | 0 | 0 | 0 |
| DB00799 | Tazarotene | 1 | 1 | 0 | 0 | 0 |
| DB00819 | Acetazolamide | 1 | 1 | 0 | 0 | 0 |
| DB00831 | Trifluoperazine | 0 | 1 | 0 | 0 | 0 |
| DB00834 | Mifepristone | 1 | 1 | 0 | 0 | 0 |
| DB00836 | Loperamide | 0 | 1 | 0 | 0 | 0 |
| DB00838 | Clocortolone | 1 | 1 | 0 | 0 | 0 |
| DB00846 | Flurandrenolide | 1 | 1 | 0 | 0 | 0 |
| DB00850 | Perphenazine | 0 | 1 | 0 | 0 | 0 |
| DB00860 | Prednisolone | 1 | 1 | 0 | 0 | 0 |
| DB00895 | Benzylpenicilloyl Polylysine | 1 | 1 | 0 | 0 | 0 |
| DB00896 | Rimexolone | 1 | 1 | 0 | 0 | 0 |
| DB00898 | Ethanol | 1 | 1 | 0 | 0 | 0 |
| DB00912 | Repaglinide | 1 | 1 | 0 | 0 | 0 |
| DB00914 | Phenformin | 0 | 1 | 0 | 0 | 0 |
| DB00925 | Phenoxybenzamine | 0 | 1 | 0 | 0 | 0 |
| DB00945 | Acetylsalicylic acid | 1 | 3 | 8 | 8 | 0 |
| DB00959 | Methylprednisolone | 1 | 1 | 0 | 0 | 0 |
| DB00966 | Telmisartan | 1 | 1 | 0 | 0 | 0 |
| DB00982 | Isotretinoin | 1 | 1 | 0 | 0 | 0 |
| DB01013 | Clobetasol propionate | 1 | 1 | 0 | 0 | 0 |
| DB01014 | Balsalazide | 1 | 1 | 0 | 0 | 0 |
| DB01016 | Glyburide | 0 | 1 | 0 | 0 | 0 |
| DB01023 | Felodipine | 0 | 1 | 0 | 0 | 0 |
| DB01029 | Irbesartan | 1 | 1 | 0 | 0 | 0 |
| DB01039 | Fenofibrate | 1 | 1 | 0 | 0 | 0 |
| DB01041 | Thalidomide | 0 | 1 | 0 | 0 | 0 |
| DB01047 | Fluocinonide | 1 | 1 | 0 | 0 | 0 |
| DB01050 | Ibuprofen | 1 | 1 | 0 | 0 | 0 |
| DB01064 | Isoprenaline | 1 | 1 | 0 | 0 | 0 |
| DB01065 | Melatonin | 0 | 1 | 0 | 0 | 0 |
| DB01067 | Glipizide | 1 | 1 | 0 | 0 | 0 |
| DB01069 | Promethazine | 0 | 1 | 0 | 0 | 0 |
| DB01097 | Leflunomide | 0 | 1 | 0 | 0 | 0 |
| DB01100 | Pimozide | 0 | 1 | 0 | 0 | 0 |
| DB01110 | Miconazole | 0 | 0 | 4 | 4 | 1 |
| DB01115 | Nifedipine | 0 | 1 | 0 | 0 | 0 |
| DB01118 | Amiodarone | 1 | 1 | 0 | 0 | 0 |
| DB01130 | Prednicarbate | 1 | 1 | 0 | 0 | 0 |
| DB01132 | Pioglitazone | 1 | 1 | 0 | 0 | 0 |
| DB01136 | Carvedilol | 1 | 2 | 4 | 4 | 1 |
| DB01169 | Arsenic trioxide | 3 | 4 | 7 | 7 | 1 |
| DB01185 | Fluoxymesterone | 1 | 1 | 0 | 0 | 0 |
| DB01222 | Budesonide | 1 | 1 | 0 | 0 | 0 |
| DB01234 | Dexamethasone | 1 | 1 | 0 | 0 | 0 |
| DB01244 | Bepridil | 0 | 1 | 0 | 0 | 0 |
| DB01254 | Dasatinib | 1 | 3 | 7 | 7 | 1 |
| DB01260 | Desonide | 1 | 1 | 0 | 0 | 0 |
| DB01327 | Cefazolin | 0 | 1 | 0 | 0 | 0 |
| DB01370 | Aluminium | 1 | 1 | 0 | 0 | 0 |
| DB01373 | Calcium | 0 | 1 | 7 | 7 | 1 |
| DB01380 | Cortisone acetate | 1 | 1 | 0 | 0 | 0 |
| DB01393 | Bezafibrate | 1 | 1 | 0 | 0 | 0 |
| DB01410 | Ciclesonide | 1 | 1 | 0 | 0 | 0 |
| DB01592 | Iron | 0 | 0 | 4 | 2 | 3 |
| DB01593 | Zinc | 2 | 4 | 22 | 21 | 5 |
| DB01599 | Probucol | 0 | 1 | 0 | 0 | 0 |
| DB02266 | Flufenamic Acid | 1 | 1 | 0 | 0 | 0 |
| DB02701 | Nicotinamide | 0 | 2 | 0 | 0 | 0 |
| DB02709 | Resveratrol | 3 | 3 | 9 | 9 | 3 |
| DB03147 | Flavin adenine dinucleotide | 1 | 0 | 7 | 6 | 1 |
| DB03756 | Doconexent | 1 | 1 | 0 | 0 | 0 |
| DB04224 | Oleic Acid | 1 | 1 | 0 | 0 | 0 |
| DB04272 | Citric acid | 1 | 1 | 5 | 5 | 0 |
| DB04825 | Prenylamine | 0 | 1 | 0 | 0 | 0 |
| DB04841 | Flunarizine | 0 | 1 | 0 | 0 | 0 |
| DB06616 | Bosutinib | 0 | 0 | 4 | 2 | 2 |
| DB06773 | Human calcitonin | 0 | 1 | 0 | 0 | 0 |
| DB06781 | Difluprednate | 1 | 1 | 0 | 0 | 0 |
| DB06782 | Dimercaprol | 1 | 1 | 0 | 0 | 0 |
| DB08604 | Triclosan | 1 | 1 | 0 | 0 | 0 |
| DB08813 | Nadroparin | 1 | 1 | 0 | 0 | 0 |
| DB08814 | Triflusal | 0 | 1 | 0 | 0 | 0 |
| DB08862 | Cholecystokinin | 1 | 1 | 0 | 0 | 0 |
| DB08867 | Ulipristal | 1 | 1 | 0 | 0 | 0 |
| DB08889 | Carfilzomib | 1 | 1 | 4 | 4 | 1 |
| DB08896 | Regorafenib | 1 | 1 | 9 | 8 | 1 |
| DB08901 | Ponatinib | 0 | 0 | 5 | 5 | 0 |
| DB08906 | Fluticasone furoate | 1 | 1 | 0 | 0 | 0 |
| DB08908 | Dimethyl fumarate | 1 | 1 | 0 | 0 | 0 |
| DB08912 | Dabrafenib | 1 | 1 | 0 | 0 | 0 |
| DB09061 | Cannabidiol | 1 | 1 | 0 | 0 | 0 |
| DB09074 | Olaparib | 0 | 1 | 0 | 0 | 0 |
| DB09091 | Tixocortol | 1 | 1 | 0 | 0 | 0 |
| DB09092 | Xanthinol | 0 | 1 | 0 | 0 | 0 |
| DB09095 | Difluocortolone | 1 | 1 | 0 | 0 | 0 |
| DB09098 | Somatrem | 0 | 1 | 0 | 0 | 0 |
| DB09118 | Stiripentol | 0 | 1 | 0 | 0 | 0 |
| DB09130 | Copper | 6 | 11 | 44 | 40 | 10 |
| DB09148 | Florbetaben (18F) | 1 | 1 | 0 | 0 | 0 |
| DB09149 | Florbetapir (18F) | 1 | 1 | 0 | 0 | 0 |
| DB09151 | Flutemetamol (18F) | 1 | 1 | 0 | 0 | 0 |
| DB09213 | Dexibuprofen | 1 | 1 | 0 | 0 | 0 |
| DB09401 | Isosorbide | 1 | 1 | 0 | 0 | 0 |
| DB09462 | Glycerin | 0 | 0 | 6 | 6 | 0 |
| DB11093 | Calcium Citrate | 0 | 2 | 9 | 8 | 2 |
| DB11120 | Turpentine | 1 | 1 | 0 | 0 | 0 |
| DB11133 | Omega-3 fatty acids | 1 | 1 | 0 | 0 | 0 |
| DB11338 | Clove oil | 1 | 1 | 0 | 0 | 0 |
| DB11348 | Calcium Phosphate | 0 | 2 | 9 | 8 | 2 |
| DB11619 | Gestrinone | 1 | 1 | 0 | 0 | 0 |
| DB11672 | Curcumin | 1 | 1 | 0 | 0 | 0 |
| DB11760 | Talazoparib | 0 | 1 | 0 | 0 | 0 |
| DB11793 | Niraparib | 0 | 1 | 0 | 0 | 0 |
| DB11817 | Baricitinib | 0 | 1 | 0 | 0 | 0 |
| DB11921 | Deflazacort | 1 | 1 | 0 | 0 | 0 |
| DB12010 | Fostamatinib | 6 | 10 | 86 | 73 | 17 |
| DB12332 | Rucaparib | 0 | 1 | 0 | 0 | 0 |
| DB13152 | Coagulation Factor IX Human | 0 | 1 | 0 | 0 | 0 |
| DB13158 | Clobetasone | 1 | 1 | 0 | 0 | 0 |
| DB13800 | Calcium levulinate | 0 | 1 | 0 | 0 | 0 |
| DB13867 | Fluticasone | 1 | 1 | 0 | 0 | 0 |
| DB13873 | Fenofibric acid | 1 | 1 | 0 | 0 | 0 |
| DB13961 | Fish oil | 1 | 1 | 0 | 0 | 0 |
| DB13998 | Lonoctocog alfa | 0 | 1 | 7 | 6 | 1 |
| DB13999 | Moroctocog alfa | 0 | 1 | 7 | 6 | 1 |
| DB14481 | Calcium phosphate dihydrate | 0 | 2 | 9 | 8 | 2 |
| DB14487 | Zinc acetate | 2 | 4 | 22 | 21 | 5 |
| DB14488 | Ferrous gluconate | 0 | 0 | 4 | 2 | 3 |
| DB14489 | Ferrous succinate | 0 | 0 | 4 | 2 | 3 |
| DB14490 | Ferrous ascorbate | 0 | 0 | 4 | 2 | 3 |
| DB14491 | Ferrous fumarate | 0 | 0 | 4 | 2 | 3 |
| DB14501 | Ferrous glycine sulfate | 0 | 0 | 4 | 2 | 3 |
| DB14512 | Mometasone furoate | 1 | 1 | 0 | 0 | 0 |
| DB14517 | Aluminium phosphate | 1 | 1 | 0 | 0 | 0 |
| DB14518 | Aluminum acetate | 1 | 1 | 0 | 0 | 0 |
| DB14533 | Zinc chloride | 2 | 4 | 22 | 21 | 5 |
| DB14539 | Hydrocortisone acetate | 1 | 1 | 0 | 0 | 0 |
| DB14540 | Hydrocortisone butyrate | 1 | 1 | 0 | 0 | 0 |
| DB14541 | Hydrocortisone cypionate | 1 | 1 | 0 | 0 | 0 |
| DB14542 | Hydrocortisone phosphate | 1 | 1 | 0 | 0 | 0 |
| DB14543 | Hydrocortisone probutate | 1 | 1 | 0 | 0 | 0 |
| DB14544 | Hydrocortisone valerate | 1 | 1 | 0 | 0 | 0 |
| DB14596 | Loteprednol etabonate | 1 | 1 | 0 | 0 | 0 |
| DB14731 | Tagraxofusp | 0 | 1 | 0 | 0 | 0 |
