## Supplemental Table 5 for "Systems Pharmacology Model Predicts Zinc and Copper Can Be Repurposed as Endometriosis Therapies"

**Supplemental Table 5. Classification of current endometriosis drugs.**

| **Endometriosis Drug** | **Type** | **Indication** |
| --- | --- | --- |
| Acetaminophen | Analgesic | Pain |
| Amitriptyline | Analgesic | Pain |
| Anastrozole | Aromatase inhibitor | Pain |
| Buserelin | GnRHa | Pain |
| Chorionic Gonadotropin (Human) | COS | Infertility |
| Clomifene | COS | Infertility |
| Cyproterone acetate | Progestin | Pain |
| Danazol | Aromatase inhibitor | Pain |
| Desogestrel | Progestin | Pain |
| Dienogest | Progestin | Pain |
| Duloxetine | Analgesic | Pain |
| Dydrogesterone | Progestin | Pain |
| Elagolix | GnRHant | Pain |
| Estradiol | Estrogen | Pain |
| Ethinylestradiol | Estrogen | Pain |
| Etonogestrel | Progestin | Pain |
| Exemestane | Aromatase inhibitor | Pain |
| Fentanyl | Opioid | Pain |
| Follitropin | COS | Infertility |
| Gabapentin | Analgesic | Pain |
| Gestrinone | Progestin | Pain |
| Goserelin | GnRHa | Pain |
| Ibuprofen | Analgesic | Pain |
| Letrozole | Aromatase inhibitor | Pain |
| Leuprolide | GnRHa | Pain |
| Levonorgestrel | Progestin | Pain |
| Medroxyprogesterone acetate | Progestin | Pain |
| Megestrol acetate | Progestin | Pain |
| Metamizole | Analgesic | Pain |
| Methadone | Opioid | Pain |
| Mifepristone | SPRM | Pain |
| Morphine | Opioid | Pain |
| Nafarelin | GnRHa | Pain |
| Naproxen | Analgesic | Pain |
| Nomegestrol | Progestin | Pain |
| Norelgestromin | Progestin | Pain |
| Norethisterone | Progestin | Pain |
| Pregabalin | Analgesic | Pain |
| Progesterone | Progestin | Pain |
| Rofecoxib | Analgesic | Pain |
| Tibolone | SERM | Pain |
| Triptorelin | GnRHa | Pain |
| Thiamine | Vitamin | Pain |
| Pyridoxine | Vitamin | Pain |

*GnRHa*, gonadotropin-releasing hormone agonist; *GnRHant*, gonadotropin-releasing hormone antagonist; *COS*, control ovarian stimulation; *SPRM*, selective progesterone receptor modulator; *SERM*, selective estrogen receptor modulator
